# A Systematic Review of Human Challenge Trials, Designs, and Safety

**DOI:** 10.1101/2022.03.20.22272658

**Authors:** Jupiter Adams-Phipps, Danny Toomey, Witold Więcek, Virginia Schmit, James Wilkinson, Keller Scholl, Euzebiusz Jamrozik, Joshua Osowicki, Meta Roestenberg, David Manheim

## Abstract

**Background:** There exists no prior systematic review of human challenge trials (HCTs) that focuses on participant safety. Key questions regarding HCTs include how risky such trials have been, how often adverse events (AEs) and serious adverse events (SAEs) occur, and whether risk mitigation measures have been effective.

**Methods:** A systematic search of PubMed and PubMed Central for articles reporting on results of HCTs published between 1980 and 2021 was performed and completed by 10/7/2021.

**Results:** Of 2,838 articles screened, 276 were reviewed in full. 15,046 challenged participants were described in 308 studies that met inclusion criteria. 286 (92.9%) of these studies reported mitigation measures used to minimize risk to the challenge population. Among 187 studies which reported on SAEs, 0.2% of participants experienced at least one challenge-related SAE. Among 94 studies that graded AEs by severity, challenge-related AEs graded “severe” were reported by between 5.6% and 15.8% of participants. AE data were provided as a range to account for unclear reporting. 80% of studies published after 2010 were registered in a trials database.

**Conclusions:** HCTs are increasingly common and used for an expanding list of diseases. Although AEs occur, severe AEs and SAEs are rare. Reporting has improved over time, though not all papers provide a comprehensive report of relevant health impacts. From the available data, most HCTs do not lead to a high number of severe symptoms or SAEs. This study was preregistered on PROSPERO as CRD42021247218.

## 1 Introduction

Human challenge trials (HCTs) are a clinical research method where volunteers are exposed to a pathogen in order to derive scientifically useful information about the pathogen and/or an intervention (1). Such trials have been conducted with ethical oversight since the development of the modern institutional review system of clinical trials in the 1970s. More recently, there has been renewed discussion about the ethical and practical aspects of conducting HCTs, largely fuelled by interest in conducting HCTs for SARS-CoV-2. Some past reviews of HCTs focused on reporting methods (2–4), but these did not explicitly evaluate the safety of HCTs by assessing reported adverse events (AEs) and serious adverse events (SAEs). Furthermore, many additional HCTs have been performed since the publication of these reviews. In order to better inform discussions about future uses of HCTs, including during pandemic response, this article presents a systematic review of challenge trials since 1980 and reports on their clinical outcomes, with particular focus on risk of adverse events and risk mitigation strategies.

HCTs are often used to support development of therapies and vaccines more efficiently than conventional clinical trials (5–8), and have recently been discussed as particularly valuable in the context of novel disease pandemics like COVID-19, Zika virus, or a future Disease X (9–11). HCTs have been used to investigate malaria (12), influenza (13), common cold (14), various enteric diseases (15,16), and cholera (16). The benefits of such trials include defining and evaluating correlates of protection (17); the first FDA-approved cholera vaccine, Vaxchora, which proved its efficacy using a small HCT (7,18); a contribution to the development of the FDA-approved therapeutic oseltamivir for influenza (19); the vi-tetanus toxoid conjugate vaccine for *Salmonella typhi* (20); and dosing schedules for RTS,S/AS01 malaria vaccine (21).

Arguments against the use of HCTs have centered around ethics of participant compensation and the populations represented, and whether the risks and lack of personal benefit can be compatible with the principle of *primum non nocere* (22–25) due to the potential risks they may inflict on a study population. Despite the debate, there is a long-standing consensus that infecting healthy volunteers is ethically justifiable, so long as the risk of harm is acceptably low (24). HCTs can therefore be ethical, based on a case by case assessment of risk as part of wider research ethics oversight mechanisms.

AEs related to challenge are one measure of health risk in HCTs. AEs refer to “any untoward medical occurrence associated with the use of a drug in humans” (26). The US FDA considers challenge agents to be (akin to) investigational new drugs (27), such that AEs in HCTs refer to any untoward medical occurrence associated with the challenge. AEs that result in death, hospitalization, disability, or permanent damage; as well as AEs that are life-threatening or other important medical events, are reported as serious adverse events (SAEs) (26). It should be noted that AEs graded “severe” by studies are distinct from SAEs in most cases, usually because they are not life-threatening or do not require hospitalization.

A systematic review was performed to characterize the frequency and nature of AEs and SAEs in HCTs related to the challenge, and the risk mitigation measures employed. The review also investigated the pathogens studied, the clinical outcomes in participants, study registration in databases, the number and uses of HCTs over time, and the quality of data reporting.

## 2 Methods

### 2.1 Search Strategy

A systematic review of records from 1980 to 2021 indexed in the PubMed and PubMed Central (PMC) databases was performed to identify published articles describing HCTs. Articles published prior to 1980 were not assessed because the modern institutional review system was not in place until after the 1979 Belmont report. The initial search was preregistered on PROSPERO as CRD42021247218 (28), but identified few studies published prior to 2010. Additional searches were performed to address this and appropriately discover studies for each decade of interest, as detailed in the amended preregistration (28) and the Supplementary Methods. The database search strategy is presented in Table 1. Further manual searches of references lists and reviews were performed to identify additional articles describing HCTs that were missed.

**Table 1.**
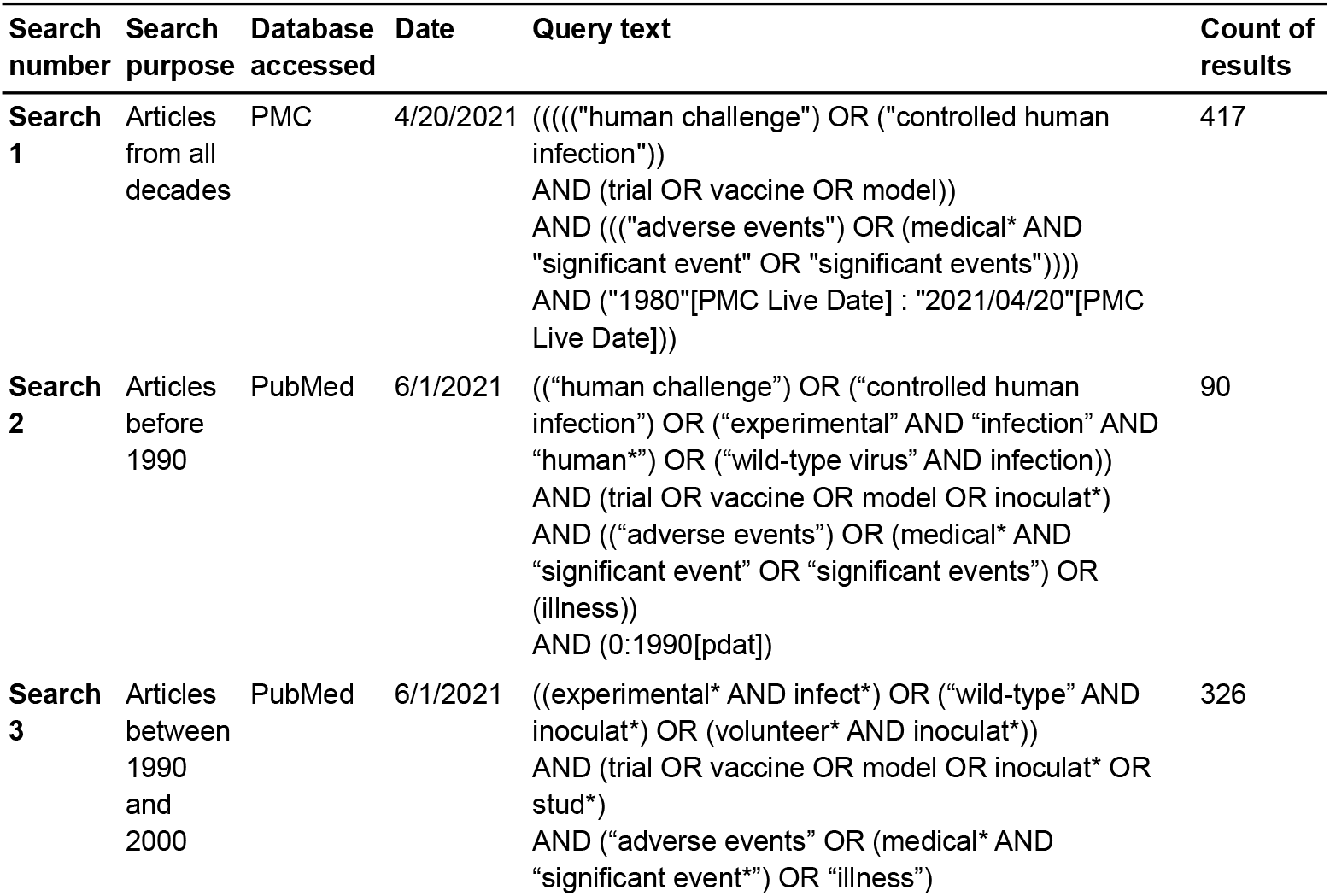
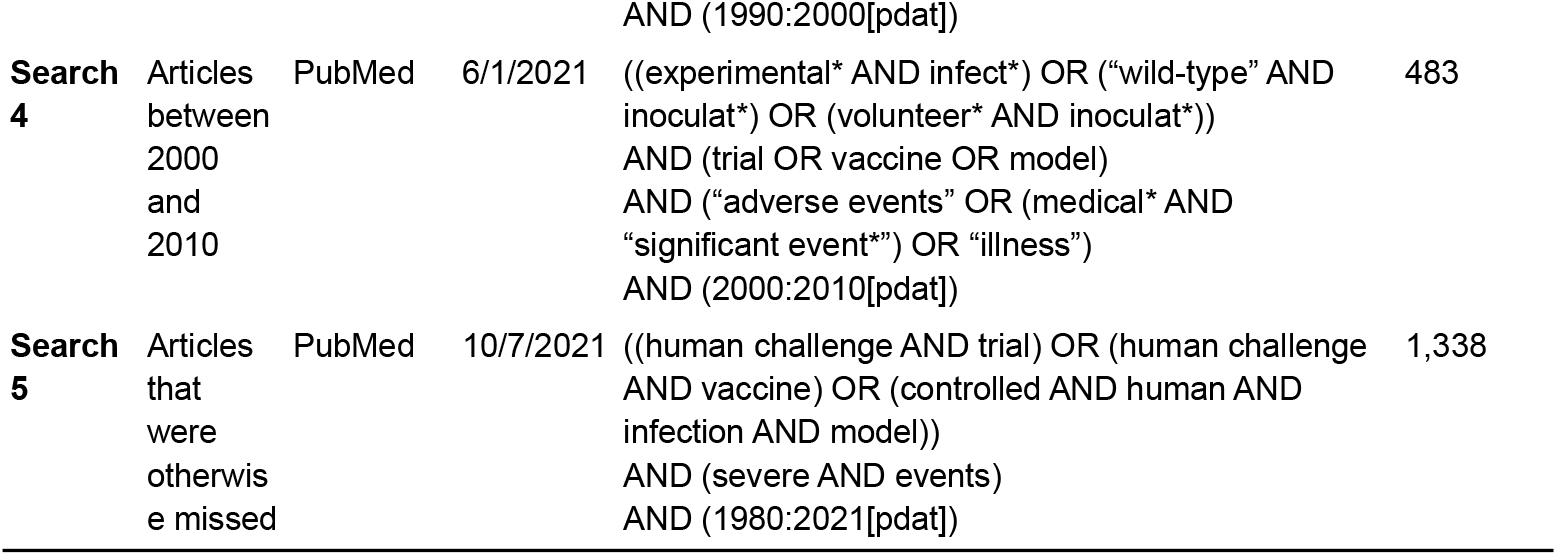
Search Strategy.

### 2.2 Screening Process

Titles and abstracts of search results were manually screened by three authors working independently to identify articles that were eligible for full text review. Case reports, reviews, articles not available in English, studies that did not meet the criteria for an HCT, and articles published prior to 1980 were excluded. Secondary reviews of two past reviews (3,4) were also performed to identify more articles that were missed by the searches. Articles that described studies that performed secondary analysis of results from previously conducted HCTs were excluded, but their reference lists were reviewed to identify the original publication of these results.

### 2.3 Full Text Review Process

The unit of analysis is the individual study, as described within a published article detailing results. Individual studies were identified by trial registration. If trial registration was not reported, studies were counted per the article description, or as a single study if participants were challenged with a single pathogen. If multiple articles were published discussing the same study, the earliest published article was included. In some cases, multiple articles were combined (see Supplementary Methods).

There is an ongoing discussion on the precise definition of an HCT (29). In general, studies that had been completed and involved intentional exposure of human volunteers to a pathogen were included. Challenges with candidate vaccine viruses were also included, as were studies where previously challenged participants were challenged again with the same pathogen (rechallenges). Consistent with Kalil *et al*., studies involving live, attenuated vaccines which were not followed by intentional infection, as well as data from phases of studies involving immunization or vaccination with live, attenuated vaccines or other methods that could have potentially resulted in infection, but which are not generally referred to as HCTs, were excluded (2).

### 2.4 Data Collection Process

At least two reviewers independently examined each publication selected for full text review and any discrepancies were either reconciled, or resolved by the senior author. Data collection was performed manually and results were input into a spreadsheet.

### 2.5 Data Extraction

The following numerical data were extracted from each study: year of article publication, size of cohort, gender breakdowns; mean or median age, standard deviation, and age range; number of participants challenged, number of challenged participants infected with pathogen, number of participants in control group (those who did not undergo a challenge), number of control participants infected with pathogen, number of control participants with at least one AE, and number of challenged participants with: (a) at least one AE, (b) at least one “severe” or “very severe” (grade 3 or higher) AE, (c) at least one SAE.

In addition, the following non-numerical data were extracted from each study: clinical trial registration, pathogen assessed, definition of infection, definition of AEs, treatments administered to participants, risk mitigations taken, ethics committee and review board approvals reported, and a brief description of the study design.

For articles that reported separate study arms that were all exposed to a pathogen within a single pathogen category, data were summed across all arms to be treated as a single study. Data from rechallenges were extracted separately and treated as individual studies. No treatment effect measures were extracted.

AEs among challenged participants that were not related to challenge (such as AEs related to vaccination or drug treatment) were not extracted (see Supplementary Methods). For studies that did not define and/or report AEs, reported symptom data were extracted instead. For studies that did not define and/or report SAEs, reported symptom data that met the 2016 definition of SAEs provided by the FDA (26) based on reviewer judgment were extracted as SAEs.

## 3 Results

### 3.1 Study Selection

Figure 1 shows a PRISMA flowchart of study selection, generated using a tool by Haddaway et al. (30). Searches yielded a total of 2,654 results. 183 additional results were added by citation searching the reference lists of two past reviews (3,4) and articles identified among search results that used data from prior HCTs. One article (31) provided updated data for another (32). 11 results were not retrieved (five with no full text available and six with unpublished data), and 47 duplicates were removed. No further efforts were made to identify unpublished or unidentified work. Results were assessed for eligibility, and 276 articles were included, describing 308 studies from which data were extracted. Excluded results were primarily reviews and articles discussing non-HCT clinical trials. See the Supplementary References for the complete reference list of included articles.

**Figure 1.**
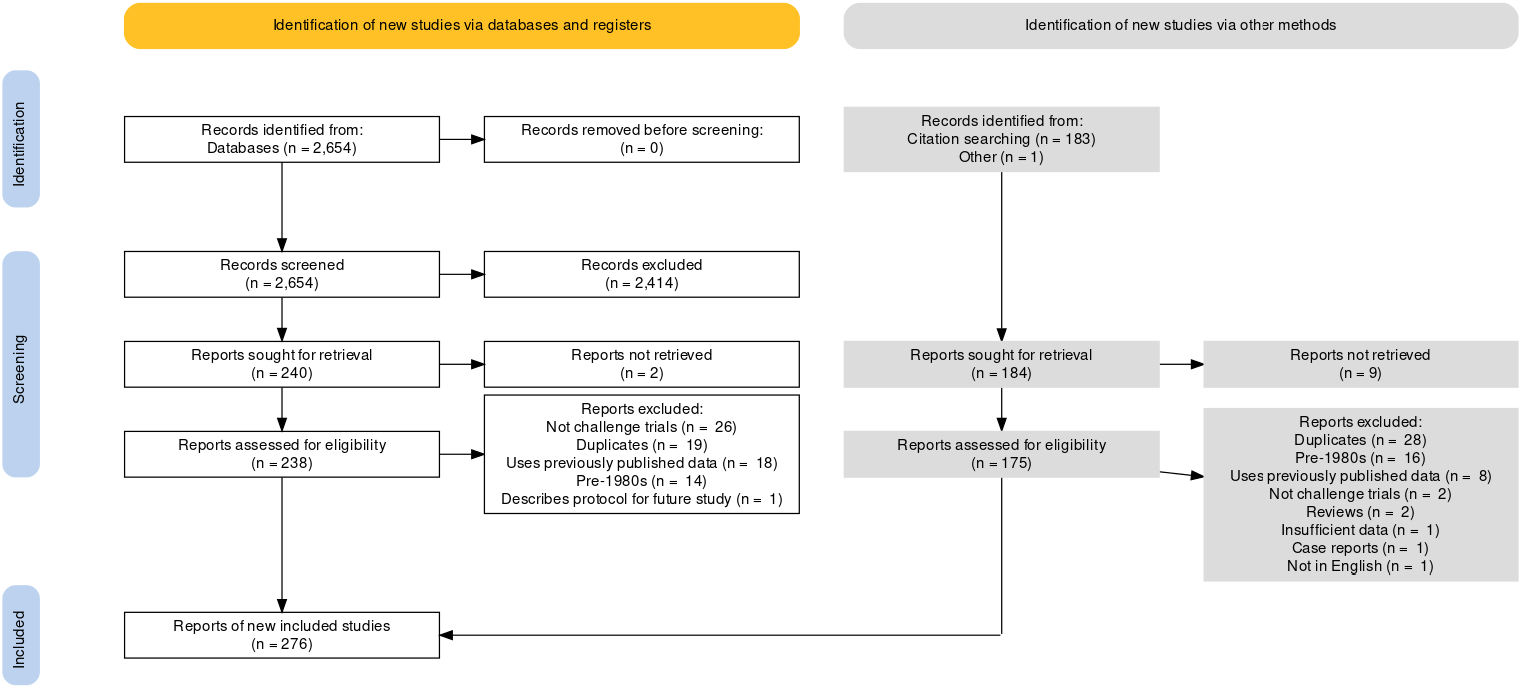
Prisma Flow Diagram.

### 3.2 Results of Individual Studies

Data from 284 studies, with 14,628 challenged participants, were extracted (Table 2). Additional data were extracted from 24 rechallenge studies (Supplementary Table 3). Between 9,917 and 10,277 challenged participants (67.8% −70.3%) were diagnosed with infection. The dataset and code used for generating all results and tables are publicly available (https://github.com/1DaySooner/HCTSystematicReview).

**Table 2.**
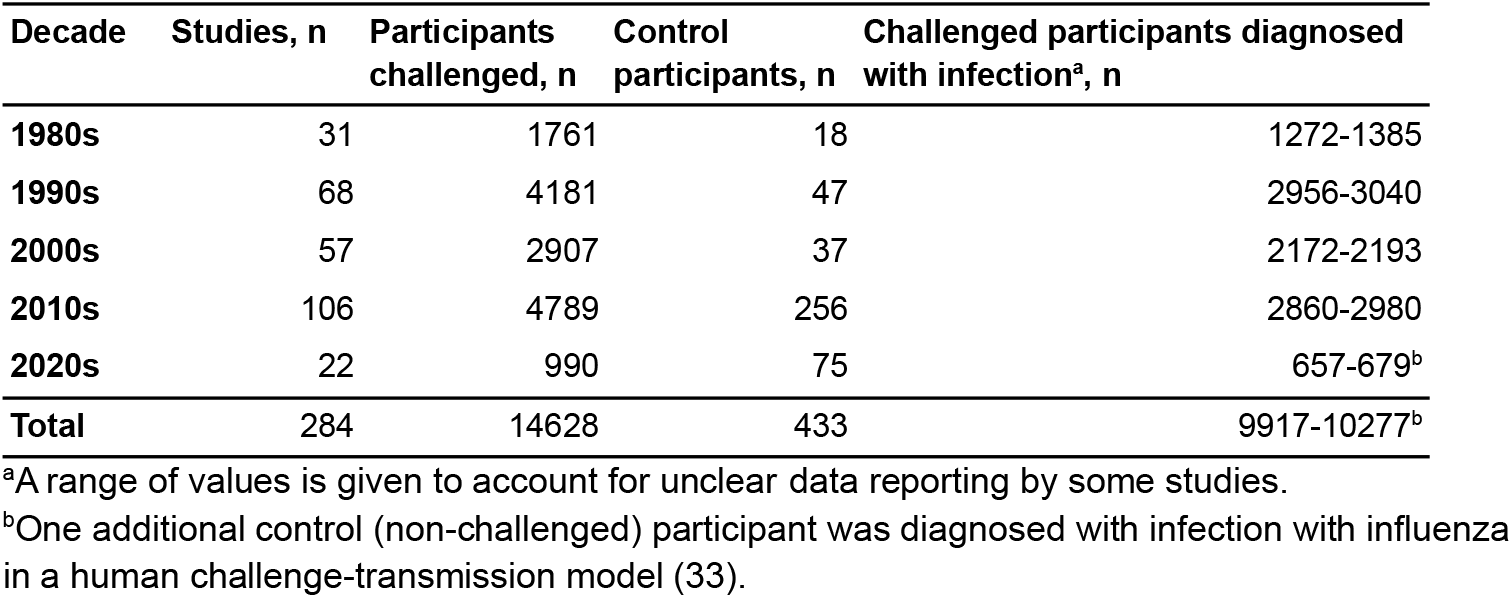
Number of Studies, Number of Participants, and Number of Infections in Published HCTs by Decade.

| Decade | Studies, n | Participants challenged, n | Control participants, n | Challenged participants diagnosed with infection<sup>a</sup>, n |
| --- | --- | --- | --- | --- |
| 1980s | 31 | 1761 | 18 | 1272-1385 |
| 1990s | 68 | 4181 | 47 | 2956-3040 |
| 2000s | 57 | 2907 | 37 | 2172-2193 |
| 2010s | 106 | 4789 | 256 | 2860-2980 |
| 2020s | 22 | 990 | 75 | 657-679<sup>b</sup> |
| **Total** | **284** | **14628** | **433** | **9917-10277<sup>b</sup>** |

<sup>a</sup>A range of values is given to account for unclear data reporting by some studies.
<sup>b</sup>One additional control (non-challenged) participant was diagnosed with infection with influenza in a human challenge-transmission model (33).

**Table 3.** Data Reporting and Database Registration in Published HCTs by Decade.

| **Decade** | **Studies, n** | **Studies that do not define AEs, n (%)** | **Studies with unclear AE data, n (%)** | **Studies with no AE data, n (%)** | **Studies that do not mention SAEs, n (%)** | **Studies with no SAE data, n (%)** |
| --- | --- | --- | --- | --- | --- | --- |
| **1980s** | 31 | 25 (80.6) | 2 (6.5) | 8 (25.8) | 23 (74.2) | 6 (19.4) |
| **1990s** | 68 | 52 (76.5) | 8 (11.9) | 27 (40.3) | 40 (58.8) | 27 (40.3) |
| **2000s** | 57 | 45 (78.9) | 7 (12.3) | 31 (54.4) | 10 (17.5) | 31 (54.4) |
| **2010s** | 106 | 33 (31.1) | 32 (30.2) | 26 (24.5) | 8 (7.5) | 32 (30.2) |
| **2020s** | 22 | 6 (27.3) | 9 (40.9) | 2 (9.1) | 0 (0.0) | 1 (4.5) |
| **Total** | 284 | 161 (56.7) | 58 (20.4) | 94 (33.1) | 81 (28.5) | 97 (34.2) |

### 3.3 Reported Adverse Events and Unreported Data

Among 284 studies, 94 and 97 did not report any AE or SAE data, respectively (Table 3). The precise number of participants experiencing at least one SAE could not be extracted from two studies: one lost some challenged subjects’ original records in a storage facility that flooded (34), and the other only reported that “All serious AEs were self-limited and resolved within several days, and none were deemed to be vaccine-related” (35).

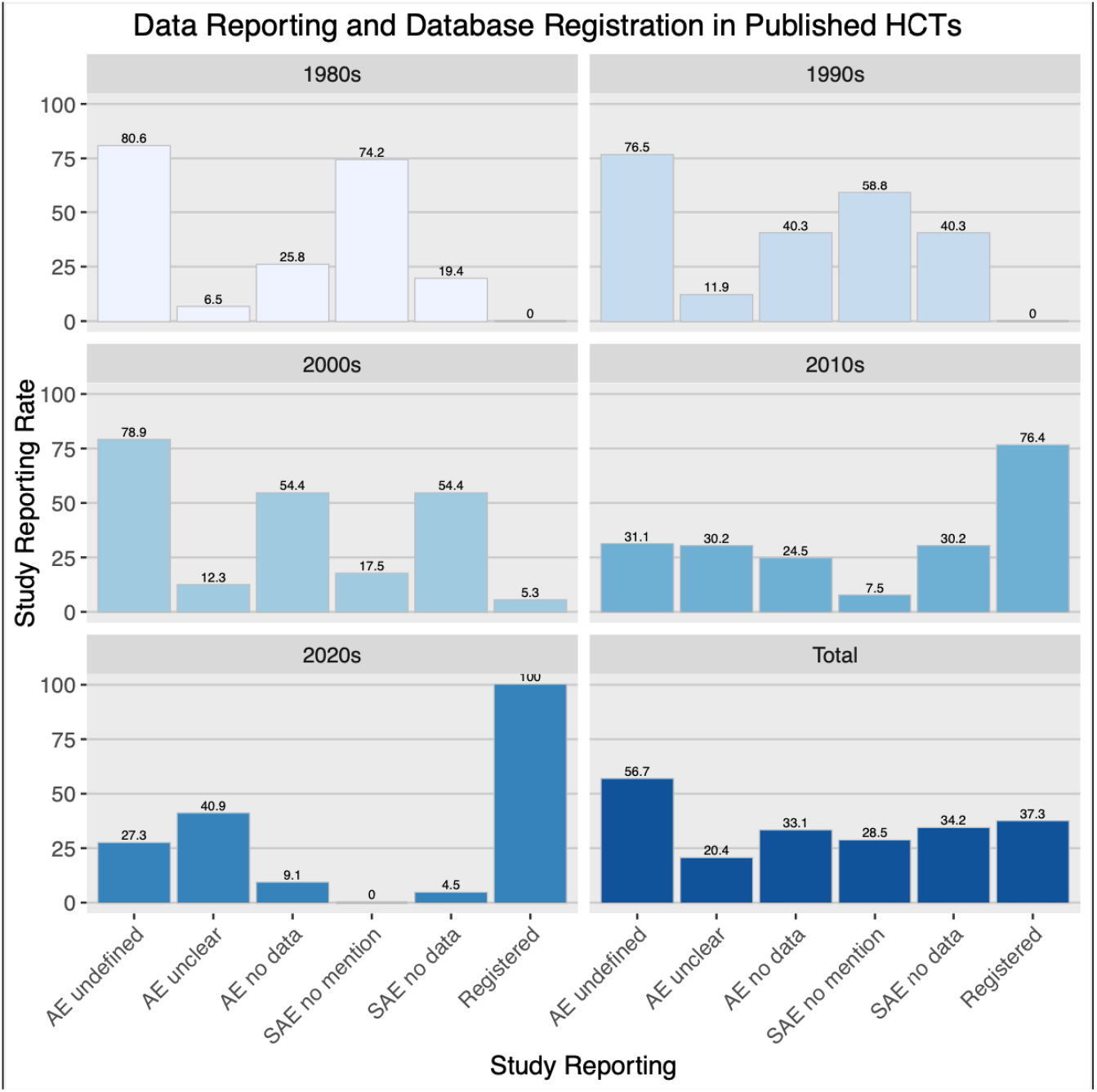

Among 10,325 challenged participants in studies that reported AEs, between 4,317 (41.8%) and 5,730 (55.5%) experienced at least one AE (Table 4). Among 5,083 challenged participants in studies that graded severity of AEs, between 285 (5.6%) and 801 (15.8%) experienced at least one severe or very severe (grade 3 or higher) AE (Table 5). The range in possible AE values is greater in more recent decades as a result of more studies reporting AEs by individual or symptom, rather than reporting the total number of participants with at least one AE. 19 studies included control (non-challenged) participants (n=433); only two of these studies reported AE data for control participants (n=69). Between seven (10.1%) and 12 (17.4%) control participants experienced at least one AE.

**Table 4.** Adverse Events in Published HCTs by Decade.

| Decade | Studies<sup>a</sup>, n | Participants challenged, n | Challenged participants with AEs (minimum<sup>b</sup>), n (%) | Challenged participants with AEs (maximum<sup>b</sup>), n (%) |
| --- | --- | --- | --- | --- |
| **1980s** | 23 | 1448 | 389 (26.9) | 428 (29.6) |
| **1990s** | 41 | 2875 | 1192 (41.5) | 1384 (48.1) |
| **2000s** | 26 | 1984 | 743 (37.4) | 1001 (50.5) |
| **2010s** | 80 | 3139 | 1576 (50.2) | 2210 (70.4) |
| **2020s** | 20 | 879 | 417 (47.4) | 707 (80.4) |
| **Total** | 190 | 10325 | 4317 (41.8) | 5730 (55.5) |

<sup>a</sup>94 studies that did not report AE data are excluded.
<sup>b</sup>Minimum and maximum values are given to account for unclear data reporting by some studies.

**Table 5.** Severe Adverse Events in Published HCTs by Decade.

| Decade | Studies<sup>a</sup>, n | Participants challenged, n | Challenged participants with severe or very severe (≥grade 3) AEs (minimum<sup>b</sup>), n (%) | Challenged participants with severe or very severe (≥grade 3) AEs (maximum<sup>b</sup>), n (%) |
| --- | --- | --- | --- | --- |
| **1980s** | 3 | 77 | 9 (11.7) | 25 (32.5) |
| **1990s** | 8 | 429 | 23 (5.4) | 23 (5.4) |
| **2000s** | 12 | 1984 | 31 (1.6) | 102 (5.1) |
| **2010s** | 57 | 1954 | 179 (9.2) | 473 (24.2) |
| **2020s** | 14 | 639 | 43 (6.7) | 178 (27.9) |
| **Total** | 94 | 5083 | 285 (5.6) | 801 (15.8) |

<sup>a</sup>190 studies that did not report severe AE data are excluded.
<sup>b</sup>Minimum and maximum values are given to account for unclear data reporting by some studies.

Among 10,016 challenged participants in studies that reported SAEs, 23 (0.2%) experienced at least one SAE (Table 6). Among 146 rechallenged participants in studies that reported SAEs, one additional participant (0.7%) experienced at least one SAE (Supplementary Table 6). No fatalities were reported. SAEs are described in more detail in Table 7, and some SAEs deemed not related to challenge are discussed further in Supplementary Table 7.

**Table 6.** Serious Adverse Events in Published HCTs by Decade.

| **Decade** | **Studies<sup>a</sup>, n** | **Participants challenged, n** | **Challenged participants with SAEs, n (%)** |
| --- | --- | --- | --- |
| **1980s** | 25 | 1469 | 6 (0.4) |
| **1990s** | 41 | 2799 | 1 (0.0) |
| **2000s** | 26 | 1623 | 1 (0.1) |
| **2010s** | 74 | 3194 | 13 (0.4) |
| **2020s** | 21 | 931 | 2 (0.2) |
| **Total** | 187 | 10016 | 23<sup>b</sup> (0.2) |

<sup>a</sup>97 studies that did not report SAE data are excluded.
<sup>b</sup>Two additional SAEs described in Table 7 (one in a rechallenge, one reported in a case report) are not included in this total.

**Table 7.**
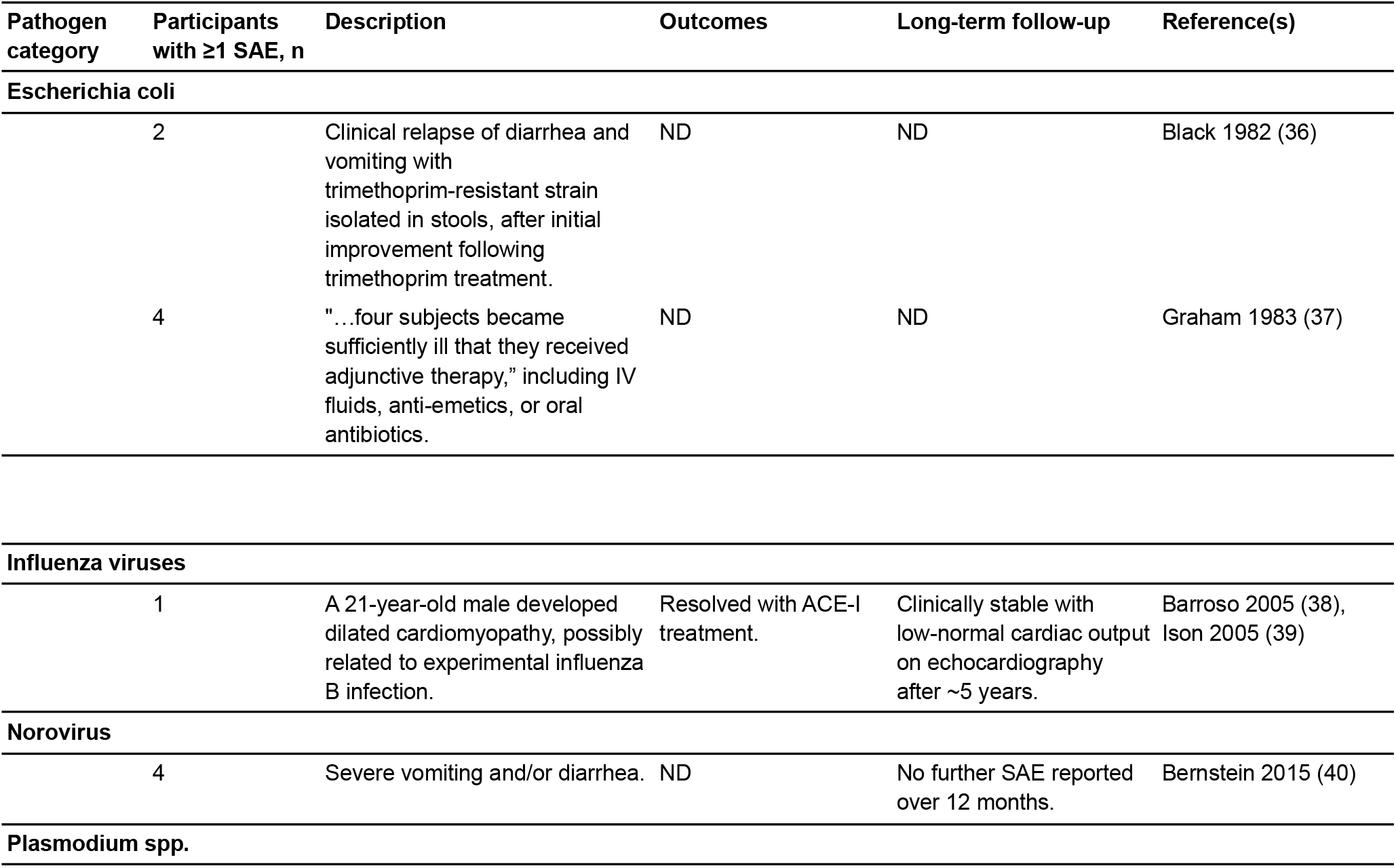
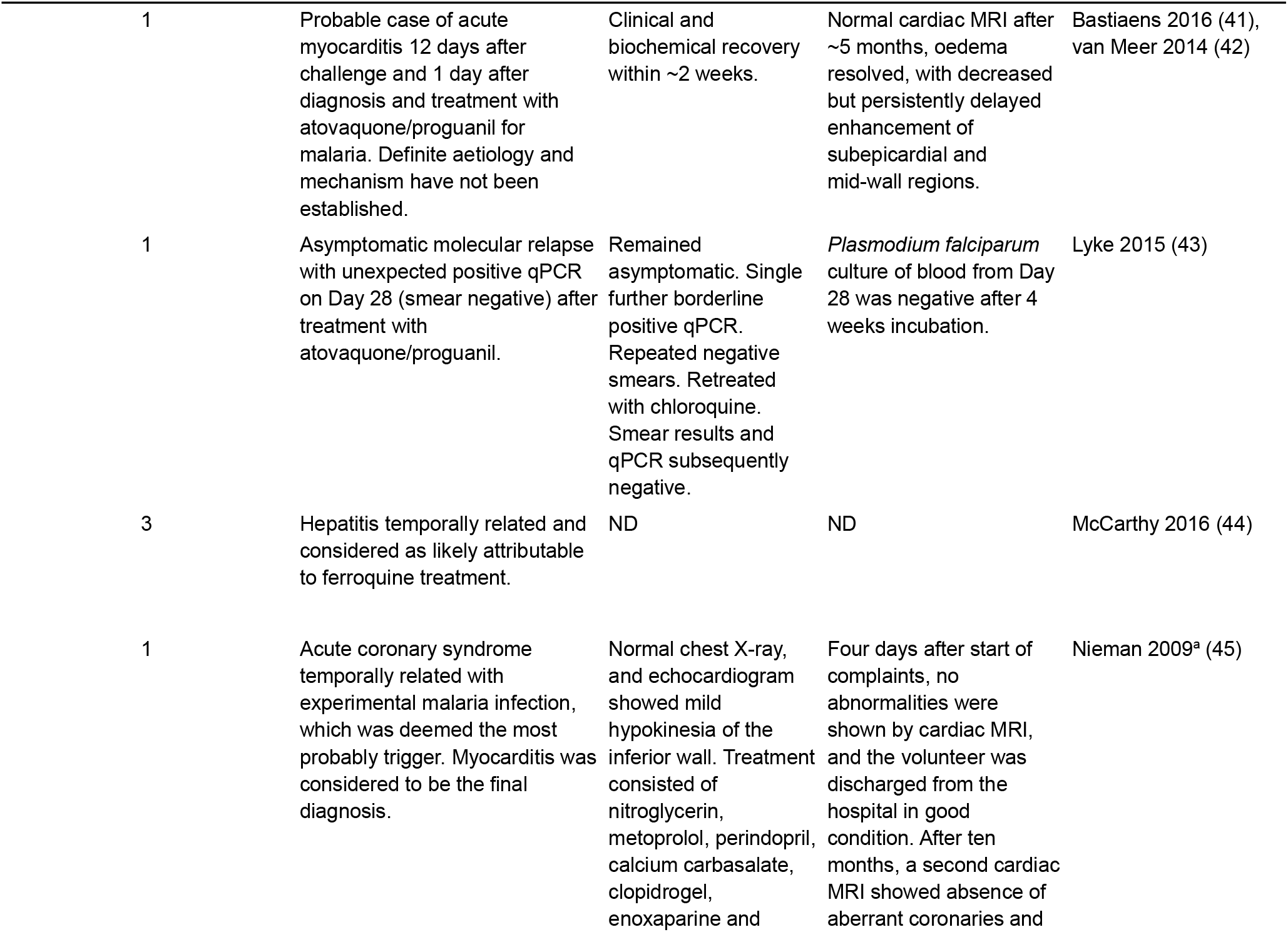
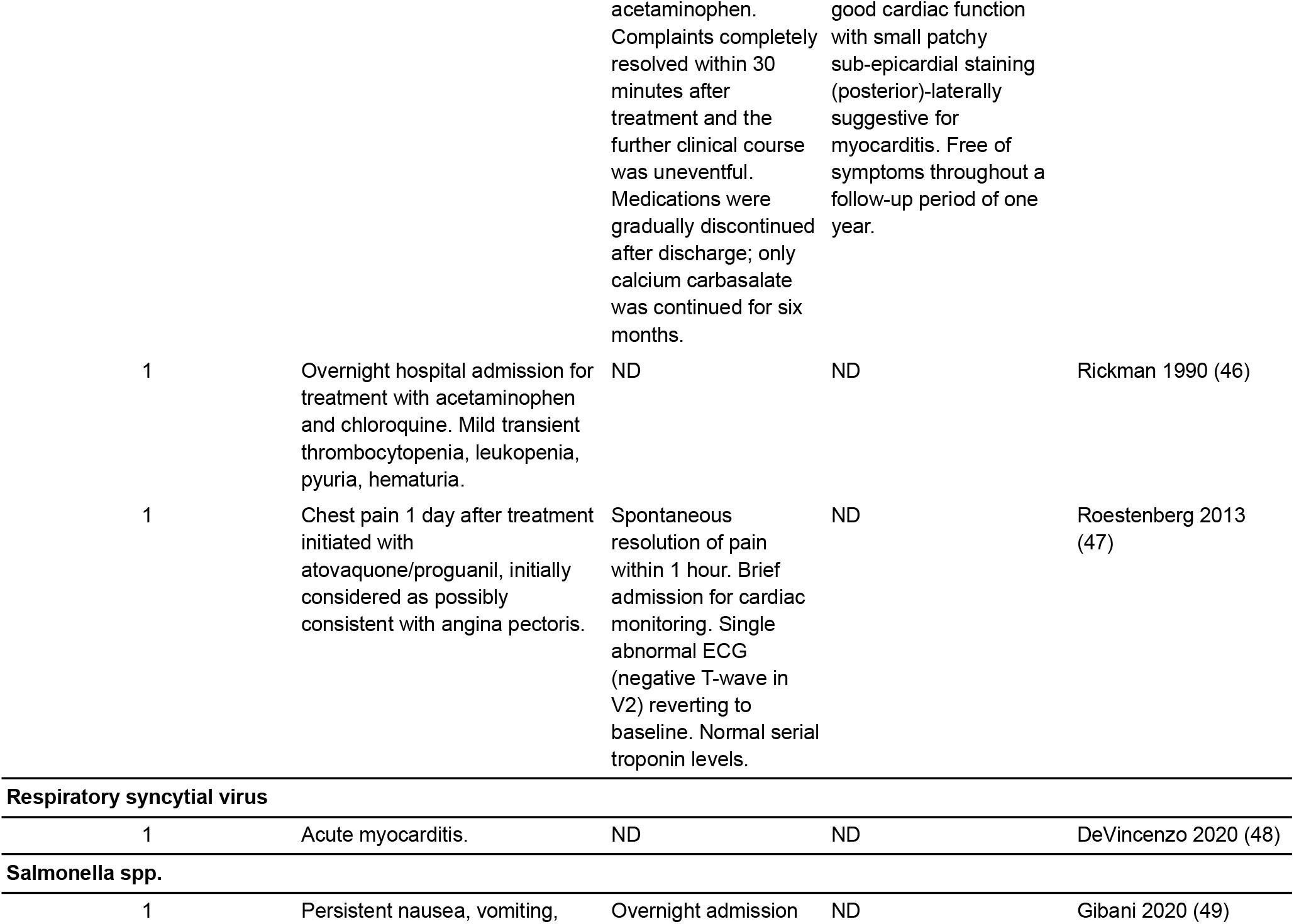
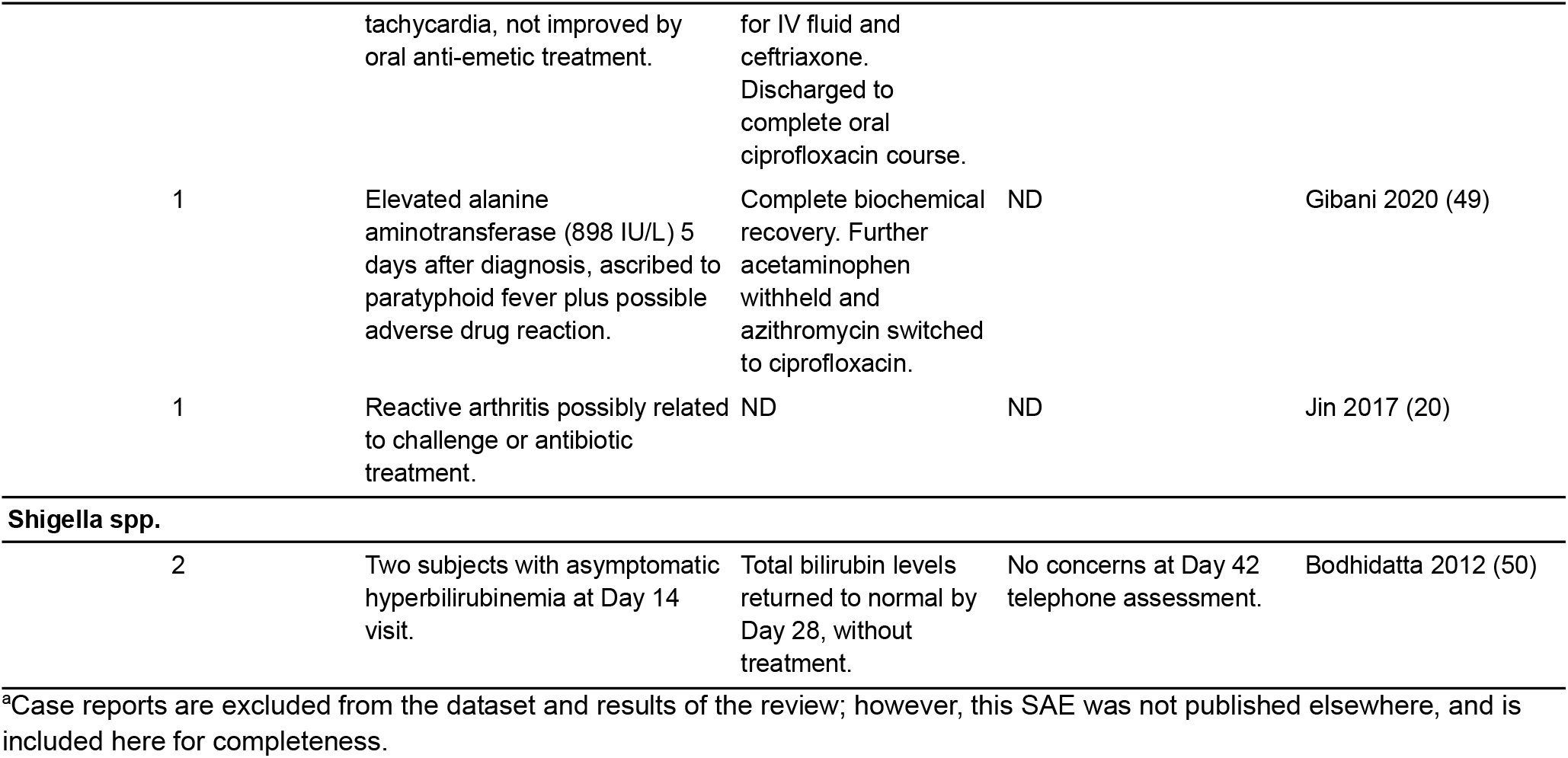
Descriptions of Serious Adverse Events in Published HCTs by Pathogen Category.

### 3.4 Studies by Pathogen

The numbers of studies and participants challenged within each category of pathogen are presented in Table 8, and Figure 2a illustrates studies of different pathogens have occurred over time. There were 28 pathogen categories, with the most commonly studied being *Plasmodium* spp. (73 studies, 1,689 participants), influenza viruses (45 studies, 3,536 participants), and rhinovirus (43 studies, 4,332 participants). Studies investigating *Plasmodium* spp. had the greatest number of challenged participants with SAEs, with seven SAEs (out of 23 in all non-rechallenge studies) occurring among 1,129 participants in 52 studies. Studies investigating norovirus had the greatest proportion of SAEs to number challenged, with four SAEs occurring among 163 participants in three studies.

**Table 8.**
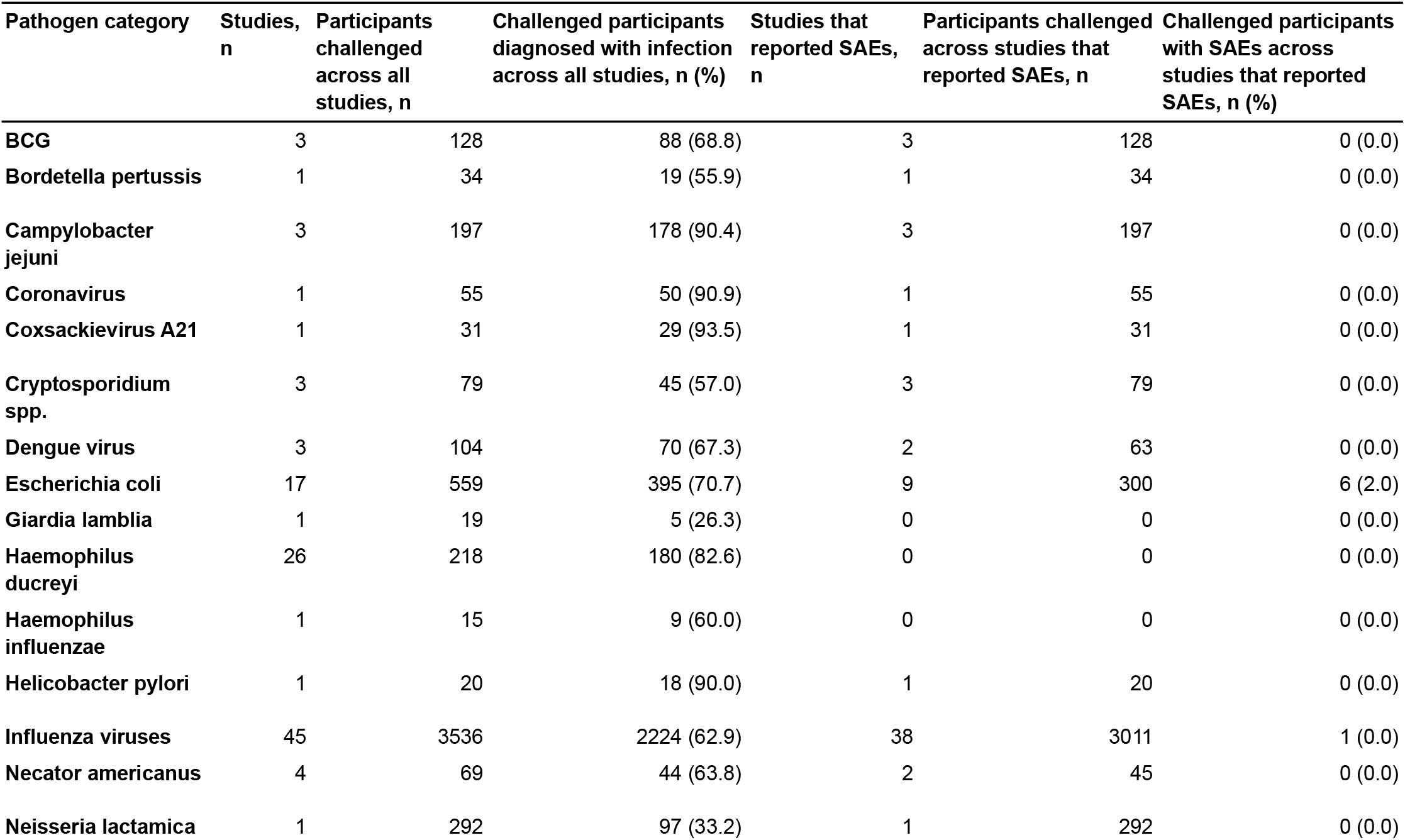
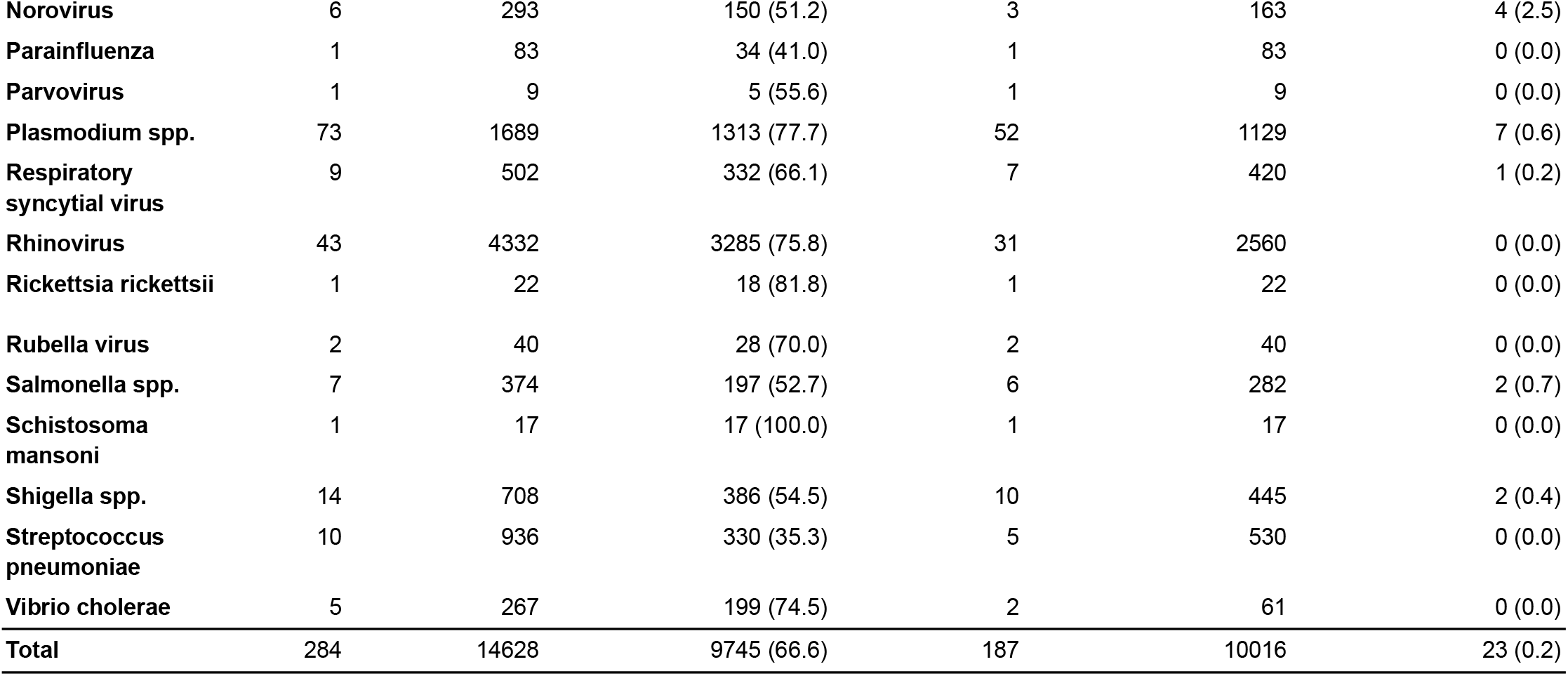
Number of Published HCTS, Number of Participants, Number Infected, and Number with Serious Adverse Events by Pathogen Category.

**Figure 2.**
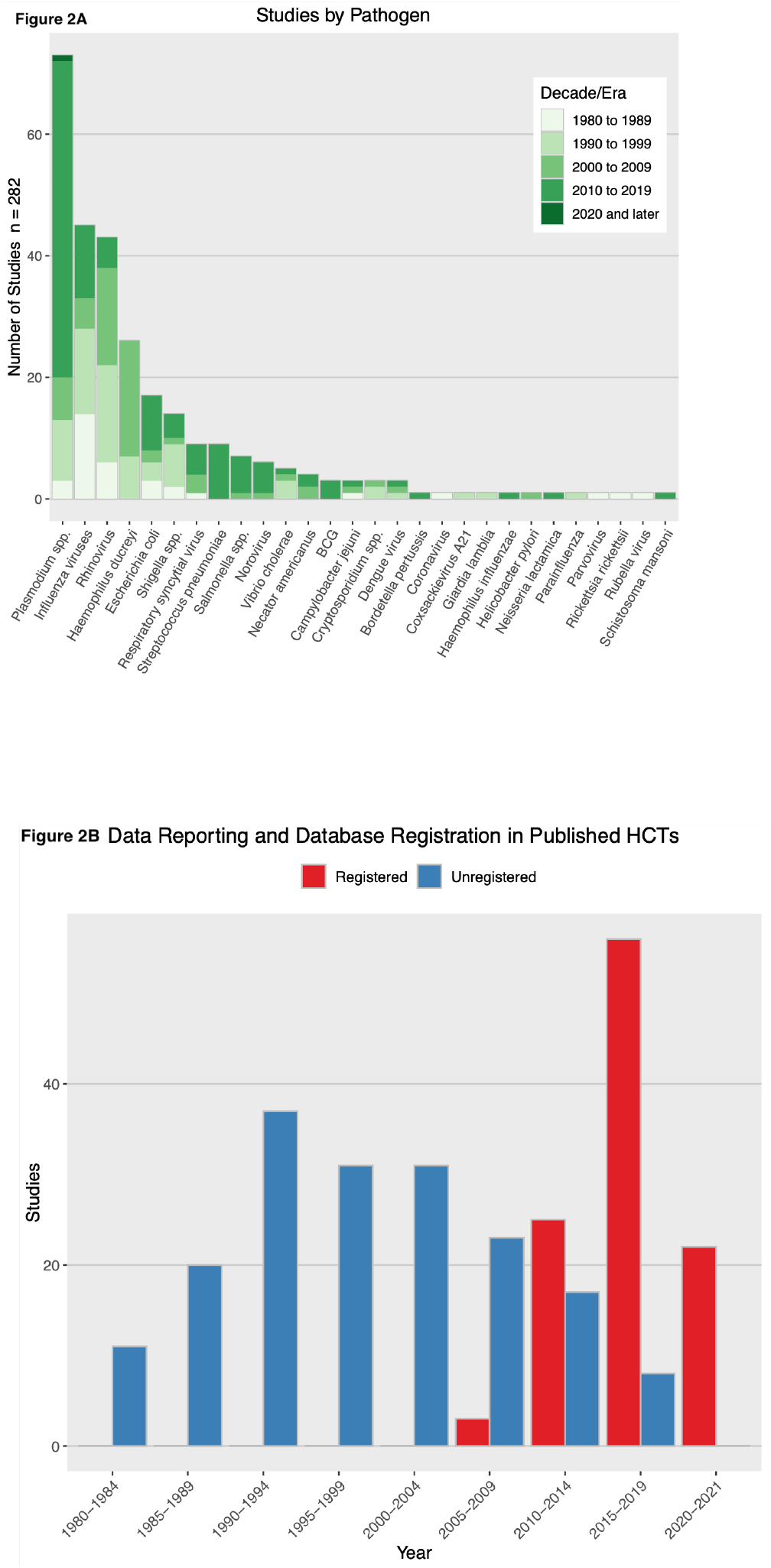

### 3.5 Reporting Adverse Events and Use of Trial Registries Over Time

Overall, the number of challenge studies has been increasing each decade (Figure 2b).

Prior to the 2000s, many studies did not report AEs, but instead reported comparable symptom data. These were extracted as AEs. Of the 283 included studies, 123 explicitly mentioned or defined AEs, but not all reported them for the challenge phase specifically. The proportion of studies with definitions has increased over time, from only 19.4%, 23.9%, and 21.1% in the 1980s, 1990s, and 2000s respectively, to 68.9% and 72.7% in the 2010s and 2020s (thus far) respectively. Results that exclude studies that did not explicitly mention AEs and SAEs are presented in Supplementary Tables 9 and 10.

The National Institutes of Health (NIH) launched ClinicalTrials.gov on February 29, 2000. For NIH-funded research, post-2007, “applicable clinical trials” are required to be registered (51). However, publication year lags year of registration, so it is unclear how much of the lack of registration is noncompliance and how much is delayed publication. Still, only 5.3% of included studies published in the 2000s were registered in at least one registry; 76.4% of included studies published in the 2010s were registered in at least one registry. Every included study published so far this decade was registered.

### 3.6 Risk Mitigation

Text describing specific risk mitigation measures was found in 286 of the 308 studies, which is included in the dataset (https://github.com/1DaySooner/HCTSystematicReview), and a descriptive summary follows. The qualitative nature of these mitigation descriptions precluded meaningful quantitative analysis.

Risk mitigation measures typically include evaluating participants’ risk of disease if exposed to a challenge agent, by using medical screening and assessing participants’ medical histories. In some cases, checking for prior exposure to the pathogen was a risk mitigation strategy, but it could also be done for other reasons. Demographic criteria, pregnancy screening, assessment of cardiac risk, and assessment of weight and/or BMI were often used to evaluate risk. Many studies reported using treatment for the challenge infection where relevant (“rescue therapies”) to ensure that volunteers were cleared of infection before discharge. Many studies reported evaluating participants’ suitability for these therapies prior to challenge.

Some studies reported mitigation strategies for risks to non-participants, such as isolation throughout the duration of the study, requiring birth control, or excluding participants with employment posing risk of spread (for example, excluding food handlers in HCTs investigating *Escherichia coli*, norovirus, and *Salmonella spp*.). Validity of informed consent was sometimes assessed by testing participants’ understanding of the study protocol.

## 4 Discussion

The present review found a total of 24 (23 reported in traditional challenges, one in a rechallenge) SAEs and zero reported deaths or cases of permanent damage among 15,046 participants in 308 studies spanning 1980 to 2021. It is unlikely that any SAEs captured in this review (Table 7) were life-threatening, as they were primarily categorized as SAEs due to involving brief hospitalization for observation or supportive care, requiring non-invasive interventions (such as re-treatment for relapses), or falling under the broad category of “other serious (important medical events)” in the FDA definition of SAEs. The proportions of studies that define AEs and mention SAEs have increased over time, although inconsistent definitions make it challenging to compare reported data, particularly across studies investigating different pathogens. Unfortunately, the proportions of studies that don’t report AE and SAE data related to challenges remained unacceptably high in the 2010s at 24.5% and 30.2%, respectively (Table 3). While a high rate of failing to report SAEs may be indicative of their rarity in the HCT setting, clearer reporting would allow for better understanding of the risks and benefits of HCTs.

Issues surrounding AE reporting in clinical trials are not exclusive to HCTs (52). However, confusion related to reporting challenge-related AEs is an issue specific to HCTs. For example, some studies identified “expected symptoms” as being distinct from AEs, only reported AEs related to interventions, or omitted discussion of AEs entirely. Additionally, clinical endpoints (such as moderate to severe diarrhea in *E. coli* HCTs) were not always reported as AEs by the study. There is a greater degree of consistency for SAE reporting (generally in agreement with the FDA definition (26)), but many studies, especially those published prior to 2000, did not define or report SAEs. Guidelines for HCT reporting have been suggested (2), but have not yet been adopted. Accordingly, a major conclusion of this review is that in addition to a greater effort to standardize AE reporting in general, which others have postulated (53,54), these standardization efforts are particularly valuable to HCTs.

The number of new HCTs has been increasing; however, it is unclear whether this increase is proportional to the general growth trend in the number of new (non-HCT) clinical trials. Since 2010, pathogens such as *Bordetella pertussis, Schistosoma mansoni*, and *Streptococcus pneumoniae* have been studied in HCTs for the first time. Figure 2a shows that the number of influenza and rhinovirus HCTs has declined somewhat over time, following the discontinuation of several research programs focused on common cold, while the number of *Plasmodium* spp. HCTs sharply increased in the 2010s. These trends demonstrate that HCTs are an increasingly ubiquitous tool, and their relative speed allows researchers to investigate new pathogens of interest more rapidly than in traditional clinical trials.

Limitations of this review are primarily related to uncertainties around the accuracy of AE reporting. This includes potential bias in AE reporting, inconsistent reporting, and difficulty in precisely estimating the rates of events based on provided data. Many studies reported either no or unclear AE and/or SAE data, and issues of censoring and misclassification are common with respect to AE reporting in general (53). To partially address issues with different standards for reporting over time, we extracted symptom data as AE and/or SAE data from studies that did not mention or define AEs/SAEs, but this means that AEs for decades in which these studies occurred are not fully comparable. The review is further limited by our inability to locate some results, including published HCTs that were not on PubMed (55) and HCTs whose results have only been published as case reports (45). These limitations further highlight the need for improvements in the field of HCTs with respect to AE reporting and availability of results. Future work building off of this review includes policy recommendations around the issues of standardization and AE reporting, investigating the registration of HCTs in databases, and further qualitative analysis of risk mitigation measures in published articles.

## 5 Conclusions

The recent literature contains hundreds of HCTs involving over 10,000 participants and only 24 SAEs, with no recorded deaths or cases of permanent health damage. HCTs are now routinely used to understand infectious dose, disease progression, clinical efficacy of novel interventions, and immune response for a wide variety of pathogens. As evidenced by recent HCTs for COVID-19, they may be conducted for novel as well as familiar diseases. This review can help support public discussion and expert deliberation regarding the safety of HCTs. It may also inform future discussions among HCT researchers and members of ethics review committees regarding the planning, conduct, and reporting of future HCTs.

### Preregistration, Protocol, and Conflict of Interest Disclosures

The review was preregistered on PROSPERO as CRD42021247218, risk outcomes and risk mitigation measures in human challenge trials: a systematic review. The review protocol is included as supplementary materials. As mentioned above, the preregistration was amended to include additional searches and data.

Thank you to 1DaySooner for supporting this work, and to the 1DaySooner Scientific Advisory board, which includes coauthors DM, VS, and WW, for reviewing the proposed study.

## Supporting information

Review workflow

## Data Availability

All data is publicly available online at https://github.com/1DaySooner/HCTSystematicReview

## Contribution Statement

DM, WW, and VS conceived of the idea. EJ, JO, MR, and WW provided initial feedback and refined the idea. DT and DM designed and preregistered the systematic review, with feedback and expert guidance from EJ, JO, and MR. DT led the initial review for inclusion, with JAP and KS. The full text reviews were done by JAP, DT, and two non-author reviewers thanked below: SK and DK. Disputes were resolved by DM. Guidance on inclusion criteria and interpretation was provided by EJ, KS, and JO. JAP and DT led writing of the manuscript, with supervision and assistance by DM, VS, WW, KS, EJ, and JO. JW managed the dataset, performed analysis, and produced data summaries and visualizations.

## Acknowledgements

We would like to thank Steffen Kamenicek and Daniel Kaufman, who assisted with full text review of papers and data extraction. We would also like to thank Steffen Kamenicek and River Bellamy for additional assistance with reviewing the text, tables, and figures prior to submission.

## Availability of Data, Code, and Other Materials

The complete dataset of included studies is available at https://github.com/1DaySooner/HCTSystematicReview.

## 6 References

1. Gordon SB, Rylance J, Luck A, Jambo KC, Ferreira DM, Manda-Taylor L, et al. A framework for Controlled Human Infection Model (CHIM) studies in Malawi: Report of a Wellcome Trust workshop on CHIM in Low Income Countries held in Blantyre, Malawi. Wellcome Open Res. 2017 Aug 24;2:70.

2. Kalil JA, Halperin SA, Langley JM. Human challenge studies: a review of adequacy of reporting methods and results. Future Microbiol. 2012 Apr 1;7(4):481–95.

3. Roestenberg M, Hoogerwerf M-A, Ferreira DM, Mordmüller B, Yazdanbakhsh M. Experimental infection of human volunteers. Lancet Infect Dis. 2018;18(10):e312–22.

4. Jamrozik E, Selgelid MJ. Human challenge studies in endemic settings: ethical and regulatory issues. Springer Nature; 2021. 134 p. (SpringerBriefs in Ethics).

5. Sherman AC, Mehta A, Dickert NW, Anderson EJ, Rouphael N. The Future of Flu: A Review of the Human Challenge Model and Systems Biology for Advancement of Influenza Vaccinology. Front Cell Infect Microbiol [Internet]. 2019 Apr 17 [cited 2021 May 15];9. Available from: https://www.ncbi.nlm.nih.gov/pmc/articles/PMC6489464/

6. Nielsen CM, Vekemans J, Lievens M, Kester KE, Regules JA, Ockenhouse CF. RTS,S malaria vaccine efficacy and immunogenicity during Plasmodium falciparum challenge is associated with HLA genotype. Vaccine. 2018 Mar 14;36(12):1637–42.

7. Mosley JF, Smith LL, Brantley P, Locke D, Como M. Vaxchora: The First FDA-Approved Cholera Vaccination in the United States. Pharm Ther. 2017 Oct;42(10):638–40.

8. Feasey NA, Levine MM. Typhoid vaccine development with a human challenge model. The Lancet. 2017 Dec 2;390(10111):2419–21.

9. Palacios R, Shah SK. When could human challenge trials be deployed to combat emerging infectious diseases? Lessons from the case of a Zika virus human challenge trial. Trials. 2019 Dec 19;20(2):702.

10. Nguyen LC, Bakerlee CW, McKelvey TG, Rose SM, Norman AJ, Joseph N, et al. Evaluating Use Cases for Human Challenge Trials in Accelerating SARS-CoV-2 Vaccine Development. Clin Infect Dis [Internet]. 2020 Jul; Available from: https://academic.oup.com/cid/advance-article/doi/10.1093/cid/ciaa935/5868014

11. Pandemic Preparedness Partnership. 100 days mission to respond to future pandemic threats [Internet]. 2021 Jun. Available from: https://www.gov.uk/government/publications/100-days-mission-to-respond-to-future-pandemic-threats

12. Chulay JD, Schneider I, Cosgriff TM, Hoffman SL, Ballou WR, Quakyi IA, et al. Malaria Transmitted to Humans by Mosquitoes Infected from Cultured Plasmodium falciparum. Am J Trop Med Hyg. 1986 Jan 1;35(1):66–8.

13. Cate TR, Couch RB. Live influenza A/Victoria/75 (H3N2) virus vaccines: reactogenicity, immunogenicity, and protection against wild-type virus challenge. Infect Immun. 1982 Oct 1;38(1):141–6.

14. Turner RB, Winther B, Hendley JO, Myglnd N, Gwaltney JM. Sites of Virus Recovery and Antigen Detection in Epithelial Cells during Experimental Rhinovirus Infection. Acta Otolaryngol (Stockh). 1984 Jan 1;98(sup417):9–14.

15. Herrington DA, Van De Verg L, Formal SB, Hale TL, Tall BD, Cryz SJ, et al. Studies in volunteers to evaluate candidate Shigella vaccines: further experience with a bivalent Salmonella typhi-Shigella sonnei vaccine and protection conferred by previous Shigella sonnei disease. Vaccine. 1990 Aug 1;8(4):353–7.

16. Tacket CO, Forrest B, Morona R, Attridge SR, LaBrooy J, Tall BD, et al. Safety, immunogenicity, and efficacy against cholera challenge in humans of a typhoid-cholera hybrid vaccine derived from Salmonella typhi Ty21a. Infect Immun. 1990 Jun 1;58(6):1620–7.

17. Memoli MJ, Shaw PA, Han A, Czajkowski L, Reed S, Athota R, et al. Evaluation of Antihemagglutinin and Antineuraminidase Antibodies as Correlates of Protection in an Influenza A/H1N1 Virus Healthy Human Challenge Model. mBio. 2016 Apr 19;7(2):e00417–00416.

18. Cabrera A, Lepage JE, Sullivan KM, Seed SM. Vaxchora: A Single-Dose Oral Cholera Vaccine. Ann Pharmacother. 2017 Jul;51(7):584–9.

19. Hayden FG, Treanor JJ, Fritz RS, Lobo M, Betts RF, Miller M, et al. Use of the oral neuraminidase inhibitor oseltamivir in experimental human influenza: randomized controlled trials for prevention and treatment. JAMA. 1999 Oct 6;282(13):1240–6.

20. Jin C, Gibani MM, Moore M, Juel HB, Jones E, Meiring J, et al. Efficacy and immunogenicity of a Vi-tetanus toxoid conjugate vaccine in the prevention of typhoid fever using a controlled human infection model of Salmonella Typhi: a randomised controlled, phase 2b trial. The Lancet. 2017 Dec 2;390(10111):2472–80.

21. Regules JA, Cicatelli SB, Bennett JW, Paolino KM, Twomey PS, Moon JE, et al. Fractional Third and Fourth Dose of RTS,S/AS01 Malaria Candidate Vaccine: A Phase 2a Controlled Human Malaria Parasite Infection and Immunogenicity Study. J Infect Dis. 2016 Sep 1;214(5):762–71.

22. Hoogerwerf M-A, Vries M de, Roestenberg M. Money-oriented risk-takers or deliberate decision-makers: a cross-sectional survey study of participants in controlled human infection trials. BMJ Open. 2020 Jul 1;10(7):e033796.

23. Lynch HF, Darton TC, Levy J, McCormick F, Ogbogu U, Payne RO, et al. Promoting Ethical Payment in Human Infection Challenge Studies. Am J Bioeth. 2021 Mar 4;21(3):11–31.

24. Hope T, McMillan J. Challenge studies of human volunteers: ethical issues. J Med Ethics. 2004 Feb 1;30(1):110.

25. Grimwade O, Savulescu J, Giubilini A, Oakley J, Osowicki J, Pollard AJ, et al. Payment in challenge studies: ethics, attitudes and a new payment for risk model. J Med Ethics. 2020;

26. Investigational New Drug Application. Code of Federal Regulations. Sect. 312.32 IND safety reporting.

27. Guidance for Clinical Investigators, Sponsors, and IRBs; Investigational New Drug Applications (INDs) — Determining Whether Human Research Studies Can Be Conducted Without an IND [Internet]. U.S. Department of Health and Human Services, Food and Drug Administration, Center for Drug Evaluation and Research (CDER), Center for Biologics Evaluation and Research (CBER), and Center for Food Safety and Applied Nutrition (CFSAN); 2013 Sep. Available from: https://www.fda.gov/files/drugs/published/Investigational-New-Drug-Applications-%28INDs%29-Determining-Whether-Human-Research-Studies-Can-Be-Conducted-Without-an-IND.pdf

28. Manheim D, Toomey D, Wiecek W, Schmit V, Adams-Phipps J, Scholl K. Risk outcomes and risk mitigation measures in human challenge trials: a systematic review. [Internet]. PROSPERO: International prospective register of systematic reviews; 2021. Available from: https://www.crd.york.ac.uk/prospero/display_record.php?ID=CRD42021247218

29. Pollard AJ, Sauerwein R, Baay M, Neels P, Balasingam S, Bejon P, et al. Third human challenge trial conference, Oxford, United Kingdom, February 6–7, 2020, a meeting report. Biologicals. 2020 Jul 1;66:41–52.

30. Haddaway NR, McGuinness LA, Pritchard CC. PRISMA2020: R package and ShinyApp for producing PRISMA 2020 compliant flow diagrams [Internet]. Zenodo; 2021 [cited 2022 Feb 8]. Available from: https://zenodo.org/record/5082518

31. Atmar RL, Opekun AR, Gilger MA, Estes MK, Crawford SE, Neill FH, et al. Determination of the 50% human infectious dose for Norwalk virus. J Infect Dis.2014 Apr 1;209(7):1016–22.

32. Atmar RL, Opekun AR, Gilger MA, Estes MK, Crawford SE, Neill FH, et al. Norwalk virus shedding after experimental human infection. Emerg Infect Dis. 2008 Oct;14(10):1553–7.

33. Nguyen-Van-Tam JS, Killingley B, Enstone J, Hewitt M, Pantelic J, Grantham ML, et al. Minimal transmission in an influenza A (H3N2) human challenge-transmission model within a controlled exposure environment. PLOS Pathog. 2020 Jul 13;16(7):e1008704.

34. Hickey BW, Lumsden JM, Reyes S, Sedegah M, Hollingdale MR, Freilich DA, et al. Mosquito bite immunization with radiation-attenuated Plasmodium falciparum sporozoites: safety, tolerability, protective efficacy and humoral immunogenicity. Malar J. 2016 Jul 22;15(1):377.

35. Chen WH, Cohen MB, Kirkpatrick BD, Brady RC, Galloway D, Gurwith M, et al. Single-dose Live Oral Cholera Vaccine CVD 103-HgR Protects Against Human Experimental Infection With Vibrio cholerae O1 El Tor. Clin Infect Dis. 2016 Jun 1;62(11):1329–35.

36. Black RE, Levine MM, Clements ML, Cisneros L, Daya V. Treatment of Experimentally Induced Enterotoxigenic Escherichia coli Diarrhea with Trimethoprim, Trimethoprim-Sulfamethoxazole, or Placebo. Rev Infect Dis. 1982 Mar 1;4(2):540–5.

37. Graham DY, Estes MK, Gentry LO. Double-blind comparison of bismuth subsalicylate and placebo in the prevention and treatment of enterotoxigenic Escherichia coli-induced diarrhea in volunteers. Gastroenterology. 1983 Nov;85(5):1017–22.

38. Barroso L, Treanor J, Gubareva L, Hayden FG. Efficacy and Tolerability of the Oral Neuraminidase Inhibitor Peramivir in Experimental Human Influenza: Randomized, Controlled Trials for Prophylaxis and Treatment. Antivir Ther. 2005 Nov 1;10(8):901–10.

39. Ison MG, Campbell V, Rembold C, Dent J, Hayden FG. Cardiac Findings during Uncomplicated Acute Influenza in Ambulatory Adults. Clin Infect Dis. 2005 Feb 1;40(3):415–22.

40. Bernstein DI, Atmar RL, Lyon GM, Treanor JJ, Chen WH, Jiang X, et al. Norovirus Vaccine Against Experimental Human GII.4 Virus Illness: A Challenge Study in Healthy Adults. J Infect Dis. 2015 Mar 15;211(6):870–8.

41. Bastiaens GJH, van Meer MPA, Scholzen A, Obiero JM, Vatanshenassan M, van Grinsven T, et al. Safety, Immunogenicity, and Protective Efficacy of Intradermal Immunization with Aseptic, Purified, Cryopreserved Plasmodium falciparum Sporozoites in Volunteers Under Chloroquine Prophylaxis: A Randomized Controlled Trial. Am J Trop Med Hyg. 2016 Mar;94(3):663–73.

42. van Meer MP, Bastiaens GJ, Boulaksil M, de Mast Q, Gunasekera A, Hoffman SL, et al. Idiopathic acute myocarditis during treatment for controlled human malaria infection: a case report. Malar J. 2014 Jan 30;13(1):38.

43. Lyke KE, Laurens MB, Strauss K, Adams M, Billingsley PF, James E, et al. Optimizing Intradermal Administration of Cryopreserved Plasmodium falciparum Sporozoites in Controlled Human Malaria Infection. Am J Trop Med Hyg. 2015 Dec;93(6):1274–84.

44. McCarthy JS, Rückle T, Djeriou E, Cantalloube C, Ter-Minassian D, Baker M, et al. A Phase II pilot trial to evaluate safety and efficacy of ferroquine against early Plasmodium falciparum in an induced blood-stage malaria infection study. Malar J. 2016 Sep 13;15:469.

45. Nieman A-E, de Mast Q, Roestenberg M, Wiersma J, Pop G, Stalenhoef A, et al. Cardiac complication after experimental human malaria infection: a case report. Malar J. 2009 Dec 3;8(1):277.

46. Rickman LS, Jones TR, Long GW, Paparello S, Schneider I, Paul CF, et al. Plasmodium falciparum-infected Anopheles stephensi inconsistently transmit malaria to humans. Am J Trop Med Hyg. 1990 Nov;43(5):441–5.

47. Roestenberg M, Bijker EM, Sim BKL, Billingsley PF, James ER, Bastiaens GJH, et al. Controlled human malaria infections by intradermal injection of cryopreserved Plasmodium falciparum sporozoites. Am J Trop Med Hyg. 2013 Jan;88(1):5–13.

48. DeVincenzo J, Tait D, Efthimiou J, Mori J, Kim Y-I, Thomas E, et al. A Randomized, Placebo-Controlled, Respiratory Syncytial Virus Human Challenge Study of the Antiviral Efficacy, Safety, and Pharmacokinetics of RV521, an Inhibitor of the RSV-F Protein. Antimicrob Agents Chemother. 2020 Jan 27;64(2):e01884–19.

49. Gibani MM, Jin C, Shrestha S, Moore M, Norman L, Voysey M, et al. Homologous and heterologous re-challenge with Salmonella Typhi and Salmonella Paratyphi A in a randomised controlled human infection model. PLoS Negl Trop Dis. 2020 Oct;14(10):e0008783.

50. Bodhidatta L, Pitisuttithum P, Chamnanchanant S, Chang KT, Islam D, Bussaratid V, et al. Establishment of a Shigella sonnei human challenge model in Thailand. Vaccine. 2012 Nov 19;30(49):7040–5.

51. Clinical Trials Registration And Results Information Submission [Internet]. US Code of Federal Regulations, Title 42(I)(A) Part 11 2016. Available from: https://www.ecfr.gov/current/title-42/chapter-I/subchapter-A/part-11

52. Ann Neuer. A Fresh Take on the Adverse Event Landscape. Clin Res [Internet]. 2019 Feb 12 [cited 2021 Nov 16];33(2). Available from: https://acrpnet.org/2019/02/12/a-fresh-take-on-the-adverse-event-landscape/

53. Schroll JB, Maund E, Gøtzsche PC. Challenges in Coding Adverse Events in Clinical Trials: A Systematic Review. PLOS ONE. 2012 Jul 20;7(7):e41174.

54. Zorzela L, Golder S, Liu Y, Pilkington K, Hartling L, Joffe A, et al. Quality of reporting in systematic reviews of adverse events: systematic review. BMJ. 2014 Jan 8;348:f7668.

55. Osowicki J, Azzopardi KI, Fabri L, Frost HR, Rivera-Hernandez T, Neeland MR, et al. A controlled human infection model of Streptococcus pyogenes pharyngitis (CHIVAS-M75): an observational, dose-finding study. Lancet Microbe. 2021 Jul 1;2(7):e291–9.

