## Supplemental Material for "A Systematic Review of Human Challenge Trials, Designs, and Safety"

### Supplementary Materials

#### Appendix A - Search Strategy Rationale

Five separate searches were performed, as shown in the paper. Search 1 was performed initially and subsequent searches 2-4 were performed to fill gaps left by search 1. After reviewing results, search 5 was performed to identify more papers that were otherwise missed. The search 1 algorithm was repeated in the PubMed and PubMed Central (PMC) databases, where it returned 24 and 417 results respectively. Because of this difference in hit rate, the PMC results were selected for screening.  However, only 27 of these results were published before 2010, leaving our dataset skewed towards one decade.

To address this skew, publications of known challenge trials that should have been included in our search results were used to identify terms that were standard usage in different decades. PubMed searches for MeSH terms in the title and abstract only, while PMC searches for MeSH terms throughout entire documents. As a result, PubMed results are pre-screened by title and abstract and usually have a lower hit count than PMC results. This is preferable in a systematic review workflow as long as PubMed's pre-screening doesn't produce too few results. Searches 2-4 cumulatively produced 899 results on PubMed and 17,593 results on PMC. Because 899 is an appropriate number of results to expect for this search, the PubMed results were screened instead of the PMC results. Likewise, search 5 was performed on PubMed, and produced 1,338 results.

#### Appendix B - Full Citation List of Studies

Note that the dataset with both included and excluded studies and all extracted data from included studies is linked in Appendix E.


1. Kirkpatrick BD, Whitehead SS, Pierce KK, Tibery CM, Grier PL, Hynes NA, et al. The live attenuated dengue vaccine TV003 elicits complete protection against dengue in a human challenge model. Sci Transl Med. 2016 Mar 16;8(330):330ra36.
2. Pitisuttithum P, Cohen MB, Phonrat B, Suthisarnsuntorn U, Bussaratid V, Desakorn V, et al. A human volunteer challenge model using frozen bacteria of the new epidemic serotype, V. cholerae O139 in Thai volunteers. Vaccine. 2001 Dec 12;20(5–6):920–5.

3. Tacket CO, Cohen MB, Wasserman SS, Losonsky G, Livio S, Kotloff K, et al. Randomized, double-blind, placebo-controlled, multicentered trial of the efficacy of a single dose of live oral cholera vaccine CVD 103-HgR in preventing cholera following challenge with Vibrio cholerae O1 El tor inaba three months after vaccination. Infect Immun. 1999 Dec;67(12):6341–5.
4. Sulyok M, Rückle T, Roth A, Mürbeth RE, Chalon S, Kerr N, et al. DSM265 for Plasmodium falciparum chemoprophylaxis: a randomised, double blinded, phase 1 trial with controlled human malaria infection. Lancet Infect Dis. 2017 Jun;17(6):636–44.
5. Feary J, Venn A, Brown A, Hooi D, Falcone FH, Mortimer K, et al. Safety of hookworm infection in individuals with measurable airway responsiveness: a randomized placebo-controlled feasibility study. Clin Exp Allergy. 2009 Jul;39(7):1060–8.
6. Black RE, Levine MM, Clements ML, Cisneros L, Daya V. Treatment of experimentally induced enterotoxigenic Escherichia coli diarrhea with trimethoprim, trimethoprim-sulfamethoxazole, or placebo. Rev Infect Dis. 1982 Apr;4(2):540–5.
7. Turner RB. Ineffectiveness of intranasal zinc gluconate for prevention of experimental rhinovirus colds. Clin Infect Dis. 2001 Dec 1;33(11):1865–70.
8. Turner RB, Bauer R, Woelkart K, Hulsey TC, Gangemi JD. An evaluation of Echinacea angustifolia in experimental rhinovirus infections. N Engl J Med. 2005 Jul 28;353(4):341–8.
9. Turner RB, Wecker MT, Pohl G, Witek TJ, McNally E, St George R, et al. Efficacy of tremacamra, a soluble intercellular adhesion molecule 1, for experimental rhinovirus infection: a randomized clinical trial. JAMA. 1999 May 19;281(19):1797–804.
10. Schiff GM, Young BC, Stefanovic' GM, Stamler EF, Knowlton DR, Grundy BJ, et al. Challenge with rubella virus after loss of detectable vaccine-induced antibody. Rev Infect Dis. 1985 Apr;7 Suppl 1:S157-163.
11. Reichenberg A, Yirmiya R, Schuld A, Kraus T, Haack M, Morag A, et al. Cytokine-associated emotional and cognitive disturbances in humans. Arch Gen Psychiatry. 2001 May;58(5):445–52.
12. Donnenberg MS, Tacket CO, Losonsky G, Frankel G, Nataro JP, Dougan G, et al. Effect of prior experimental human enteropathogenic Escherichia coli infection on illness following homologous and heterologous rechallenge. Infect Immun. 1998 Jan;66(1):52–8.
13. Cate TR, Couch RB. Live influenza A/Victoria/75 (H3N2) virus vaccines: reactogenicity, immunogenicity, and protection against wild-type virus challenge. Infect Immun. 1982 Oct;38(1):141–6.
14. Humphreys TL, Schnizlein-Bick CT, Katz BP, Baldridge LA, Hood AF, Hromas RA, et al. Evolution of the cutaneous immune response to experimental Haemophilus ducreyi infection and its relevance to HIV-1 acquisition. J Immunol. 2002 Dec 1;169(11):6316–23.
15. Bernstein DI, Atmar RL, Lyon GM, Treanor JJ, Chen WH, Jiang X, et al. Norovirus vaccine against experimental human GII.4 virus illness: a challenge study in healthy adults. J Infect Dis. 2015 Mar 15;211(6):870–8.
16. Tacket CO, Binion SB, Bostwick E, Losonsky G, Roy MJ, Edelman R. Efficacy of bovine milk immunoglobulin concentrate in preventing illness after Shigella flexneri challenge. Am J Trop Med Hyg. 1992 Sep;47(3):276–83.

17. Langenberg MCC, Hoogerwerf M-A, Koopman JPR, Janse JJ, Kos-van Oosterhoud J, Feijt C, et al. A controlled human Schistosoma mansoni infection model to advance novel drugs, vaccines and diagnostics. Nat Med. 2020 Mar;26(3):326–32.
18. Hoogerwerf M-A, Koopman JPR, Janse JJ, Langenberg MCC, van Schuijlenburg R, Kruize YCM, et al. A Randomized Controlled Trial to Investigate Safety and Variability of Egg Excretion After Repeated Controlled Human Hookworm Infection. J Infect Dis. 2021 Mar 3;223(5):905–13.
19. Wright AKA, Ferreira DM, Gritzfeld JF, Wright AD, Armitage K, Jambo KC, et al. Human nasal challenge with Streptococcus pneumoniae is immunising in the absence of carriage. PLoS Pathog. 2012;8(4):e1002622.
20. Winokur PL, Chaloner K, Doern GV, Ferreira J, Apicella MA. Safety and immunological outcomes following human inoculation with nontypeable Haemophilus influenzae. J Infect Dis. 2013 Sep 1;208(5):728–38.
21. Wilkinson TM, Li CKF, Chui CSC, Huang AKY, Perkins M, Liebner JC, et al. Preexisting influenza-specific CD4+ T cells correlate with disease protection against influenza challenge in humans. Nat Med. 2012 Jan 29;18(2):274–80.
22. Waddington CS, Darton TC, Jones C, Haworth K, Peters A, John T, et al. An outpatient, ambulant-design, controlled human infection model using escalating doses of Salmonella Typhi challenge delivered in sodium bicarbonate solution. Clin Infect Dis. 2014 May;58(9):1230–40.
23. Tribble DR, Baqar S, Carmolli MP, Porter C, Pierce KK, Sadigh K, et al. Campylobacter jejuni strain CG8421: a refined model for the study of Campylobacteriosis and evaluation of Campylobacter vaccines in human subjects. Clin Infect Dis. 2009 Nov 15;49(10):1512–9.
24. Spring MD, Cummings JF, Ockenhouse CF, Dutta S, Reidler R, Angov E, et al. Phase 1/2a study of the malaria vaccine candidate apical membrane antigen-1 (AMA-1) administered in adjuvant system AS01B or AS02A. PLoS One. 2009;4(4):e5254.
25. Spinola SM, Bong CTH, Faber AL, Fortney KR, Bennett SL, Townsend CA, et al. Differences in host susceptibility to disease progression in the human challenge model of Haemophilus ducreyi infection. Infect Immun. 2003 Nov;71(11):6658–63.
26. Smith CM, Jerkovic A, Truong TT, Foote SJ, McCarthy JS, McMorran BJ. Griseofulvin impairs intraerythrocytic growth of Plasmodium falciparum through ferrochelatase inhibition but lacks activity in an experimental human infection study. Sci Rep. 2017 Feb 8;7:41975.
27. Seder RA, Chang L-J, Enama ME, Zephir KL, Sarwar UN, Gordon IJ, et al. Protection against malaria by intravenous immunization with a nonreplicating sporozoite vaccine. Science. 2013 Sep 20;341(6152):1359–65.
28. Ramos EL, Mitcham JL, Koller TD, Bonavia A, Usner DW, Balaratnam G, et al. Efficacy and safety of treatment with an anti-m2e monoclonal antibody in experimental human influenza. J Infect Dis. 2015 Apr 1;211(7):1038–44.
29. Pennington SH, Pojar S, Mitsi E, Gritzfeld JF, Nikolaou E, Solórzano C, et al. Polysaccharide-Specific Memory B Cells Predict Protection against Experimental Human Pneumococcal Carriage. Am J Respir Crit Care Med. 2016 Dec 15;194(12):1523–31.

30. Pasay CJ, Rockett R, Sekuloski S, Griffin P, Marquart L, Peatey C, et al. Piperaquine Monotherapy of Drug-Susceptible Plasmodium falciparum Infection Results in Rapid Clearance of Parasitemia but Is Followed by the Appearance of Gametocytemia. J Infect Dis. 2016 Jul 1;214(1):105–13.
31. Nash TE, Herrington DA, Levine MM, Conrad JT, Merritt JW. Antigenic variation of Giardia lamblia in experimental human infections. J Immunol. 1990 Jun 1;144(11):4362–9.
32. Muehling LM, Mai DT, Kwok WW, Heymann PW, Pomés A, Woodfolk JA. Circulating Memory CD4+ T Cells Target Conserved Epitopes of Rhinovirus Capsid Proteins and Respond Rapidly to Experimental Infection in Humans. J Immunol. 2016 Oct 15;197(8):3214–24.
33. Mordmüller B, Surat G, Lagler H, Chakravarty S, Ishizuka AS, Lalremruata A, et al. Sterile protection against human malaria by chemoattenuated PfSPZ vaccine. Nature. 2017 Feb 23;542(7642):445–9.
34. Minassian AM, Satti I, Poulton ID, Meyer J, Hill AVS, McShane H. A human challenge model for Mycobacterium tuberculosis using Mycobacterium bovis bacille Calmette-Guerin. J Infect Dis. 2012 Apr 1;205(7):1035–42.
35. Memoli MJ, Shaw PA, Han A, Czajkowski L, Reed S, Athota R, et al. Evaluation of Antihemagglutinin and Antineuraminidase Antibodies as Correlates of Protection in an Influenza A/H1N1 Virus Healthy Human Challenge Model. mBio. 2016 Apr 19;7(2):e00417-00416.
36. McCarthy JS, Lotharius J, Rückle T, Chalon S, Phillips MA, Elliott S, et al. Safety, tolerability, pharmacokinetics, and activity of the novel long-acting antimalarial DSM265: a two-part first-in-human phase 1a/1b randomised study. Lancet Infect Dis. 2017 Jun;17(6):626–35.
37. McCarthy JS, Rückle T, Djeriou E, Cantalloube C, Ter-Minassian D, Baker M, et al. A Phase II pilot trial to evaluate safety and efficacy of ferroquine against early Plasmodium falciparum in an induced blood-stage malaria infection study. Malar J. 2016 Sep 13;15:469.
38. Mammen MP, Lyons A, Innis BL, Sun W, McKinney D, Chung RCY, et al. Evaluation of dengue virus strains for human challenge studies. Vaccine. 2014 Mar 14;32(13):1488–94.
39. Mallia P, Footitt J, Sotero R, Jepson A, Contoli M, Trujillo-Torralbo M-B, et al. Rhinovirus infection induces degradation of antimicrobial peptides and secondary bacterial infection in chronic obstructive pulmonary disease. Am J Respir Crit Care Med. 2012 Dec 1;186(11):1117–24.
40. Lindesmith L, Moe C, Marionneau S, Ruvoen N, Jiang X, Lindblad L, et al. Human susceptibility and resistance to Norwalk virus infection. Nat Med. 2003 May;9(5):548–53.
41. Keitel WA, Couch RB, Cate TR, Six HR, Baxter BD. Cold recombinant influenza B/Texas/1/84 vaccine virus (CRB 87): attenuation, immunogenicity, and efficacy against homotypic challenge. J Infect Dis. 1990 Jan;161(1):22–6.

42. Jones S, Evans K, McElwaine-Johnn H, Sharpe M, Oxford J, Lambkin-Williams R, et al. DNA vaccination protects against an influenza challenge in a double-blind randomised placebo-controlled phase 1b clinical trial. Vaccine. 2009 Apr 21;27(18):2506–12.
43. Habibi MS, Jozwik A, Makris S, Dunning J, Paras A, DeVincenzo JP, et al. Impaired Antibody-mediated Protection and Defective IgA B-Cell Memory in Experimental Infection of Adults with Respiratory Syncytial Virus. Am J Respir Crit Care Med. 2015 May 1;191(9):1040–9.
44. Gubareva LV, Kaiser L, Matrosovich MN, Soo-Hoo Y, Hayden FG. Selection of influenza virus mutants in experimentally infected volunteers treated with oseltamivir. J Infect Dis. 2001 Feb 15;183(4):523–31.
45. Gritzfeld JF, Wright AD, Collins AM, Pennington SH, Wright AKA, Kadioglu A, et al. Experimental human pneumococcal carriage. J Vis Exp. 2013 Feb 15;(72):50115.
46. Gritzfeld JF, Cremers AJH, Ferwerda G, Ferreira DM, Kadioglu A, Hermans PWM, et al. Density and duration of experimental human pneumococcal carriage. Clin Microbiol Infect. 2014 Dec;20(12):O1145-1151.
47. Graham DY, Opekun AR, Osato MS, El-Zimaity HMT, Lee CK, Yamaoka Y, et al. Challenge model for Helicobacter pylori infection in human volunteers. Gut. 2004 Sep;53(9):1235–43.
48. Graham DY, Estes MK, Gentry LO. Double-blind comparison of bismuth subsalicylate and placebo in the prevention and treatment of enterotoxigenic Escherichia coli-induced diarrhea in volunteers. Gastroenterology. 1983 Nov;85(5):1017–22.
49. Glennie S, Gritzfeld JF, Pennington SH, Garner-Jones M, Coombes N, Hopkins MJ, et al. Modulation of nasopharyngeal innate defenses by viral coinfection predisposes individuals to experimental pneumococcal carriage. Mucosal Immunol. 2016 Jan;9(1):56–67.
50. Frenck R, Bernstein DI, Xia M, Huang P, Zhong W, Parker S, et al. Predicting susceptibility to norovirus GII.4 by use of a challenge model involving humans. J Infect Dis. 2012 Nov;206(9):1386–93.
51. Evans DG, Evans DJ, Opekun AR, Graham DY. Non-replicating oral whole cell vaccine protective against enterotoxigenic Escherichia coli (ETEC) diarrhea: stimulation of anti-CFA (CFA/I) and anti-enterotoxin (anti-LT) intestinal IgA and protection against challenge with ETEC belonging to heterologous serotypes. FEMS Microbiol Immunol. 1988 Dec;1(3):117–25.
52. DuPont HL, Chappell CL, Sterling CR, Okhuysen PC, Rose JB, Jakubowski W. The infectivity of Cryptosporidium parvum in healthy volunteers. N Engl J Med. 1995 Mar 30;332(13):855–9.
53. Dobinson HC, Gibani MM, Jones C, Thomaides-Brears HB, Voysey M, Darton TC, et al. Evaluation of the Clinical and Microbiological Response to Salmonella Paratyphi A Infection in the First Paratyphoid Human Challenge Model. Clin Infect Dis. 2017 Apr 15;64(8):1066–73.
54. Deye GA, Miller RS, Miller L, Salas CJ, Tosh D, Macareo L, et al. Prolonged protection provided by a single dose of atovaquone-proguanil for the chemoprophylaxis of Plasmodium falciparum malaria in a human challenge model. Clin Infect Dis. 2012 Jan 15;54(2):232–9.

55. Deasy AM, Guccione E, Dale AP, Andrews N, Evans CM, Bennett JS, et al. Nasal Inoculation of the Commensal Neisseria lactamica Inhibits Carriage of Neisseria meningitidis by Young Adults: A Controlled Human Infection Study. Clin Infect Dis. 2015 May 15;60(10):1512–20.
56. Croese J, O’neil J, Masson J, Cooke S, Melrose W, Pritchard D, et al. A proof of concept study establishing Necator americanus in Crohn’s patients and reservoir donors. Gut. 2006 Jan;55(1):136–7.
57. Croese J, Giacomin P, Navarro S, Clouston A, McCann L, Dougall A, et al. Experimental hookworm infection and gluten microchallenge promote tolerance in celiac disease. J Allergy Clin Immunol. 2015 Feb;135(2):508–16.
58. Cremers AJ, Zomer AL, Gritzfeld JF, Ferwerda G, van Hijum SA, Ferreira DM, et al. The adult nasopharyngeal microbiome as a determinant of pneumococcal acquisition. Microbiome. 2014;2:44.
59. Collins AM, Wright AD, Mitsi E, Gritzfeld JF, Hancock CA, Pennington SH, et al. First human challenge testing of a pneumococcal vaccine. Double-blind randomized controlled trial. Am J Respir Crit Care Med. 2015 Oct 1;192(7):853–8.
60. Chappell CL, Okhuysen PC, Langer-Curry R, Widmer G, Akiyoshi DE, Tanriverdi S, et al. Cryptosporidium hominis: experimental challenge of healthy adults. Am J Trop Med Hyg. 2006 Nov;75(5):851–7.
61. Bennett JW, Yadava A, Tosh D, Sattabongkot J, Komisar J, Ware LA, et al. Phase 1/2a Trial of Plasmodium vivax Malaria Vaccine Candidate VMP001/AS01B in Malaria-Naive Adults: Safety, Immunogenicity, and Efficacy. PLoS Negl Trop Dis. 2016 Feb;10(2):e0004423.
62. Barroso L, Treanor J, Gubareva L, Hayden FG. Efficacy and tolerability of the oral neuraminidase inhibitor peramivir in experimental human influenza: randomized, controlled trials for prophylaxis and treatment. Antivir Ther. 2005;10(8):901–10.
63. Jongo SA, Shekalaghe SA, Church LWP, Ruben AJ, Schindler T, Zenklusen I, et al. Safety, Immunogenicity, and Protective Efficacy against Controlled Human Malaria Infection of Plasmodium falciparum Sporozoite Vaccine in Tanzanian Adults. Am J Trop Med Hyg. 2018 Aug;99(2):338–49.
64. Herrera S, Fernández O, Manzano MR, Murrain B, Vergara J, Blanco P, et al. Successful sporozoite challenge model in human volunteers with Plasmodium vivax strain derived from human donors. Am J Trop Med Hyg. 2009 Nov;81(5):740–6.
65. Dejon-Agobe JC, Ateba-Ngoa U, Lalremruata A, Homoet A, Engelhorn J, Nouatin OP, et al. Controlled Human Malaria Infection of Healthy Adults With Lifelong Malaria Exposure to Assess Safety, Immunogenicity, and Efficacy of the Asexual Blood Stage Malaria Vaccine Candidate GMZ2. Clin Infect Dis. 2019 Sep 27;69(8):1377–84.
66. Chulay JD, Schneider I, Cosgriff TM, Hoffman SL, Ballou WR, Quakyi IA, et al. Malaria transmitted to humans by mosquitoes infected from cultured Plasmodium falciparum. Am J Trop Med Hyg. 1986 Jan;35(1):66–8.
67. Arévalo-Herrera M, Vásquez-Jiménez JM, Lopez-Perez M, Vallejo AF, Amado-Garavito AB, Céspedes N, et al. Protective Efficacy of Plasmodium vivax Radiation-Attenuated Sporozoites in Colombian Volunteers: A Randomized Controlled Trial. PLoS Negl Trop Dis. 2016 Oct;10(10):e0005070.

68. Arévalo-Herrera M, Forero-Peña DA, Rubiano K, Gómez-Hincapie J, Martínez NL, Lopez-Perez M, et al. Plasmodium vivax sporozoite challenge in malaria-naïve and semi-immune Colombian volunteers. PLoS One. 2014;9(6):e99754.
69. Suntharasamai P, Migasena S, Vongsthongsri U, Supanaranond W, Pitisuttitham P, Supeeranan L, et al. Clinical and bacteriological studies of El Tor cholera after ingestion of known inocula in Thai volunteers. Vaccine. 1992;10(8):502–5.
70. Shekalaghe S, Rutaihwa M, Billingsley PF, Chemba M, Daubenberger CA, James ER, et al. Controlled human malaria infection of Tanzanians by intradermal injection of aseptic, purified, cryopreserved Plasmodium falciparum sporozoites. Am J Trop Med Hyg. 2014 Sep;91(3):471–80.
71. Kapulu MC, Njuguna P, Hamaluba M, Kimani D, Ngoi JM, Musembi J, et al. Safety and PCR monitoring in 161 semi-immune Kenyan adults following controlled human malaria infection. JCI Insight. 2021 Sep 8;6(17):146443.
72. Frenck RW, Conti V, Ferruzzi P, Ndiaye AGW, Parker S, McNeal MM, et al. Efficacy, safety, and immunogenicity of the Shigella sonnei 1790GAHB GMMA candidate vaccine: Results from a phase 2b randomized, placebo-controlled challenge study in adults. EClinicalMedicine. 2021 Sep;39:101076.
73. Chapman PR, Webster R, Giacomin P, Llewellyn S, Becker L, Pearson MS, et al. Vaccination of human participants with attenuated Necator americanus hookworm larvae and human challenge in Australia: a dose-finding study and randomised, placebo-controlled, phase 1 trial. Lancet Infect Dis. 2021 Dec;21(12):1725–36.
74. Talaat KR, Alaimo C, Martin P, Bourgeois AL, Dreyer AM, Kaminski RW, et al. Human challenge study with a Shigella bioconjugate vaccine: Analyses of clinical efficacy and correlate of protection. EBioMedicine. 2021 Apr;66:103310.
75. McCarthy JS, Donini C, Chalon S, Woodford J, Marquart L, Collins KA, et al. A Phase 1, Placebo-controlled, Randomized, Single Ascending Dose Study and a Volunteer Infection Study to Characterize the Safety, Pharmacokinetics, and Antimalarial Activity of the Plasmodium Phosphatidylinositol 4-Kinase Inhibitor MMV390048. Clin Infect Dis. 2020 Dec 17;71(10):e657–64.
76. Zuckerman MA, Cox RJ, Oxford JS. Attenuation of virulence in influenza B viral infection of volunteers. J Infect. 1994 Jan;28(1):41–8.
77. Winther B, Buchert D, Turner RB, Hendley JO, Tschaikin M. Decreased rhinovirus shedding after intranasal oxymetazoline application in adults with induced colds compared with intranasal saline. Am J Rhinol Allergy. 2010 Oct;24(5):374–7.
78. Weidner TG, Anderson BN, Kaminsky LA, Dick EC, Schurr T. Effect of a rhinovirus-caused upper respiratory illness on pulmonary function test and exercise responses. Med Sci Sports Exerc. 1997 May;29(5):604–9.
79. Walker JB, Hussey EK, Treanor JJ, Montalvo A, Hayden FG. Effects of the neuraminidase inhibitor zanamavir on otologic manifestations of experimental human influenza. J Infect Dis. 1997 Dec;176(6):1417–22.
80. Tyrrell DA, Cohen S, Schlarb JE. Signs and symptoms in common colds. Epidemiol Infect. 1993 Aug;111(1):143–56.
81. Turner RB, Dutko FJ, Goldstein NH, Lockwood G, Hayden FG. Efficacy of oral WIN 54954 for prophylaxis of experimental rhinovirus infection. Antimicrob Agents Chemother. 1993 Feb;37(2):297–300.

82. Turner RB, Riker DK, Gangemi JD. Ineffectiveness of echinacea for prevention of experimental rhinovirus colds. Antimicrob Agents Chemother. 2000 Jun;44(6):1708–9.
83. Turner RB, Cetnarowski WE. Effect of treatment with zinc gluconate or zinc acetate on experimental and natural colds. Clin Infect Dis. 2000 Nov;31(5):1202–8.
84. Turner RB, Winther B, Hendley JO, Mygind N, Gwaltney JM. Sites of virus recovery and antigen detection in epithelial cells during experimental rhinovirus infection. Acta Otolaryngol Suppl. 1984;413:9–14.
85. Tribble DR, Baqar S, Scott DA, Oplinger ML, Trespalacios F, Rollins D, et al. Assessment of the duration of protection in Campylobacter jejuni experimental infection in humans. Infect Immun. 2010 Apr;78(4):1750–9.
86. Tian SF, Zhang LX, Murphy BR. Characterization of the genotype and level of attenuation of an influenza A reassortant virus produced by mating the Xia-ts donor virus with A/Beijing/70 (H1N1) wild type virus. Vaccine. 1984 Sep;2(3):189–92.
87. Tacket CO, Forrest B, Morona R, Attridge SR, LaBrooy J, Tall BD, et al. Safety, immunogenicity, and efficacy against cholera challenge in humans of a typhoid-cholera hybrid vaccine derived from Salmonella typhi Ty21a. Infect Immun. 1990 Jun;58(6):1620–7.
88. Sperber SJ, Shah LP, Gilbert RD, Ritchey TW, Monto AS. Echinacea purpurea for prevention of experimental rhinovirus colds. Clin Infect Dis. 2004 May 15;38(10):1367–71.
89. Sperber SJ, Doyle WJ, McBride TP, Sorrentino JV, Riker DK, Hayden FG. Otologic effects of interferon beta serine in experimental rhinovirus colds. Arch Otolaryngol Head Neck Surg. 1992 Sep;118(9):933–6.
90. Sperber SJ, Hendley JO, Hayden FG, Riker DK, Sorrentino JV, Gwaltney JM. Effects of naproxen on experimental rhinovirus colds. A randomized, double-blind, controlled trial. Ann Intern Med. 1992 Jul 1;117(1):37–41.
91. Snyder MH, Betts RF, DeBorde D, Tierney EL, Clements ML, Herrington D, et al. Four viral genes independently contribute to attenuation of live influenza A/Ann Arbor/6/60 (H2N2) cold-adapted reassortant virus vaccines. J Virol. 1988 Feb;62(2):488–95.
92. Snyder MH, Clements ML, Betts RF, Dolin R, Buckler-White AJ, Tierney EL, et al. Evaluation of live avian-human reassortant influenza A H3N2 and H1N1 virus vaccines in seronegative adult volunteers. J Clin Microbiol. 1986 May;23(5):852–7.
93. Snyder MH, Clements ML, De Borde D, Maassab HF, Murphy BR. Attenuation of wild-type human influenza A virus by acquisition of the PA polymerase and matrix protein genes of influenza A/Ann Arbor/6/60 cold-adapted donor virus. J Clin Microbiol. 1985 Nov;22(5):719–25.
94. Sears SD, Clements ML. Protective efficacy of low-dose amantadine in adults challenged with wild-type influenza A virus. Antimicrob Agents Chemother. 1987 Oct;31(10):1470–3.
95. Schiff GM, Sherwood JR. Clinical activity of pleconaril in an experimentally induced coxsackievirus A21 respiratory infection. J Infect Dis. 2000 Jan;181(1):20–6.
96. Roestenberg M, McCall M, Hopman J, Wiersma J, Luty AJF, van Gemert GJ, et al. Protection against a malaria challenge by sporozoite inoculation. N Engl J Med. 2009 Jul 30;361(5):468–77.

97. Reuman PD, Bernstein DI, Keefer MC, Young EC, Sherwood JR, Schiff GM. Efficacy and safety of low dosage amantadine hydrochloride as prophylaxis for influenza A. Antiviral Res. 1989 Feb;11(1):27–40.
98. Atmar RL, Opekun AR, Gilger MA, Estes MK, Crawford SE, Neill FH, et al. Determination of the 50% human infectious dose for Norwalk virus. J Infect Dis. 2014 Apr 1;209(7):1016–22.
99. Atmar RL, Opekun AR, Gilger MA, Estes MK, Crawford SE, Neill FH, et al. Norwalk virus shedding after experimental human infection. Emerg Infect Dis. 2008 Oct;14(10):1553–7.
100. Murphy BR, Chanock RM, Clements ML, Anthony WC, Sear AJ, Cisneros LA, et al. Evaluation of A/Alaska/6/77 (H3N2) cold-adapted recombinant viruses derived from A/Ann Arbor/6/60 cold-adapted donor virus in adult seronegative volunteers. Infect Immun. 1981 May;32(2):693–7.
101. Clements ML, Wisseman CL, Woodward TE, Fiset P, Dumler JS, McNamee W, et al. Reactogenicity, immunogenicity, and efficacy of a chick embryo cell-derived vaccine for Rocky Mountain spotted fever. J Infect Dis. 1983 Nov;148(5):922–30.
102. Black RE, Levine MM, Clements ML, Losonsky G, Herrington D, Berman S, et al. Prevention of shigellosis by a Salmonella typhi-Shigella sonnei bivalent vaccine. J Infect Dis. 1987 Jun;155(6):1260–5.
103. Peterson KM, O'Shea M, Stam W, Mohede ICM, Patrie JT, Hayden FG. Effects of dietary supplementation with conjugated linoleic acid on experimental human rhinovirus infection and illness. Antivir Ther. 2009;14(1):33–43.
104. Oxford JS, Schild GC, Corcoran T, Newman R, Major D, Robertson J, et al. A host-cell-selected variant of influenza B virus with a single nucleotide substitution in HA affecting a potential glycosylation site was attenuated in virulence for volunteers. Arch Virol. 1990;110(1–2):37–46.
105. Noah TL, Becker S. Chemokines in nasal secretions of normal adults experimentally infected with respiratory syncytial virus. Clin Immunol. 2000 Oct;97(1):43–9.
106. Murphy AW, Platts-Mills TA, Lobo M, Hayden F. Respiratory nitric oxide levels in experimental human influenza. Chest. 1998 Aug;114(2):452–6.
107. Moss DM, Chappell CL, Okhuysen PC, DuPont HL, Arrowood MJ, Hightower AW, et al. The antibody response to 27-, 17-, and 15-kDa Cryptosporidium antigens following experimental infection in humans. J Infect Dis. 1998 Sep;178(3):827–33.
108. Black RE, Levine MM, Clements ML, Hughes TP, Blaser MJ. Experimental Campylobacter jejuni infection in humans. J Infect Dis. 1988 Mar;157(3):472–9.
109. Mallia P, Message SD, Kebadze T, Parker HL, Kon OM, Johnston SL. An experimental model of rhinovirus induced chronic obstructive pulmonary disease exacerbations: a pilot study. Respir Res. 2006 Sep 6;7:116.
110. Mackowiak PA, Wasserman SS, Tacket CO, Vaughn DW, Eckels KH, Dubois DR, et al. Quantitative relationship between oral temperature and severity of illness following inoculation with candidate attenuated dengue virus vaccines. Clin Infect Dis. 1994 Nov;19(5):948–50.

111. Mackowiak PA, Wasserman SS, Levine MM. An analysis of the quantitative relationship between oral temperature and severity of illness in experimental shigellosis. J Infect Dis. 1992 Nov;166(5):1181–4.
112. Lemanske RF, Dick EC, Swenson CA, Vrtis RF, Busse WW. Rhinovirus upper respiratory infection increases airway hyperreactivity and late asthmatic reactions. J Clin Invest. 1989 Jan;83(1):1–10.
113. Kotloff KL, Herrington DA, Hale TL, Newland JW, Van De Verg L, Cogan JP, et al. Safety, immunogenicity, and efficacy in monkeys and humans of invasive Escherichia coli K-12 hybrid vaccine candidates expressing Shigella flexneri 2a somatic antigen. Infect Immun. 1992 Jun;60(6):2218–24.
114. Kotloff KL, Nataro JP, Losonsky GA, Wasserman SS, Hale TL, Taylor DN, et al. A modified Shigella volunteer challenge model in which the inoculum is administered with bicarbonate buffer: clinical experience and implications for Shigella infectivity. Vaccine. 1995 Nov;13(16):1488–94.
115. Hobbins TE, Hughes TP, Rennels MB, Murphy BR, Levine MM. Bronchial reactivity in experimental infections with influenza virus. J Infect Dis. 1982 Oct;146(4):468–71.
116. Higgins PG, al-Nakib W, Barrow GI, Tyrrell DA. Recombinant human interferon-gamma as prophylaxis against rhinovirus colds in volunteers. J Interferon Res. 1988 Oct;8(5):591–6.
117. Message SD, Laza-Stanca V, Mallia P, Parker HL, Zhu J, Kebadze T, et al. Rhinovirus-induced lower respiratory illness is increased in asthma and related to virus load and Th1/2 cytokine and IL-10 production. Proc Natl Acad Sci U S A. 2008 Sep 9;105(36):13562–7.
118. Herrington DA, Van de Verg L, Formal SB, Hale TL, Tall BD, Cryz SJ, et al. Studies in volunteers to evaluate candidate Shigella vaccines: further experience with a bivalent Salmonella typhi-Shigella sonnei vaccine and protection conferred by previous Shigella sonnei disease. Vaccine. 1990 Aug;8(4):353–7.
119. Hayden FG, Treanor JJ, Fritz RS, Lobo M, Betts RF, Miller M, et al. Use of the oral neuraminidase inhibitor oseltamivir in experimental human influenza: randomized controlled trials for prevention and treatment. JAMA. 1999 Oct 6;282(13):1240–6.
120. Hayden FG, Tunkel AR, Treanor JJ, Betts RF, Allerheiligen S, Harris J. Oral LY217896 for prevention of experimental influenza A virus infection and illness in humans. Antimicrob Agents Chemother. 1994 May;38(5):1178–81.
121. Hayden FG, Andries K, Janssen PA. Safety and efficacy of intranasal pirodavir (R77975) in experimental rhinovirus infection. Antimicrob Agents Chemother. 1992 Apr;36(4):727–32.
122. Hayden FG, Treanor JJ, Betts RF, Lobo M, Esinhart JD, Hussey EK. Safety and efficacy of the neuraminidase inhibitor GG167 in experimental human influenza. JAMA. 1996 Jan 24;275(4):295–9.
123. Hayden FG, Gwaltney JM, Colonno RJ. Modification of experimental rhinovirus colds by receptor blockade. Antiviral Res. 1988 Jul;9(4):233–47.

124. Hayden FG, Zylidnikov DM, Iljenko VI, Padolka YV. Comparative therapeutic effect of aerosolized and oral rimantadine HCl in experimental human influenza A virus infection. Antiviral Res. 1982 Aug;2(3):147–53.
125. Harris JM, Gwaltney JM. Incubation periods of experimental rhinovirus infection and illness. Clin Infect Dis. 1996 Dec;23(6):1287–90.
126. Gwaltney JM, Winther B, Patrie JT, Hendley JO. Combined antiviral-antimediator treatment for the common cold. J Infect Dis. 2002 Jul 15;186(2):147–54.
127. Gwaltney JM, Park J, Paul RA, Edelman DA, O'Connor RR, Turner RB. Randomized controlled trial of clemastine fumarate for treatment of experimental rhinovirus colds. Clin Infect Dis. 1996 Apr;22(4):656–62.
128. Gwaltney JM, Druce HM. Efficacy of brompheniramine maleate for the treatment of rhinovirus colds. Clin Infect Dis. 1997 Nov;25(5):1188–94.
129. Gustafson LM, Proud D, Hendley JO, Hayden FG, Gwaltney JM. Oral prednisone therapy in experimental rhinovirus infections. J Allergy Clin Immunol. 1996 Apr;97(4):1009–14.
130. Grünberg K, Sharon RF, Sont JK, In 't Veen JC, Van Schadewijk WA, De Klerk EP, et al. Rhinovirus-induced airway inflammation in asthma: effect of treatment with inhaled corticosteroids before and during experimental infection. Am J Respir Crit Care Med. 2001 Nov 15;164(10 Pt 1):1816–22.
131. Gern JE, Stone CK, Nakano M, Muchmore DB, de la Peña A, Park S, et al. Effect of upper respiratory tract infection on AIR inhaled insulin pharmacokinetics and glucodynamics in healthy subjects. Clin Pharmacol Ther. 2008 Feb;83(2):307–11.
132. Gentile DA, Fireman P, Skoner DP. Elevations of local leukotriene C4 levels during viral upper respiratory tract infections. Ann Allergy Asthma Immunol. 2003 Sep;91(3):270–4.
133. Feldman PJ, Cohen S, Doyle WJ, Skoner DP, Gwaltney JM. The impact of personality on the reporting of unfounded symptoms and illness. J Pers Soc Psychol. 1999 Aug;77(2):370–8.
134. Drake CL, Roehrs TA, Royer H, Koshorek G, Turner RB, Roth T. Effects of an experimentally induced rhinovirus cold on sleep, performance, and daytime alertness. Physiol Behav. 2000 Oct 1;71(1–2):75–81.
135. Doyle WJ, Gentile DA, Cohen S. Emotional style, nasal cytokines, and illness expression after experimental rhinovirus exposure. Brain Behav Immun. 2006 Mar;20(2):175–81.
136. Doyle WJ, Riker DK, McBride TP, Hayden FG, Hendley JO, Swarts JD, et al. Therapeutic effects of an anticholinergic-sympathomimetic combination in induced rhinovirus colds. Ann Otol Rhinol Laryngol. 1993 Jul;102(7):521–7.
137. Doyle WJ, Skoner DP, Alper CM, Allen G, Moody SA, Seroky JT, et al. Effect of rimantadine treatment on clinical manifestations and otologic complications in adults experimentally infected with influenza A (H1N1) virus. J Infect Dis. 1998 May;177(5):1260–5.

138. Cohen S, Doyle WJ, Skoner DP, Rabin BS, Gwaltney JM. Social ties and susceptibility to the common cold. JAMA. 1997 Jun 25;277(24):1940–4.
139. Doyle WJ, Skoner DP, Hayden F, Buchman CA, Seroky JT, Fireman P. Nasal and otologic effects of experimental influenza A virus infection. Ann Otol Rhinol Laryngol. 1994 Jan;103(1):59–69.
140. Cummings JF, Spring MD, Schwenk RJ, Ockenhouse CF, Kester KE, Polhemus ME, et al. Recombinant Liver Stage Antigen-1 (LSA-1) formulated with AS01 or AS02 is safe, elicits high titer antibody and induces IFN-gamma/IL-2 CD4+ T cells but does not protect against experimental Plasmodium falciparum infection. Vaccine. 2010 Jul 12;28(31):5135–44.
141. Coster TS, Wolf MK, Hall ER, Cassels FJ, Taylor DN, Liu CT, et al. Immune response, ciprofloxacin activity, and gender differences after human experimental challenge by two strains of enterotoxigenic Escherichia coli. Infect Immun. 2007 Jan;75(1):252–9.
142. Cohen S, Doyle WJ, Skoner DP. Psychological stress, cytokine production, and severity of upper respiratory illness. Psychosom Med. 1999 Apr;61(2):175–80.
143. Clements ML, Belshe RB, King J, Newman F, Westblom TU, Tierney EL, et al. Evaluation of bovine, cold-adapted human, and wild-type human parainfluenza type 3 viruses in adult volunteers and in chimpanzees. J Clin Microbiol. 1991 Jun;29(6):1175–82.
144. Clements ML, O'Donnell S, Levine MM, Chanock RM, Murphy BR. Dose response of A/Alaska/6/77 (H3N2) cold-adapted reassortant vaccine virus in adult volunteers: role of local antibody in resistance to infection with vaccine virus. Infect Immun. 1983 Jun;40(3):1044–51.
145. Clements ML, Snyder MH, Sears SD, Maassab HF, Murphy BR. Evaluation of the infectivity, immunogenicity, and efficacy of live cold-adapted influenza B/Ann Arbor/1/86 reassortant virus vaccine in adult volunteers. J Infect Dis. 1990 May;161(5):869–77.
146. Clements ML, Betts RF, Tierney EL, Murphy BR. Resistance of adults to challenge with influenza A wild-type virus after receiving live or inactivated virus vaccine. J Clin Microbiol. 1986 Jan;23(1):73–6.
147. Clements ML, Betts RF, Murphy BR. Advantage of live attenuated cold-adapted influenza A virus over inactivated vaccine for A/Washington/80 (H3N2) wild-type virus infection. Lancet. 1984 Mar 31;1(8379):705–8.
148. Shmuklarsky MJ, Boudreau EF, Pang LW, Smith JI, Schneider I, Fleckenstein L, et al. Failure of doxycycline as a causal prophylactic agent against Plasmodium falciparum malaria in healthy nonimmune volunteers. Ann Intern Med. 1994 Feb 15;120(4):294–9.
149. Rickman LS, Jones TR, Long GW, Paparello S, Schneider I, Paul CF, et al. Plasmodium falciparum-infected Anopheles stephensi inconsistently transmit malaria to humans. Am J Trop Med Hyg. 1990 Nov;43(5):441–5.
150. Gordon DM, McGovern TW, Krzych U, Cohen JC, Schneider I, LaChance R, et al. Safety, immunogenicity, and efficacy of a recombinantly produced Plasmodium falciparum circumsporozoite protein-hepatitis B surface antigen subunit vaccine. J Infect Dis. 1995 Jun;171(6):1576–85.

151. Fries LF, Gordon DM, Schneider I, Beier JC, Long GW, Gross M, et al. Safety, immunogenicity, and efficacy of a Plasmodium falciparum vaccine comprising a circumsporozoite protein repeat region peptide conjugated to Pseudomonas aeruginosa toxin A. Infect Immun. 1992 May;60(5):1834–9.
152. Egan JE, Hoffman SL, Haynes JD, Sadoff JC, Schneider I, Grau GE, et al. Humoral immune responses in volunteers immunized with irradiated Plasmodium falciparum sporozoites. Am J Trop Med Hyg. 1993 Aug;49(2):166–73.
153. Ballou WR, Hoffman SL, Sherwood JA, Hollingdale MR, Neva FA, Hockmeyer WT, et al. Safety and efficacy of a recombinant DNA Plasmodium falciparum sporozoite vaccine. Lancet. 1987 Jun 6;1(8545):1277–81.
154. Cannon JG, Tompkins RG, Gelfand JA, Michie HR, Stanford GG, van der Meer JW, et al. Circulating interleukin-1 and tumor necrosis factor in septic shock and experimental endotoxin fever. J Infect Dis. 1990 Jan;161(1):79–84.
155. Calhoun WJ, Swenson CA, Dick EC, Schwartz LB, Lemanske RF, Busse WW. Experimental rhinovirus 16 infection potentiates histamine release after antigen bronchoprovocation in allergic subjects. Am Rev Respir Dis. 1991 Dec;144(6):1267–73.
156. Calfee DP, Peng AW, Cass LM, Lobo M, Hayden FG. Safety and efficacy of intravenous zanamivir in preventing experimental human influenza A virus infection. Antimicrob Agents Chemother. 1999 Jul;43(7):1616–20.
157. Buchman CA, Doyle WJ, Pilcher O, Gentile DA, Skoner DP. Nasal and otologic effects of experimental respiratory syncytial virus infection in adults. Am J Otolaryngol. 2002 Apr;23(2):70–5.
158. Bjornson AB, Mellencamp MA, Schiff GM. Complement is activated in the upper respiratory tract during influenza virus infection. Am Rev Respir Dis. 1991 May;143(5 Pt 1):1062–6.
159. Anderson MJ, Higgins PG, Davis LR, Willman JS, Jones SE, Kidd IM, et al. Experimental parvoviral infection in humans. J Infect Dis. 1985 Aug;152(2):257–65.
160. Alper CM, Doyle WJ, Skoner DP, Buchman CA, Seroky JT, Gwaltney JM, et al. Prechallenge antibodies: moderators of infection rate, signs, and symptoms in adults experimentally challenged with rhinovirus type 39. Laryngoscope. 1996 Oct;106(10):1298–305.
161. Al-Nakib W, Higgins PG, Barrow I, Tyrrell DA, Lenox-Smith I, Ishitsuka H. Intranasal chalcone, Ro 09-0410, as prophylaxis against rhinovirus infection in human volunteers. J Antimicrob Chemother. 1987 Dec;20(6):887–92.
162. Al-Nakib W, Higgins PG, Willman J, Tyrrell DA, Swallow DL, Hurst BC, et al. Prevention and treatment of experimental influenza A virus infection in volunteers with a new antiviral ICI 130,685. J Antimicrob Chemother. 1986 Jul;18(1):119–29.
163. Abisheganaden JA, Avila PC, Kishiyama JL, Liu J, Yagi S, Schnurr D, et al. Effect of clarithromycin on experimental rhinovirus-16 colds: a randomized, double-blind, controlled trial. Am J Med. 2000 Apr 15;108(6):453–9.
164. Watts RE, Odedra A, Marquart L, Webb L, Abd-Rahman AN, Cascales L, et al. Safety and parasite clearance of artemisinin-resistant Plasmodium falciparum infection: A pilot and a randomised volunteer infection study in Australia. PLoS Med. 2020 Aug;17(8):e1003203.

165. Watson JM, Francis JN, Mesens S, Faiman GA, Makin J, Patriarca P, et al. Characterisation of a wild-type influenza (A/H1N1) virus strain as an experimental challenge agent in humans. Virol J. 2015 Feb 3;12:13.
166. Roestenberg M, Teirlinck AC, McCall MBB, Teelen K, Makamdop KN, Wiersma J, et al. Long-term protection against malaria after experimental sporozoite inoculation: an open-label follow-up study. Lancet. 2011 May 21;377(9779):1770–6.
167. Bijker EM, Schats R, Obiero JM, Behet MC, van Gemert G-J, van de Vegte-Bolmer M, et al. Sporozoite immunization of human volunteers under mefloquine prophylaxis is safe, immunogenic and protective: a double-blind randomized controlled clinical trial. PLoS One. 2014;9(11):e112910.
168. McCall MBB, Wammes LJ, Langenberg MCC, van Gemert G-J, Walk J, Hermsen CC, et al. Infectivity of Plasmodium falciparum sporozoites determines emerging parasitemia in infected volunteers. Sci Transl Med. 2017 Jun 21;9(395):eaag2490.
169. van Hoffen E, Mercenier A, Vidal K, Benyacoub J, Schloesser J, Kardinaal A, et al. Characterization of the pathophysiological determinants of diarrheagenic Escherichia coli infection using a challenge model in healthy adults. Sci Rep. 2021 Mar 15;11(1):6060.
170. Teirlinck AC, Roestenberg M, van de Vegte-Bolmer M, Scholzen A, Heinrichs MJL, Siebelink-Stoter R, et al. NF135.C10: a new Plasmodium falciparum clone for controlled human malaria infections. J Infect Dis. 2013 Feb 15;207(4):656–60.
171. Talley AK, Healy SA, Finney OC, Murphy SC, Kublin J, Salas CJ, et al. Safety and comparability of controlled human Plasmodium falciparum infection by mosquito bite in malaria-naïve subjects at a new facility for sporozoite challenge. PLoS One. 2014;9(11):e109654.
172. Talaat KR, Porter CK, Bourgeois AL, Lee TK, Duplessis CA, Maciel M, et al. Oral delivery of Hyperimmune bovine serum antibodies against CS6-expressing enterotoxigenic Escherichia coli as a prophylactic against diarrhea. Gut Microbes. 2020 Nov 9;12(1):1732852.
173. Talaat KR, Porter CK, Jaep KM, Duplessis CA, Gutierrez RL, Maciel M, et al. Refinement of the CS6-expressing enterotoxigenic Escherichia coli strain B7A human challenge model: A randomized trial. PLoS One. 2020;15(12):e0239888.
174. Skrede S, Steinsland H, Sommerfelt H, Aase A, Brandtzaeg P, Langeland N, et al. Experimental infection of healthy volunteers with enterotoxigenic Escherichia coli wild-type strain TW10598 in a hospital ward. BMC Infect Dis. 2014 Sep 4;14:482.
175. Sinha A, Lutter R, Xu B, Dekker T, Dierdorp B, Sterk PJ, et al. Loss of adaptive capacity in asthmatic patients revealed by biomarker fluctuation dynamics after rhinovirus challenge. Elife. 2019 Nov 5;8:e47969.
176. Sheehy SH, Spencer AJ, Douglas AD, Sim BKL, Longley RJ, Edwards NJ, et al. Optimising Controlled Human Malaria Infection Studies Using Cryopreserved P. falciparum Parasites Administered by Needle and Syringe. PLoS One. 2013;8(6):e65960.
177. Seitz SR, Leon JS, Schwab KJ, Lyon GM, Dowd M, McDaniels M, et al. Norovirus infectivity in humans and persistence in water. Appl Environ Microbiol. 2011 Oct;77(19):6884–8.
178. Sakkestad ST, Steinsland H, Skrede S, Kleppa E, Lillebø K, Sævik M, et al. Experimental Infection of Human Volunteers with the Heat-Stable Enterotoxin-Producing Enterotoxigenic Escherichia coli Strain TW11681. Pathogens. 2019 Jun 22;8(2):E84.

179. Sakkestad ST, Steinsland H, Skrede S, Lillebø K, Skutlaberg DH, Guttormsen AB, et al. A new human challenge model for testing heat-stable toxin-based vaccine candidates for enterotoxigenic Escherichia coli diarrhea - dose optimization, clinical outcomes, and CD4+ T cell responses. PLoS Negl Trop Dis. 2019 Oct;13(10):e0007823.
180. Rylance J, de Steenhuijsen Piters WAA, Mina MJ, Bogaert D, French N, Ferreira DM, et al. Two Randomized Trials of the Effect of Live Attenuated Influenza Vaccine on Pneumococcal Colonization. Am J Respir Crit Care Med. 2019 May 1;199(9):1160–3.
181. Roestenberg M, Bijker EM, Sim BKL, Billingsley PF, James ER, Bastiaens GJH, et al. Controlled human malaria infections by intradermal injection of cryopreserved Plasmodium falciparum sporozoites. Am J Trop Med Hyg. 2013 Jan;88(1):5–13.
182. Reuling IJ, van de Schans LA, Coffeng LE, Lanke K, Meerstein-Kessel L, Graumans W, et al. A randomized feasibility trial comparing four antimalarial drug regimens to induce Plasmodium falciparum gametocytemia in the controlled human malaria infection model. Elife. 2018 Feb 27;7:e31549.
183. Pleguezuelos O, James E, Fernandez A, Lopes V, Rosas LA, Cervantes-Medina A, et al. Efficacy of FLU-v, a broad-spectrum influenza vaccine, in a randomized phase IIb human influenza challenge study. NPJ Vaccines. 2020;5(1):22.
184. Pitisuttithum P, Islam D, Chamnanchanunt S, Ruamsap N, Khantapura P, Kaewkungwal J, et al. Clinical Trial of an Oral Live Shigella sonnei Vaccine Candidate, WRSS1, in Thai Adults. Clin Vaccine Immunol. 2016 Jul;23(7):564–75.
185. Payne RO, Milne KH, Elias SC, Edwards NJ, Douglas AD, Brown RE, et al. Demonstration of the Blood-Stage Plasmodium falciparum Controlled Human Malaria Infection Model to Assess Efficacy of the P. falciparum Apical Membrane Antigen 1 Vaccine, FMP2.1/AS01. J Infect Dis. 2016 Jun 1;213(11):1743–51.
186. Nguyen-Van-Tam JS, Killingley B, Enstone J, Hewitt M, Pantelic J, Grantham ML, et al. Minimal transmission in an influenza A (H3N2) human challenge-transmission model within a controlled exposure environment. PLoS Pathog. 2020 Jul;16(7):e1008704.
187. Murphy SC, Duke ER, Shipman KJ, Jensen RL, Fong Y, Ferguson S, et al. A Randomized Trial Evaluating the Prophylactic Activity of DSM265 Against Preerythrocytic Plasmodium falciparum Infection During Controlled Human Malarial Infection by Mosquito Bites and Direct Venous Inoculation. J Infect Dis. 2018 Feb 14;217(5):693–702.
188. Mordmüller B, Supan C, Sim KL, Gómez-Pérez GP, Ospina Salazar CL, Held J, et al. Direct venous inoculation of Plasmodium falciparum sporozoites for controlled human malaria infection: a dose-finding trial in two centres. Malar J. 2015 Mar 18;14:117.
189. Minhinnick A, Harris S, Wilkie M, Peter J, Stockdale L, Manjaly-Thomas Z-R, et al. Optimization of a Human Bacille Calmette-Guérin Challenge Model: A Tool to Evaluate Antimycobacterial Immunity. J Infect Dis. 2016 Mar 1;213(5):824–30.
190. McClain MT, Henao R, Williams J, Nicholson B, Veldman T, Hudson L, et al. Differential evolution of peripheral cytokine levels in symptomatic and asymptomatic responses to experimental influenza virus challenge. Clin Exp Immunol. 2016 Mar;183(3):441–51.
191. McCarthy JS, Griffin PM, Sekuloski S, Bright AT, Rockett R, Looke D, et al. Experimentally induced blood-stage Plasmodium vivax infection in healthy volunteers. J Infect Dis. 2013 Nov 15;208(10):1688–94.

192. McCarthy JS, Baker M, O'Rourke P, Marquart L, Griffin P, Hooft van Huijsduijnen R, et al. Efficacy of OZ439 (artefenomel) against early Plasmodium falciparum blood-stage malaria infection in healthy volunteers. J Antimicrob Chemother. 2016 Sep;71(9):2620–7.
193. McCarthy JS, Marquart L, Sekuloski S, Trenholme K, Elliott S, Griffin P, et al. Linking Murine and Human Plasmodium falciparum Challenge Models in a Translational Path for Antimalarial Drug Development. Antimicrob Agents Chemother. 2016 Jun;60(6):3669–75.
194. McBride JM, Lim JJ, Burgess T, Deng R, Derby MA, Maia M, et al. Phase 2 Randomized Trial of the Safety and Efficacy of MHAA4549A, a Broadly Neutralizing Monoclonal Antibody, in a Human Influenza A Virus Challenge Model. Antimicrob Agents Chemother. 2017 Nov;61(11):e01154-17.
195. Mateo R, Lindesmith LC, Garg SJ, Gottlieb K, Lin K, Said S, et al. Production and Clinical Evaluation of Norwalk GI.1 Virus Lot 001-09NV in Norovirus Vaccine Development. J Infect Dis. 2020 Mar 2;221(6):919–26.
196. Lyke KE, Laurens M, Adams M, Billingsley PF, Richman A, Loyevsky M, et al. Plasmodium falciparum malaria challenge by the bite of aseptic Anopheles stephensi mosquitoes: results of a randomized infectivity trial. PLoS One. 2010 Oct 21;5(10):e13490.
197. Livezey J, Twomey P, Morrison M, Cicatelli S, Duncan EH, Hamer M, et al. An open label study of the safety and efficacy of a single dose of weekly chloroquine and azithromycin administered for malaria prophylaxis in healthy adults challenged with 7G8 chloroquine-resistant Plasmodium falciparum in a controlled human malaria infection model. Malar J. 2020 Sep 16;19(1):336.
198. Lillie PJ, Duncan CJA, Sheehy SH, Meyer J, O'Hara GA, Gilbert SC, et al. Distinguishing malaria and influenza: early clinical features in controlled human experimental infection studies. Travel Med Infect Dis. 2012 Jul;10(4):192–6.
199. Lell B, Mordmüller B, Dejon Agobe J-C, Honkpehedji J, Zinsou J, Mengue JB, et al. Impact of Sickle Cell Trait and Naturally Acquired Immunity on Uncomplicated Malaria after Controlled Human Malaria Infection in Adults in Gabon. Am J Trop Med Hyg. 2018 Feb;98(2):508–15.
200. Laurens MB, Billingsley P, Richman A, Eappen AG, Adams M, Li T, et al. Successful human infection with P. falciparum using three aseptic Anopheles stephensi mosquitoes: a new model for controlled human malaria infection. PLoS One. 2013;8(7):e68969.
201. Langenberg MCC, Wammes LJ, McCall MBB, Bijker EM, van Gemert G-J, Graumans W, et al. Controlled Human Malaria Infection with Graded Numbers of Plasmodium falciparum NF135.C10- or NF166.C8-Infected Mosquitoes. Am J Trop Med Hyg. 2018 Sep;99(3):709–12.
202. Lambkin-Williams R, Gelder C, Broughton R, Mallett CP, Gilbert AS, Mann A, et al. An Intranasal Proteosome-Adjuvanted Trivalent Influenza Vaccine Is Safe, Immunogenic & Efficacious in the Human Viral Influenza Challenge Model. Serum IgG & Mucosal IgA Are Important Correlates of Protection against Illness Associated with Infection. PLoS One. 2016;11(12):e0163089.

203. Krause A, Dingemanse J, Mathis A, Marquart L, Möhrle JJ, McCarthy JS. Pharmacokinetic/pharmacodynamic modelling of the antimalarial effect of Actelion-451840 in an induced blood stage malaria study in healthy subjects. Br J Clin Pharmacol. 2016 Aug;82(2):412–21.
204. Jin C, Gibani MM, Moore M, Juel HB, Jones E, Meiring J, et al. Efficacy and immunogenicity of a Vi-tetanus toxoid conjugate vaccine in the prevention of typhoid fever using a controlled human infection model of Salmonella Typhi: a randomised controlled, phase 2b trial. Lancet. 2017 Dec 2;390(10111):2472–80.
205. Young RS, Fortney KR, Gelfanova V, Phillips CL, Katz BP, Hood AF, et al. Expression of cytolethal distending toxin and hemolysin is not required for pustule formation by Haemophilus ducreyi in human volunteers. Infect Immun. 2001 Mar;69(3):1938–42.
206. Young RS, Filiatrault MJ, Fortney KR, Hood AF, Katz BP, Munson RS, et al. Haemophilus ducreyi lipooligosaccharide mutant defective in expression of beta-1,4-glucosyltransferase is virulent in humans. Infect Immun. 2001 Jun;69(6):4180–4.
207. Young RS, Fortney K, Haley JC, Hood AF, Campagnari AA, Wang J, et al. Expression of sialylated or paragloboside-like lipooligosaccharides are not required for pustule formation by Haemophilus ducreyi in human volunteers. Infect Immun. 1999 Dec;67(12):6335–40.
208. Throm RE, Al-Tawfiq JA, Fortney KR, Katz BP, Hood AF, Slaughter CA, et al. Evaluation of an isogenic major outer membrane protein-deficient mutant in the human model of Haemophilus ducreyi infection. Infect Immun. 2000 May;68(5):2602–7.
209. Thornton AC, O'Mara EM, Sorensen SJ, Hiltke TJ, Fortney K, Katz B, et al. Prevention of experimental Haemophilus ducreyi infection: a randomized, controlled clinical trial. J Infect Dis. 1998 Jun;177(6):1608–13.
210. Spinola SM, Fortney KR, Katz BP, Latimer JL, Mock JR, Vakevainen M, et al. Haemophilus ducreyi requires an intact flp gene cluster for virulence in humans. Infect Immun. 2003 Dec;71(12):7178–82.
211. Spinola SM, Wild LM, Apicella MA, Gaspari AA, Campagnari AA. Experimental human infection with Haemophilus ducreyi. J Infect Dis. 1994 May;169(5):1146–50.
212. Palmer KL, Thornton AC, Fortney KR, Hood AF, Munson RS, Spinola SM. Evaluation of an isogenic hemolysin-deficient mutant in the human model of Haemophilus ducreyi infection. J Infect Dis. 1998 Jul;178(1):191–9.
213. Leduc I, Banks KE, Fortney KR, Patterson KB, Billings SD, Katz BP, et al. Evaluation of the repertoire of the TonB-dependent receptors of Haemophilus ducreyi for their role in virulence in humans. J Infect Dis. 2008 Apr 15;197(8):1103–9.
214. Janowicz DM, Tenner-Racz K, Racz P, Humphreys TL, Schnizlein-Bick C, Fortney KR, et al. Experimental infection with Haemophilus ducreyi in persons who are infected with HIV does not cause local or augment systemic viral replication. J Infect Dis. 2007 May 15;195(10):1443–51.
215. Janowicz D, Luke NR, Fortney KR, Katz BP, Campagnari AA, Spinola SM. Expression of OmpP2A and OmpP2B is not required for pustule formation by Haemophilus ducreyi in human volunteers. Microb Pathog. 2006 Mar;40(3):110–5.

216. Janowicz D, Leduc I, Fortney KR, Katz BP, Elkins C, Spinola SM. A DltA mutant of Haemophilus ducreyi Is partially attenuated in its ability to cause pustules in human volunteers. Infect Immun. 2006 Feb;74(2):1394–7.
217. Janowicz DM, Fortney KR, Katz BP, Latimer JL, Deng K, Hansen EJ, et al. Expression of the LspA1 and LspA2 proteins by Haemophilus ducreyi is required for virulence in human volunteers. Infect Immun. 2004 Aug;72(8):4528–33.
218. Humphreys TL, Li L, Li X, Janowicz DM, Fortney KR, Zhao Q, et al. Dysregulated immune profiles for skin and dendritic cells are associated with increased host susceptibility to Haemophilus ducreyi infection in human volunteers. Infect Immun. 2007 Dec;75(12):5686–97.
219. Fulcher RA, Cole LE, Janowicz DM, Toffer KL, Fortney KR, Katz BP, et al. Expression of Haemophilus ducreyi collagen binding outer membrane protein NcaA is required for virulence in swine and human challenge models of chancroid. Infect Immun. 2006 May;74(5):2651–8.
220. Fortney KR, Young RS, Bauer ME, Katz BP, Hood AF, Munson RS, et al. Expression of peptidoglycan-associated lipoprotein is required for virulence in the human model of Haemophilus ducreyi infection. Infect Immun. 2000 Nov;68(11):6441–8.
221. Bong CTH, Fortney KR, Katz BP, Hood AF, San Mateo LR, Kawula TH, et al. A superoxide dismutase C mutant of Haemophilus ducreyi is virulent in human volunteers. Infect Immun. 2002 Mar;70(3):1367–71.
222. Bong CT, Throm RE, Fortney KR, Katz BP, Hood AF, Elkins C, et al. DsrA-deficient mutant of Haemophilus ducreyi is impaired in its ability to infect human volunteers. Infect Immun. 2001 Mar;69(3):1488–91.
223. Bauer ME, Townsend CA, Doster RS, Fortney KR, Zwickl BW, Katz BP, et al. A fibrinogen-binding lipoprotein contributes to the virulence of Haemophilus ducreyi in humans. J Infect Dis. 2009 Mar 1;199(5):684–92.
224. Banks KE, Fortney KR, Baker B, Billings SD, Katz BP, Munson RS, et al. The enterobacterial common antigen-like gene cluster of Haemophilus ducreyi contributes to virulence in humans. J Infect Dis. 2008 Jun 1;197(11):1531–6.
225. Al-Tawfiq JA, Fortney KR, Katz BP, Hood AF, Elkins C, Spinola SM. An isogenic hemoglobin receptor-deficient mutant of Haemophilus ducreyi is attenuated in the human model of experimental infection. J Infect Dis. 2000 Mar;181(3):1049–54.
226. Al-Tawfiq JA, Bauer ME, Fortney KR, Katz BP, Hood AF, Ketterer M, et al. A pilus-deficient mutant of Haemophilus ducreyi is virulent in the human model of experimental infection. J Infect Dis. 2000 Mar;181(3):1176–9.
227. Al-Tawfiq JA, Palmer KL, Chen CY, Haley JC, Katz BP, Hood AF, et al. Experimental infection of human volunteers with Haemophilus ducreyi does not confer protection against subsequent challenge. J Infect Dis. 1999 May;179(5):1283–7.
228. Al-Tawfiq JA, Thornton AC, Katz BP, Fortney KR, Todd KD, Hood AF, et al. Standardization of the experimental model of Haemophilus ducreyi infection in human subjects. J Infect Dis. 1998 Dec;178(6):1684–7.
229. Hodgson SH, Juma E, Salim A, Magiri C, Kimani D, Njenga D, et al. Evaluating controlled human malaria infection in Kenyan adults with varying degrees of prior exposure to Plasmodium falciparum using sporozoites administered by intramuscular injection. Front Microbiol. 2014;5:686.

230. Hodgson SH, Ewer KJ, Bliss CM, Edwards NJ, Rampling T, Anagnostou NA, et al. Evaluation of the efficacy of ChAd63-MVA vectored vaccines expressing circumsporozoite protein and ME-TRAP against controlled human malaria infection in malaria-naive individuals. J Infect Dis. 2015 Apr 1;211(7):1076–86.
231. Herrera S, Solarte Y, Jordán-Villegas A, Echavarría JF, Rocha L, Palacios R, et al. Consistent safety and infectivity in sporozoite challenge model of Plasmodium vivax in malaria-naive human volunteers. Am J Trop Med Hyg. 2011 Feb;84(2 Suppl):4–11.
232. Healy SA, Murphy SC, Hume JCC, Shelton L, Kuntz S, Van Voorhis WC, et al. Chemoprophylaxis Vaccination: Phase I Study to Explore Stage-specific Immunity to Plasmodium falciparum in US Adults. Clin Infect Dis. 2020 Sep 12;71(6):1481–90.
233. Harro C, Chakraborty S, Feller A, DeNearing B, Cage A, Ram M, et al. Refinement of a human challenge model for evaluation of enterotoxigenic Escherichia coli vaccines. Clin Vaccine Immunol. 2011 Oct;18(10):1719–27.
234. Harro C, Louis Bourgeois A, Sack D, Walker R, DeNearing B, Brubaker J, et al. Live attenuated enterotoxigenic Escherichia coli (ETEC) vaccine with dmLT adjuvant protects human volunteers against virulent experimental ETEC challenge. Vaccine. 2019 Mar 28;37(14):1978–86.
235. Harris SA, Meyer J, Satti I, Marsay L, Poulton ID, Tanner R, et al. Evaluation of a human BCG challenge model to assess antimycobacterial immunity induced by BCG and a candidate tuberculosis vaccine, MVA85A, alone and in combination. J Infect Dis. 2014 Apr 15;209(8):1259–68.
236. Hales C, Jochems SP, Robinson R, Solórzano C, Carniel B, Pojar S, et al. Symptoms associated with influenza vaccination and experimental human pneumococcal colonisation of the nasopharynx. Vaccine. 2020 Feb 28;38(10):2298–306.
237. Gómez-Pérez GP, Legarda A, Muñoz J, Sim BKL, Ballester MR, Dobaño C, et al. Controlled human malaria infection by intramuscular and direct venous inoculation of cryopreserved Plasmodium falciparum sporozoites in malaria-naïve volunteers: effect of injection volume and dose on infectivity rates. Malar J. 2015 Aug 7;14:306.
238. Gibani MM, Jones E, Barton A, Jin C, Meek J, Camara S, et al. Investigation of the role of typhoid toxin in acute typhoid fever in a human challenge model. Nat Med. 2019 Jul;25(7):1082–8.
239. Gibani MM, Jin C, Shrestha S, Moore M, Norman L, Voysey M, et al. Homologous and heterologous re-challenge with Salmonella Typhi and Salmonella Paratyphi A in a randomised controlled human infection model. PLoS Negl Trop Dis. 2020 Oct;14(10):e0008783.
240. Fullen DJ, Murray B, Mori J, Catchpole A, Borley DW, Murray EJ, et al. A Tool for Investigating Asthma and COPD Exacerbations: A Newly Manufactured and Well Characterised GMP Wild-Type Human Rhinovirus for Use in the Human Viral Challenge Model. PLoS One. 2016;11(12):e0166113.

241. Fullen DJ, Noulin N, Catchpole A, Fathi H, Murray EJ, Mann A, et al. Accelerating Influenza Research: Vaccines, Antivirals, Immunomodulators and Monoclonal Antibodies. The Manufacture of a New Wild-Type H3N2 Virus for the Human Viral Challenge Model. PLoS One. 2016;11(1):e0145902.
242. Ferreira DM, Neill DR, Bangert M, Gritzfeld JF, Green N, Wright AKA, et al. Controlled human infection and rechallenge with Streptococcus pneumoniae reveals the protective efficacy of carriage in healthy adults. Am J Respir Crit Care Med. 2013 Apr 15;187(8):855–64.
243. Eiden J, Volckaert B, Rudenko O, Ellis R, Aitchison R, Herber R, et al. 2748. Single Intranasal (IN) Dose of M2SR (M2-Deficient Single Replication) Live Influenza Vaccine Protects Adults Against Subsequent Challenge with a Substantially Drifted H3N2 Strain. Open Forum Infect Dis. 2019 Oct 23;6(Suppl 2):S967–8.
244. DeVincenzo J, Tait D, Efthimiou J, Mori J, Kim Y-I, Thomas E, et al. A Randomized, Placebo-Controlled, Respiratory Syncytial Virus Human Challenge Study of the Antiviral Efficacy, Safety, and Pharmacokinetics of RV521, an Inhibitor of the RSV-F Protein. Antimicrob Agents Chemother. 2020 Jan 27;64(2):e01884-19.
245. de Graaf H, Ibrahim M, Hill AR, Gbesemete D, Vaughan AT, Gorringe A, et al. Controlled Human Infection With Bordetella pertussis Induces Asymptomatic, Immunizing Colonization. Clin Infect Dis. 2020 Jul 11;71(2):403–11.
246. Darton TC, Jones C, Blohmke CJ, Waddington CS, Zhou L, Peters A, et al. Using a Human Challenge Model of Infection to Measure Vaccine Efficacy: A Randomised, Controlled Trial Comparing the Typhoid Vaccines M01ZH09 with Placebo and Ty21a. PLoS Negl Trop Dis. 2016 Aug;10(8):e0004926.
247. Darsley MJ, Chakraborty S, DeNearing B, Sack DA, Feller A, Buchwaldt C, et al. The oral, live attenuated enterotoxigenic Escherichia coli vaccine ACE527 reduces the incidence and severity of diarrhea in a human challenge model of diarrheal disease. Clin Vaccine Immunol. 2012 Dec;19(12):1921–31.
248. Collins KA, Wang CY, Adams M, Mitchell H, Rampton M, Elliott S, et al. A controlled human malaria infection model enabling evaluation of transmission-blocking interventions. J Clin Invest. 2018 Apr 2;128(4):1551–62.
249. Chakraborty S, Harro C, DeNearing B, Brubaker J, Connor S, Maier N, et al. Impact of lower challenge doses of enterotoxigenic Escherichia coli on clinical outcome, intestinal colonization and immune responses in adult volunteers. PLoS Negl Trop Dis. 2018 Apr;12(4):e0006442.
250. Bodhidatta L, Pitisuttithum P, Chamnanchanant S, Chang KT, Islam D, Bussaratid V, et al. Establishment of a Shigella sonnei human challenge model in Thailand. Vaccine. 2012 Nov 19;30(49):7040–5.
251. Bijker EM, Bastiaens GJH, Teirlinck AC, van Gemert G-J, Graumans W, van de Vegte-Bolmer M, et al. Protection against malaria after immunization by chloroquine prophylaxis and sporozoites is mediated by preerythrocytic immunity. Proc Natl Acad Sci U S A. 2013 May 7;110(19):7862–7.

252. Bijker EM, Teirlinck AC, Schats R, van Gemert G-J, van de Vegte-Bolmer M, van Lieshout L, et al. Cytotoxic markers associate with protection against malaria in human volunteers immunized with Plasmodium falciparum sporozoites. J Infect Dis. 2014 Nov 15;210(10):1605–15.
253. Bastiaens GJH, van Meer MPA, Scholzen A, Obiero JM, Vatanshenassan M, van Grinsven T, et al. Safety, Immunogenicity, and Protective Efficacy of Intradermal Immunization with Aseptic, Purified, Cryopreserved Plasmodium falciparum Sporozoites in Volunteers Under Chloroquine Prophylaxis: A Randomized Controlled Trial. Am J Trop Med Hyg. 2016 Mar;94(3):663–73.
254. Adler H, German EL, Mitsi E, Nikolaou E, Pojar S, Hales C, et al. Experimental Human Pneumococcal Colonization in Older Adults Is Feasible and Safe, Not Immunogenic. Am J Respir Crit Care Med. 2021 Mar 1;203(5):604–13.
255. Achan J, Reuling IJ, Yap XZ, Dabira E, Ahmad A, Cox M, et al. Serologic Markers of Previous Malaria Exposure and Functional Antibodies Inhibiting Parasite Growth Are Associated With Parasite Kinetics Following a Plasmodium falciparum Controlled Human Infection. Clin Infect Dis. 2020 Jun 10;70(12):2544–52.
256. Walk J, de Bree LCJ, Graumans W, Stoter R, van Gemert G-J, van de Vegte-Bolmer M, et al. Outcomes of controlled human malaria infection after BCG vaccination. Nat Commun. 2019 Feb 20;10(1):874.
257. Stevens M, Rusch S, DeVincenzo J, Kim Y-I, Harrison L, Meals EA, et al. Antiviral Activity of Oral JNJ-53718678 in Healthy Adult Volunteers Challenged With Respiratory Syncytial Virus: A Placebo-Controlled Study. J Infect Dis. 2018 Jul 24;218(5):748–56.
258. Taylor DN, McKenzie R, Durbin A, Carpenter C, Atzinger CB, Haake R, et al. Rifaximin, a nonabsorbed oral antibiotic, prevents shigellosis after experimental challenge. Clin Infect Dis. 2006 May 1;42(9):1283–8.
259. Kotloff KL, Losonsky GA, Nataro JP, Wasserman SS, Hale TL, Taylor DN, et al. Evaluation of the safety, immunogenicity, and efficacy in healthy adults of four doses of live oral hybrid Escherichia coli-Shigella flexneri 2a vaccine strain EcSf2a-2. Vaccine. 1995 Apr;13(5):495–502.
260. Coster TS, Hoge CW, VanDeVerg LL, Hartman AB, Oaks EV, Venkatesan MM, et al. Vaccination against shigellosis with attenuated Shigella flexneri 2a strain SC602. Infect Immun. 1999 Jul;67(7):3437–43.
261. Malcolm BA, Aerts CA, Dubois KJ, Geurts FJ, Marien K, Rusch S, et al. PrEP-001 prophylactic effect against rhinovirus and influenza virus - RESULTS of 2 randomized trials. Antiviral Res. 2018 May;153:70–7.
262. Hickey BW, Lumsden JM, Reyes S, Sedegah M, Hollingdale MR, Freilich DA, et al. Mosquito bite immunization with radiation-attenuated Plasmodium falciparum sporozoites: safety, tolerability, protective efficacy and humoral immunogenicity. Malar J. 2016 Jul 22;15(1):377.
263. Lyke KE, Ishizuka AS, Berry AA, Chakravarty S, DeZure A, Enama ME, et al. Attenuated PfSPZ Vaccine induces strain-transcending T cells and durable protection against heterologous controlled human malaria infection. Proc Natl Acad Sci U S A. 2017 Mar 7;114(10):2711–6.

264. Lyke KE, Laurens MB, Strauss K, Adams M, Billingsley PF, James E, et al. Optimizing Intradermal Administration of Cryopreserved Plasmodium falciparum Sporozoites in Controlled Human Malaria Infection. Am J Trop Med Hyg. 2015 Dec;93(6):1274–84.
265. Ishizuka AS, Lyke KE, DeZure A, Berry AA, Richie TL, Mendoza FH, et al. Protection against malaria at 1 year and immune correlates following PfSPZ vaccination. Nat Med. 2016 Jun;22(6):614–23.
266. Hoffman SL, Edelman R, Bryan JP, Schneider I, Davis J, Sedegah M, et al. Safety, immunogenicity, and efficacy of a malaria sporozoite vaccine administered with monophosphoryl lipid A, cell wall skeleton of mycobacteria, and squalane as adjuvant. Am J Trop Med Hyg. 1994 Nov;51(5):603–12.
267. Herrington D, Davis J, Nardin E, Beier M, Cortese J, Eddy H, et al. Successful immunization of humans with irradiated malaria sporozoites: humoral and cellular responses of the protected individuals. Am J Trop Med Hyg. 1991 Nov;45(5):539–47.
268. Herrington DA, Clyde DF, Davis JR, Baqar S, Murphy JR, Cortese JF, et al. Human studies with synthetic peptide sporozoite vaccine (NANP)3-TT and immunization with irradiated sporozoites. Bull World Health Organ. 1990;68 Suppl:33–7.
269. Herrington DA, Clyde DF, Losonsky G, Cortesia M, Murphy JR, Davis J, et al. Safety and immunogenicity in man of a synthetic peptide malaria vaccine against Plasmodium falciparum sporozoites. Nature. 1987 Jul 16;328(6127):257–9.
270. Epstein JE, Tewari K, Lyke KE, Sim BKL, Billingsley PF, Laurens MB, et al. Live attenuated malaria vaccine designed to protect through hepatic CD8+ T cell immunity. Science. 2011 Oct 28;334(6055):475–80.
271. Edelman R, Hoffman SL, Davis JR, Beier M, Sztein MB, Losonsky G, et al. Long-term persistence of sterile immunity in a volunteer immunized with X-irradiated Plasmodium falciparum sporozoites. J Infect Dis. 1993 Oct;168(4):1066–70.
272. Davis JR, Cortese JF, Herrington DA, Murphy JR, Clyde DF, Thomas AW, et al. Plasmodium falciparum: in vitro characterization and human infectivity of a cloned line. Exp Parasitol. 1992 Mar;74(2):159–68.
273. DeVincenzo JP, Whitley RJ, Mackman RL, Scaglioni-Weinlich C, Harrison L, Farrell E, et al. Oral GS-5806 activity in a respiratory syncytial virus challenge study. N Engl J Med. 2014 Aug 21;371(8):711–22.
274. DeVincenzo JP, McClure MW, Symons JA, Fathi H, Westland C, Chanda S, et al. Activity of Oral ALS-008176 in a Respiratory Syncytial Virus Challenge Study. N Engl J Med. 2015 Nov 19;373(21):2048–58.
275. Chuang I, Sedegah M, Cicatelli S, Spring M, Polhemus M, Tamminga C, et al. DNA prime/Adenovirus boost malaria vaccine encoding P. falciparum CSP and AMA1 induces sterile protection associated with cell-mediated immunity. PLoS One. 2013;8(2):e55571.
276. Chen WH, Cohen MB, Kirkpatrick BD, Brady RC, Galloway D, Gurwith M, et al. Single-dose Live Oral Cholera Vaccine CVD 103-HgR Protects Against Human Experimental Infection With Vibrio cholerae O1 El Tor. Clin Infect Dis. 2016 Jun 1;62(11):1329–35.

### Appendix C - Data Categorization

The following tags were used to categorize data from each study:

- Challenge: describes a traditional challenge trial
- Prechallenge: describes an immunization phase or a vaccination phase that occurred prior to a traditional challenge phase
- Rechallenge: describes a cohort of volunteers that have been previously exposed to a challenge agent (in a previous study or in an earlier challenge phase of the current study), that were then challenged again (either with the same strain or a different strain of the challenge agent)
- Attenuated: describes a challenge with an attenuated challenge agent
- Vaccine: describes a challenge with a vaccine
- Summed: previously extracted data from multiple phases, studies, or cohorts that was summed to be treated as a single study
- NAE: studies where AEs were not mentioned/defined, and other reported data (symptoms, illness, etc.) was extracted instead
- NSAE: studies where SAEs were not mentioned in text, and the current FDA definition of SAEs was manually applied to reported symptom/illness data in order to identify possible SAEs
- Excluded: studies that were excluded for any reason

### Appendix D - Supplementary Tables

Also linked here: Systematic Review - Tables.

##### Supplementary Table 1 - Reasons for Exclusion

| Reasons for exclusion | Excluded during screening, n | Excluded during full text review, n |
|---|---|---|
| Not challenge trials | 1053 | 28 |
| Review, guidelines, editorials, or comments | 974 | 2 |
| Animal studies | 327 | 0 |
| Not in English | 38 | 1 |

| | | |
|---|---|---|
| Case report | 21 | 1 |
| Duplicate | 1 | 47 |
| Pre-1980s | 0 | 30 |
| Uses previously published data | 0 | 26 |
| Unpublished data | 0 | 6 |
| Unable to locate or access (full text not available) | 0 | 5 |
| Insufficient data | 0 | 1 |
| Describes protocol for a future study | 0 | 1 |
| Total | 2414 | 148 |

**Supplementary Table 2 - Reasons for Exclusion during Full-Text Review (By Method of Identification)**

| Reasons for exclusion | Excluded (search results), n | Excluded (other reports), n |
|---|---|---|
| Not challenge trials | 26 | 0 |
| Duplicates | 19 | 28 |
| Uses previously published data | 18 | 8 |
| Pre-1980s | 14 | 16 |
| Unpublished data | 1 | 5 |
| Unable to locate/access (no full text available) | 1 | 4 |
| Describes protocol for future study | 1 | 0 |
| Reviews | 0 | 2 |
| Not challenge trials | 0 | 2 |
| Case reports | 0 | 1 |
| Insufficient data | 0 | 1 |

| | | |
|---|---|---|
| Not in English | 0 | 1 |
| Total | 80 | 68 |

**Supplementary Table 3 - Number of Studies, Number of Participants, Number of Infections, Data Reporting, and Database Registration in Published HCTs by Decade (Rechallenges) (Generated 2/14/22, Excluded Tags: Challenge, Prechallenge, Excluded)**

| Decade | Studies, n | Participants challenged, n | Challenged participants diagnosed with infection, n | Studies that do not define AEs, n (%) | Studies with unclear AE data, n (%) | Studies with no AE data, n (%) | Studies that do not mention SAEs, n (%) | Studies with no SAE data, n (%) | Studies registered in a database, n (%) |
|---|---|---|---|---|---|---|---|---|---|
| **1980s** | 2 | 28 | 15 | 2 (100.0) | 0 (0.0) | 0 (0.0) | 2 (100.0) | 0 (0.0) | 0 (0.0) |
| **1990s** | 9 | 69 | 54-55a | 8 (88.9) | 0 (0.0) | 8 (88.9) | 1 (11.1) | 8 (88.9) | 0 (0.0) |
| **2000s** | 3 | 32 | 20 | 3 (100.0) | 0 (0.0) | 3 (100.0) | 0 (0.0) | 3 (100.0) | 0 (0.0) |
| **2010s** | 8 | 203 | 89 | 1 (12.5) | 1 (12.5) | 4 (50.0) | 1 (12.5) | 6 (75.0) | 6 (75.0) |
| **2020s** | 2 | 86 | 40 | 1 (50.0) | 2 (100.) | 0 (0.0) | 0 (0.0) | 0 (0.0) | 2 (100.0) |
| **Total** | 24 | 418 | 218-219a | 15 (62.5) | 3 (12.5) | 15 (62.5) | 4 (16.7) | 17 (70.8) | 8 (33.3) |

a) A range of values is given to account for unclear data reporting by one study.

**Supplementary Table 4 - Adverse Events in Published HCTs by Decade (Rechallenges) (Excluded Tags: Challenge, Prechallenge, Excluded)**

| Decade | Studiesa, n | Participants challenged, n | Challenged participants with AEs (minimumb), n (%) | Challenged participants with AEs (maximumb), n (%) |
|---|---|---|---|---|
| **1980s** | 2 | 28 | 2 (7.1) | 2 (7.1) |
| **1990s** | 1 | 13 | 0 (0.0) | 0 (0.0) |
| **2000s** | 0 | 0 | 0 (0.0) | 0 (0.0) |

| | | | | |
|---|---|---|---|---|
| **2010s** | 4 | 35 | 17 (48.6) | 20 (57.1) |
| **2020s** | 2 | 86 | 8 (9.3) | 86 (100.0) |
| **Total** | 9 | 162 | 27 (16.7) | 108 (66.7) |

a) 15 studies that did not report AE data are excluded.

b) Minimum and maximum values are given to account for unclear data reporting by some studies.

#### Supplementary Table 5 - Severe Adverse Events in Published HCTs by Decade (Rechallenges) (Excluded Tags: Challenge, Prechallenge, Excluded)

| Decade | Studiesa, n | Participants challenged, n | Challenged participants with severe or very severe (≥grade 3) AEs (minimumb), n (%) | Challenged participants with severe or very severe (≥grade 3) AEs (maximumb), n (%) |
|---|---|---|---|---|
| **1980s** | 0 | 0 | 0 (0.0) | 0 (0.0) |
| **1990s** | 0 | 0 | 0 (0.0) | 0 (0.0) |
| **2000s** | 0 | 0 | 0 (0.0) | 0 (0.0) |
| **2010s** | 3 | 31 | 5 (16.1) | 14 (45.2) |
| **2020s** | 0 | 0 | 0 (0.0) | 0 (0.0) |
| **Total** | 3 | 31 | 5 (16.1) | 14 (45.2) |

a) 21 studies that did not report severe AE data are excluded.

b) Minimum and maximum values are given to account for unclear data reporting by some studies.

#### Supplementary Table 6 - Serious Adverse Events in Published HCTs by Decade (Rechallenges) (Excluded Tags: Challenge, Prechallenge, Excluded)

| Decade | Studiesa, n | Participants challenged, n | Challenged participants with SAEs, n (%) |
|---|---|---|---|

| | | | |
|---|---|---|---|
| **1980s** | 2 | 28 | 0 (0.0) |
| **1990s** | 1 | 13 | 0 (0.0) |
| **2000s** | 0 | 0 | 0 (0.0) |
| **2010s** | 2 | 19 | 0 (0.0) |
| **2020s** | 2 | 86 | 1 (1.2) |
| **Total** | 7 | 146 | 1 (0.7) |

a) 17 studies that did not report SAE data are excluded.

**Supplementary Table 7 - Descriptions of Additional Serious Adverse Events (Not Related to Challenge) in Published HCTs by Pathogen Category**

| Pathogen category | Participants with ≥1 SAE, n | Description | Outcomes | File name |
|---|---|---|---|---|
| **Escherichia coli** | | | | |
| | 1 | "There were no serious AEs following vaccination, but one occurred in a placebo recipient during long-term follow-up postchallenge. This event was not considered to be related to participation in the study." | ND | Darsley 2012 |
| **Neisseria lactamica** | | | | |
| | 4 | "Four participants experienced serious adverse events but none were related to the study." | ND | Deasy 2015 |
| **Norovirus** | | | | |
| | 1 | "There was 1 serious adverse event during the study and it was assessed as unrelated. A placebo recipient developed a partial retinal detachment 74 days after receipt of study | ND | Atmar 2014 |

| | | | | |
|---|---|---|---|---|
| | | product." | | |
| **Plasmodium spp.** | | | | |
| | 1 | "All volunteers successfully recovered clinically within 2–3 days without any serious AE. None of the volunteers required hospitalization for malaria. A volunteer presented with severe diarrhea and abdominal pain (Grade 4) on day 18 and was diagnosed with parasitic gastroenteritis (hookworms and trichuriasis) and was held under observation for <24 hours in the hospital, requiring intravenous rehydration and pain management." | ND | Arévalo-Herrera 2014 |
| | 1 | "One SAE (ductal carcinoma in situ of the breast) occurred during the study and it was determined that the event was not related to the study vaccine or CHMI." | ND | Bennett 2016 |
| | 1 | "The only serious adverse event (recurrent tonsillitis) was considered unrelated to the clinical trial treatment." | ND | McCarthy 2016a |
| | 1 | "One subject was diagnosed with colon cancer and did not participate in CHMI." | ND | Seder 2013 |
| | 1 | "After CHMI, one vaccine recipient developed anxiety and headache 14 days after completing chloroquine treatment and was hospitalized overnight for diagnostic purposes." | "The symptoms, which were attributed to chloroquine, resolved within 1 month." | Seder 2013 |
| | 1 | "One adverse event (renal colic) was classified as serious; however, this event was determined not to be related to study treatments." | "All adverse events had resolved by the end of study." | Smith 2017 |
| | 1 | "During follow-up, one serious adverse event occurred in cohort 1A: a bilateral pulmonary embolism occurred in a non-smoking woman under oral contraceptive treatment 34 days after DSM265 administration while she was on a transatlantic flight." | "The volunteer never became parasitaemic and recovered fully, except for a grade 1 sinus tachycardia, which still persisted at the last follow-up visit." | Sulyok 2017 |
| **Rhinovirus** | | | | |

| | | | | |
|---|---|---|---|---|
| | 1 | "One serious adverse event (SAE) was reported in the 1 TCID50 group. The SAE was a Grade 4 self-limiting raised creatine phosphokinase considered to be of medical importance, but not considered to be related to the Challenge Virus, study procedures, concomitant medications or study assessments." | "No action was taken and the SAE resolved." | Fullen 2016b |
| **Salmonella spp.** | | | | |
| | 1 | "One serious adverse event was recorded (appendicitis 6 months after S. Paratyphi A challenge) and was assessed as being unrelated to study procedures." | ND | Dobinson 2017 |
| | 1 | "Generalised tonic clonic seizure": "Occurred 9 months after completion of challenge. Reported a history of childhood seizures that was not disclosed at screening or on available medical records. Commenced on anticonvulsants and ongoing follow up with neurology at end of study" | ND | Gibani 2019 |
| | 1 | "Deliberate self harm": "Episode of deliberate self-harm 6 months after completion of challenge, attributed to an acute life stressor. No specific treatment was administered." | ND | Gibani 2019 |
| | 1 | "Episode of collapse with loss of consciousness 12 months after challenge. Reported left arm and left leg weakness 24 hours' duration. Diagnosed as possible generalised seizure following neurology review. Instructed not to drive for 6/12." | "No medication prescribed, and no subsequent events." | Gibani 2020 |
| | 1 | "Right loin pain with evidence of renal calculus on CT KUB. Diagnosed after enrolment but prior to challenge. Withdrawn from study and not challenged." | ND | Gibani 2020 |
| | 1 | "One Vi-TT participant was diagnosed with inflammatory bowel disease and withdrawn from the study (symptoms preceded study enrolment)" | ND | Jin 2017 |
| | 2 | "two Vi-PS participants were hospitalised (urinary retention and semi-elective tonsillectomy)" | ND | Jin 2017 |

| | | | | |
|---|---|---|---|---|
| **Shigella spp.** | | | | |
| | 1 | "In the Placebo group, one participant experienced four SAEs following the challenge dose: deep vein thrombosis (moderate intensity) and pelvic venous thrombosis (severe) at day 107, haematoma (severe) at day 121, and carotid artery aneurysm at day 130 from challenge." | "All events were assessed by the investigator as unrelated to any study treatment and were resolved." | Frenck 2021 |

**Supplementary Table 8 - Number of Published HCTS, Number of Participants, Number Infected, and Number with Serious Adverse Events by Pathogen Category (Rechallenges) (FINAL - Generated 2/22/22, Excluded Tags: Challenge, Prechallenge, Excluded)**

| Pathogen category | Studies, n | Participants challenged across all studies, n | Challenged participants diagnosed with infection across all studies, n (%) | Studies that reported SAEs, n | Participants challenged across studies that reported SAEs, n | Challenged participants with SAEs across studies that reported SAEs, n (%) |
|---|---|---|---|---|---|---|
| **Campylobacter jejuni** | 2 | 24 | 18 (75.0) | 2 | 24 | 0 (0.0) |
| **Escherichia coli** | 4 | 40 | 33 (82.5) | 1 | 11 | 0 (0.0) |
| **Haemophilus ducreyi** | 5 | 54 | 27 (50.0) | 0 | 0 | 0 (0.0) |
| **Influenza viruses** | 2 | 32 | 16 (50.0) | 2 | 32 | 0 (0.0) |
| **Necator americanus** | 2 | 17 | 14 (82.4) | 0 | 0 | 0 (0.0) |
| **Neisseria lactamica** | 1 | 127 | 44 (34.6) | 0 | 0 | 0 (0.0) |
| **Plasmodium spp.** | 6 | 43 | 13 (30.2) | 1 | 4 | 0 (0.0) |

| | | | | | | |
|---|---|---|---|---|---|---|
| **Salmonella spp.** | 1 | 75 | 35 (46.7) | 1 | 75 | 1 (1.3) |
| **Shigella spp.** | 1 | 6 | 1 (16.7) | 0 | 0 | 0 (0.0) |
| **Total** | 24 | 418 | 201 (48.1) | 7 | 146 | 1 (0.7) |

##### Supplementary Table 9 - Adverse Events in Published HCTs by Decade (Excluding manual AE counts) (Excluded Tags: Prechallenge, Rechallenge, NAE, Excluded)

| Decade | Studiesa, n | Participants challenged, n | Challenged participants with AEs (minimumb), n (%) | Challenged participants with AEs (maximumb), n (%) |
|---|---|---|---|---|
| **1980s** | 0 | 0 | 0 (0.0) | 0 (0.0) |
| **1990s** | 3 | 518 | 136 (26.3) | 153 (29.5) |
| **2000s** | 16 | 1627 | 524 (32.2) | 768 (47.2) |
| **2010s** | 71 | 2697 | 1324 (49.1) | 1939 (71.9) |
| **2020s** | 20 | 879 | 417 (47.4) | 707 (80.4) |
| **Total** | 110 | 5721 | 2401 (42.0) | 3567 (62.3) |

a) 93 studies that did not report AE data are excluded.

b) Minimum and maximum values are given to account for unclear data reporting by some studies.

##### Supplementary Table 10 - Serious Adverse Events in Published HCTs by Decade (Excluding manual SAE counts) (Excluded Tags: Prechallenge, Rechallenge, NSAE, Excluded)

| Decade | Studiesa, n | Challenged participants, n | Challenged participants with SAEs, n (%) |
|---|---|---|---|
| **1980s** | 2 | 17 | 0 (0.0) |

| | | | |
|---|---|---|---|
| **1990s** | 1 | 15 | 0 (0.0) |
| **2000s** | 14 | 796 | 1 (0.1) |
| **2010s** | 65 | 2832 | 13 (0.5) |
| **2020s** | 21 | 931 | 3 (0.3) |
| **Total** | 103 | 4591 | 17 (0.4) |

a) 97 studies that did not report SAE data are excluded.

### Appendix E - Dataset

The full dataset, including the code used for generating tables and the output tables, will be available online here. The dataset can be used to generate the tables and figures in the paper, but care should be taken to properly include/exclude studies in the interface.

### Appendix F - PRISMA Checklist

| Section and Topic | Item # | Checklist item | Location where item is reported |
|---|---|---|---|
| **TITLE** | | | |
| Title | 1 | Identify the report as a systematic review. | Title |
| **ABSTRACT** | | | |
| Abstract | 2 | See the PRISMA 2020 for Abstracts checklist. | Abstract |
| **INTRODUCTION** | | | |
| Rationale | 3 | Describe the rationale for the review in the context of existing knowledge. | Introduction |
| Objectives | 4 | Provide an explicit statement of the objective(s) or question(s) the review addresses. | Introduction |
| **METHODS** | | | |
| Eligibility criteria | 5 | Specify the inclusion and exclusion criteria for the review and how studies were grouped for the syntheses. | Sec 2.4-5 |
| Information sources | 6 | Specify all databases, registers, websites, organisations, reference lists and other sources searched or consulted to identify studies. Specify the date when each source was last searched or consulted. | Sec. 2.1 |
| Search strategy | 7 | Present the full search strategies for all databases, registers and websites, including any filters and limits used. | Sec. 2.1 |
| Selection process | 8 | Specify the methods used to decide whether a study met the inclusion criteria of the review, including how many reviewers screened each record and each report retrieved, whether they worked independently, and if applicable, details of automation tools used in the process. | Sec 2.1-4 |
| Data collection process | 9 | Specify the methods used to collect data from reports, including how many reviewers collected data from each report, whether they worked independently, any processes for obtaining or confirming data from study investigators, and if applicable, details of automation tools used in the process. | Sec 2.4 |
| Data items | 10a | List and define all outcomes for which data were sought. Specify whether all results that were compatible with each outcome domain in each study were sought (e.g. for all measures, time points, analyses), and if not, the methods used to decide which results to collect. | Sec 2.5 |
| | 10b | List and define all other variables for which data were sought (e.g. participant and intervention characteristics, funding sources). Describe any assumptions made about any missing or unclear information. | Sec 2.5 |
| Study risk of bias assessment | 11 | Specify the methods used to assess risk of bias in the included studies, including details of the tool(s) used, how many reviewers assessed each study and whether they worked independently, and if applicable, details of automation tools used in the process. | Note 1 |
| Effect measures | 12 | Specify for each outcome the effect measure(s) (e.g. risk ratio, mean difference) used in the synthesis or presentation of results. | N/A |
| Synthesis | 13a | Describe the processes used to decide which studies were eligible for each synthesis (e.g. tabulating the | N/A |

| | | | |
|---|---|---|---|
| methods | | study intervention characteristics and comparing against the planned groups for each synthesis (item #5)). | |
| | 13b | Describe any methods required to prepare the data for presentation or synthesis, such as handling of missing summary statistics, or data conversions. | N/A |
| | 13c | Describe any methods used to tabulate or visually display results of individual studies and syntheses. | N/A |
| | 13d | Describe any methods used to synthesize results and provide a rationale for the choice(s). If meta-analysis was performed, describe the model(s), method(s) to identify the presence and extent of statistical heterogeneity, and software package(s) used. | N/A |
| | 13e | Describe any methods used to explore possible causes of heterogeneity among study results (e.g. subgroup analysis, meta-regression). | N/A |
| | 13f | Describe any sensitivity analyses conducted to assess robustness of the synthesized results. | N/A |
| Reporting bias assessment | 14 | Describe any methods used to assess risk of bias due to missing results in a synthesis (arising from reporting biases). | Sec 2.6, 3.6 |
| Certainty assessment | 15 | Describe any methods used to assess certainty (or confidence) in the body of evidence for an outcome. | N/A |
| **RESULTS** | | | |
| Study selection | 16a | Describe the results of the search and selection process, from the number of records identified in the search to the number of studies included in the review, ideally using a flow diagram. | Sec. 3.1 |
| | 16b | Cite studies that might appear to meet the inclusion criteria, but which were excluded, and explain why they were excluded. | Appendix G (letter may change) |
| Study characteristics | 17 | Cite each included study and present its characteristics. | Appendix B |
| Risk of bias in studies | 18 | Present assessments of risk of bias for each included study. | N/A |
| Results of individual studies | 19 | For all outcomes, present, for each study: (a) summary statistics for each group (where appropriate) and (b) an effect estimate and its precision (e.g. confidence/credible interval), ideally using structured tables or plots. | Sec 3.2-6 |
| Results of syntheses | 20a | For each synthesis, briefly summarise the characteristics and risk of bias among contributing studies. | N/A |
| | 20b | Present results of all statistical syntheses conducted. If meta-analysis was done, present for each the summary estimate and its precision (e.g. confidence/credible interval) and measures of statistical heterogeneity. If comparing groups, describe the direction of the effect. | N/A |
| | 20c | Present results of all investigations of possible causes of heterogeneity among study results. | N/A |
| | 20d | Present results of all sensitivity analyses conducted to assess the robustness of the synthesized results. | N/A |
| Reporting biases | 21 | Present assessments of risk of bias due to missing results (arising from reporting biases) for each synthesis assessed. | Sec 3.6, N/A |
| Certainty of evidence | 22 | Present assessments of certainty (or confidence) in the body of evidence for each outcome assessed. | N/A |
| **DISCUSSION** | | | |
| Discussion | 23a | Provide a general interpretation of the results in the context of other evidence. | Sec 4 |

| | | | |
|---|---|---|---|
| | 23b | Discuss any limitations of the evidence included in the review. | Sec 4 |
| | 23c | Discuss any limitations of the review processes used. | Sec 2.4, 4 |
| | 23d | Discuss implications of the results for practice, policy, and future research. | Sec 4, 5 |
| **OTHER INFORMATION** | | | |
| Registration and protocol | 24a | Provide registration information for the review, including register name and registration number, or state that the review was not registered. | Sec 2.1 |
| | 24b | Indicate where the review protocol can be accessed, or state that a protocol was not prepared. | Appendix R |
| | 24c | Describe and explain any amendments to information provided at registration or in the protocol. | Amended Preregistration |
| Support | 25 | Describe sources of financial or non-financial support for the review, and the role of the funders or sponsors in the review. | Conflict of Interest Disclosures |
| Competing interests | 26 | Declare any competing interests of review authors. | Note |
| Availability of data, code and other materials | 27 | Report which of the following are publicly available and where they can be found: template data collection forms; data extracted from included studies; data used for all analyses; analytic code; any other materials used in the review. | Appendix E, R, |

*From:* Page MJ, McKenzie JE, Bossuyt PM, Boutron I, Hoffmann TC, Mulrow CD, et al. The PRISMA 2020 statement: an updated guideline for reporting systematic reviews. BMJ 2021;372:n71. doi: 10.1136/bmj.n71

Notes:

1) Risk of bias in the conduct of individual studies was not assessed because the relevant aim of the review was to review reported data, and thereby assess whether data reporting in challenge trials has been sufficient.

### Appendix G - Excluded Studies

Articles that described studies that performed secondary analysis of results from previously conducted HCTs, as well as articles that reported results from studies described in other articles which had already been included, were excluded.


1. Mitsi E, Roche AM, Reiné J, et al. Agglutination by anti-capsular polysaccharide antibody is associated with protection against experimental human pneumococcal carriage. Mucosal Immunol **2017**; 10:385–394.
2. Zhu J, Message SD, Qiu Y, et al. Airway Inflammation and Illness Severity in Response to Experimental Rhinovirus Infection in Asthma. CHEST **2014**; 145:1219–1229.
3. Medema GJ, Teunis PFM, Havelaar AH, Haas CN. Assessment of the dose-response relationship of Campylobacter jejuni. International Journal of Food Microbiology **1996**; 30:101–111.
4. van Wolfswinkel ME, Langenberg MCC, Wammes LJ, et al. Changes in total and differential leukocyte counts during the clinically silent liver phase in a controlled human malaria infection in malaria-naïve Dutch volunteers. Malaria Journal **2017**; 16:457.
5. Gelfanova V, Humphreys TL, Spinola SM. Characterization of Haemophilus ducreyi-Specific T-Cell Lines from Lesions of Experimentally Infected Human Subjects. Infection and Immunity **2001**; 69:4224–4231.
6. Porter CK, Lynen A, Riddle MS, et al. Clinical endpoints in the controlled human challenge model for Shigella: A call for standardization and the development of a disease severity score. PLOS ONE **2018**; 13:e0194325.
7. Church LWP, Le TP, Bryan JP, et al. Clinical Manifestations of Plasmodium falciparum Malaria Experimentally Induced by Mosquito Challenge. The Journal of Infectious Diseases **1997**; 175:915–920.
8. Roestenberg M, O'Hara GA, Duncan CJA, et al. Comparison of Clinical and Parasitological Data from Controlled Human Malaria Infection Trials. PLOS ONE **2012**; 7:e38434.
9. Walk J, Schats R, Langenberg MCC, et al. Diagnosis and treatment based on quantitative PCR after controlled human malaria infection. Malaria Journal **2016**; 15:398.
10. Ducarmon QR, Hoogerwerf MA, Janse JJ, et al. Dynamics of the bacterial gut microbiota during controlled human infection with Necator americanus larvae. Gut Microbes **2020**; 12:1840764.
11. Plaisance KI, Kudaravalli S, Wasserman SS, Levine MM, Mackowiak PA. Effect of Antipyretic Therapy on the Duration of Illness in Experimental Influenza A, Shigella sonnei, and Rickettsia rickettsii Infections. Pharmacotherapy: The Journal of Human Pharmacology and Drug Therapy **2000**; 20:1417–1422.

12. Turner RB, Cetnarowski WE. Effect of Treatment with Zinc Gluconate or Zinc Acetate on Experimental and Natural Colds. Clinical Infectious Diseases **2000**; 31:1202–1208.
13. Lamsfus Calle C, Fendel R, Singh A, et al. Expansion of Functional Myeloid-Derived Suppressor Cells in Controlled Human Malaria Infection. Frontiers in Immunology **2021**; 12:690.
14. Janowicz DM, Ofner S, Katz BP, Spinola SM. Experimental Infection of Human Volunteers with Haemophilus ducreyi: Fifteen Years of Clinical Data and Experience. The Journal of Infectious Diseases **2009**; 199:1671–1679.
15. Shimanovich AA, Buskirk AD, Heine SJ, et al. Functional and Antigen-Specific Serum Antibody Levels as Correlates of Protection against Shigellosis in a Controlled Human Challenge Study. Clinical and Vaccine Immunology 24:e00412-16.
16. Matsumiya M, Satti I, Chomka A, et al. Gene Expression and Cytokine Profile Correlate With Mycobacterial Growth in a Human BCG Challenge Model. The Journal of Infectious Diseases **2015**; 211:1499–1509.
17. Banks KE, Humphreys TL, Li W, Katz BP, Wilkes DS, Spinola SM. Haemophilus ducreyi Partially Activates Human Myeloid Dendritic Cells. Infection and Immunity **2007**; 75:5678–5685.
18. Doyle WJ, Alper CM, Buchman CA, Moody SA, Skoner DP, Cohen S. Illness and Otological Changes During Upper Respiratory Virus Infection. The Laryngoscope **1999**; 109:324–328.
19. Hodgson SH, Juma E, Salim A, et al. Lessons learnt from the first controlled human malaria infection study conducted in Nairobi, Kenya. Malaria Journal **2015**; 14:182.
20. Bauer ME, Spinola SM. Localization of Haemophilus ducreyi at the Pustular Stage of Disease in the Human Model of Infection. Infection and Immunity **2000**; 68:2309–2314.
21. Bong CTH, Harezlak J, Katz BP, Spinola SM. Men Are More Susceptible Than Women to Pustule Formation in the Experimental Model of Haemophilus ducreyi Infection. Sexually Transmitted Diseases **2002**; 29:114–118.
22. Vallejo AF, García J, Amado-Garavito AB, Arévalo-Herrera M, Herrera S. Plasmodium vivax gametocyte infectivity in sub-microscopic infections. Malaria Journal **2016**; 15:48.
23. Black RE, Levine MM, Clements ML, et al. Prevention of Shigellosis by a Salmonella typhi-Shigella sonnei Bivalent Vaccine. The Journal of Infectious Diseases **1987**; 155:1260–1265.
24. Bennett JW, Pybus BS, Yadava A, et al. Primaquine Failure and Cytochrome P-450 2D6 in Plasmodium vivax Malaria. http://dx.doi.org/10.1056/NEJMc1301936. 2013; Available at: https://www.nejm.org/doi/10.1056/NEJMc1301936. Accessed 7 December 2021.
25. Hoffman SL, Goh LML, Luke TC, et al. Protection of Humans against Malaria by Immunization with Radiation-Attenuated Plasmodium falciparum Sporozoites. The Journal of Infectious Diseases **2002**; 185:1155–1164.
26. Hewson CA, Haas JJ, Bartlett NW, et al. Rhinovirus induces MUC5AC in a human infection model and in vitro via NF-κB and EGFR pathways. European Respiratory Journal **2010**; 36:1425–1435.

27. Reeck A, Kavanagh O, Estes MK, et al. Serological Correlate of Protection against Norovirus-Induced Gastroenteritis. The Journal of Infectious Diseases **2010**; 202:1212–1218.
28. Atmar RL, Bernstein DI, Lyon GM, et al. Serological Correlates of Protection against a GII.4 Norovirus. Clinical and Vaccine Immunology **2015**; 22:923–929.
29. Clements ML, Betts RF, Tierney EL, Murphy BR. Serum and nasal wash antibodies associated with resistance to experimental challenge with influenza A wild-type virus. Journal of Clinical Microbiology **1986**; 24:157–160.
30. Van de Verg L, Herrington DA, Murphy JR, Wasserman SS, Formal SB, Levine MM. Specific immunoglobulin A-secreting cells in peripheral blood of humans following oral immunization with a bivalent Salmonella typhi-Shigella sonnei vaccine or infection by pathogenic S. sonnei. Infection and Immunity **1990**; 58:2002–2004.
31. Gwaltney JM Jr, Hendley JO, Patrie JT. Symptom Severity Patterns in Experimental Common Colds and Their Usefulness in Timing Onset of Illness in Natural Colds. Clinical Infectious Diseases **2003**; 36:714–723.
32. Friedman-Klabanoff DJ, Laurens MB, Berry AA, et al. The Controlled Human Malaria Infection Experience at the University of Maryland. The American Journal of Tropical Medicine and Hygiene **2019**; 100:556–565.
33. Palmer KL, Schnizlein-Bick CT, Orazi A, et al. The Immune Response to Haemophilus ducreyi Resembles a Delayed-Type Hypersensitivity Reaction throughout Experimental Infection of Human Subjects. The Journal of Infectious Diseases **1998**; 178:1688–1697.
34. Mayo-Smith LM, Simon JK, Chen WH, et al. The Live Attenuated Cholera Vaccine CVD 103-HgR Primes Responses to the Toxin-Coregulated Pilus Antigen TcpA in Subjects Challenged with Wild-Type Vibrio cholerae. Clinical and Vaccine Immunology 24:e00470-16.
35. Jin C, Gibani MM, Pennington SH, et al. Treatment responses to Azithromycin and Ciprofloxacin in uncomplicated Salmonella Typhi infection: A comparison of Clinical and Microbiological Data from a Controlled Human Infection Model. PLOS Neglected Tropical Diseases **2019**; 13:e0007955.

### Appendix H - Serious Adverse Events

Detailed descriptions of all SAEs extracted are presented below (and are online here). Some SAEs that aren't related to challenge are presented as well.

| File Name | Pathogen Category | Number with ≥1 SAE | Challenge related? | SAE(s) Described | Outcomes | Long-term follow-up |
|---|---|---|---|---|---|---|
| Arévalo-Herrera 2014 | Malaria | 1 | No | "All volunteers successfully recovered clinically within 2–3 days without any serious AE. None of the volunteers required hospitalization for malaria. A volunteer presented with severe diarrhea and abdominal pain (Grade 4) on day 18 and was diagnosed with parasitic gastroenteritis (hookworms and trichuriasis) and was held under observation for <24 hours in the hospital, requiring intravenous rehydration and pain management." | ND | ND |
| Atmar 2014 | Norovirus | 1 | No | "There was 1 serious adverse event during the study and it was assessed as unrelated. A placebo recipient developed a partial retinal detachment 74 days after receipt of study product." | ND | ND |
| Barroso 2005 | Influenza | 1 | Yes | "One serious adverse event was reported after a subject completed the study. A 21 year old black male developed a dilated cardiomyopathy, which resolved with angiotensin-converting enzyme-inhibitor treatment. As described elsewhere [19], this event was considered | ND | "He was clinically stable with low normal cardiac output on echocardiography when last seen in |

| | | | | | | |
|---|---|---|---|---|---|---|
| | | | | possibly related to the experimental influenza B infection but very unlikely related to the study drug." | | 2005, nearly five years after the event." |
| Bastiaens 2016 | Malaria | 1 | Yes | Cardiac serious AE: probable case of acute myocarditis 12 days after CHMI and 1 day after diagnosis of and initiation of treatment for malaria. "The occurrence of the cardiac event relates in time with residual parasitaemia during curative Malarone treatment post-CHMI that might be suggestive of a causal relationship." "A number of possible aetiological factors may explain or have contributed to this case of myocarditis including, i) P. falciparum infection, ii) rhinovirus infection, iii) unidentified pathogens, iv) hyper-immunization (the volunteer received six travel vaccines between the last immunization and the CHMI), v) atovaquone/proguanil treatment, or vi) a combination of these factors. Definitive aetiology and pathophysiological mechanism for the myocarditis have not been established." | "Troponin T levels were normal within 16 days and the volunteer recovered without clinical sequelae." | "Follow-up cardiac MRI at almost five months showed normal function of both ventricles and disappearance of oedema. Delayed enhancement of subepicardial and midwall regions decreased, but was still present." |
| Bennett 2016 | Malaria | 1 | No | "One SAE (ductal carcinoma in situ of the breast) occurred during the study and it was determined that the event was not related to the study vaccine or CHMI." | ND | ND |
| Bernstein 2015 | Norovirus | 4 | Yes | "There were no SAEs reported throughout the study period (excluding severe acute gastroenteritis due to challenge virus)." We count 4 instances of severe vomiting and/or diarrhea as SAEs based on this quote. | ND | "Serious adverse events (SAEs) were collected for 1 year after last study dose." |

| | | | | | | |
|---|---|---|---|---|---|---|
| Black 1982 | E. coli | 2 | Yes | "Two TMP-treated volunteers appeared to suffer clinical relapses 84 and 110 hr, respectively, after recovery from the initial diarrheal illness; in both cases this second illness began 36-48 hr after the first isolation of TMP-resistant E. coli from their stools. In one case this relapse was severe, resulting in over 5 liters of stool and vomitus in less than 12 hr and necessitating iv fluid replacement." | ND | ND |
| Bodhidatta 2012 | Shigella | 2 | Yes | "At Day 14 visit, one subject in Group 1 and one in Group 3 experienced a single elevation of total bilirubin of 2.65 and 1.79 mg/dL, respectively, a serious adverse event classified as grade IV according to the Center for Biologics Evaluation and Research (CBER) guidelines. However, neither patient showed other associated signs and symptoms." | "By Day 28, the total bilirubin levels returned to normal without treatment." | "All subjects reported no abnormalities on Day 42 by telephone assessments." |
| Darsley 2012 | E. coli | 1 | No | "There were no serious AEs following vaccination, but one occurred in a placebo recipient during long-term follow-up postchallenge. This event was not considered to be related to participation in the study." | ND | ND |
| Deasy 2015 | Neisseria lactamica | 4 | No | "Four participants experienced serious adverse events but none were related to the study." | ND | ND |
| DeVincenzo 2020 | RSV | 1 | Yes | "One SAE (acute myocarditis) was reported." | ND | ND |
| Dobinson 2017 | Salmonella paratyphi | 1 | No | "One serious adverse event was recorded (appendicitis 6 months after S. Paratyphi A challenge) and was assessed as being unrelated to study procedures." | ND | ND |

| | | | | | | |
|---|---|---|---|---|---|---|
| Frenck 2021 | Shigella | 1 | No | "In the Placebo group, one participant experienced four SAEs following the challenge dose: deep vein thrombosis (moderate intensity) and pelvic venous thrombosis (severe) at day 107, haematoma (severe) at day 121, and carotid artery aneurysm at day 130 from challenge." | "All events were assessed by the investigator as unrelated to any study treatment and were resolved." | ND |
| Fullen 2016b | Rhinovirus | 1 | No | "One serious adverse event (SAE) was reported in the 1 TCID50 group. The SAE was a Grade 4 self-limiting raised creatine phosphokinase considered to be of medical importance, but not considered to be related to the Challenge Virus, study procedures, concomitant medications or study assessments." | "No action was taken and the SAE resolved." | ND |
| Gibani 2019 | Salmonella typhi | 1 | No | "Generalised tonic clonic seizure": "Occurred 9 months after completion of challenge. Reported a history of childhood seizures that was not disclosed at screening or on available medical records. Commenced on anticonvulsants and ongoing follow up with neurology at end of study" | ND | ND |
| Gibani 2019 | Salmonella typhi | 1 | No | "Deliberate self harm": "Episode of deliberate self-harm 6 months after completion of challenge, attributed to an acute life stressor. No specific treatment was administered." | ND | ND |
| Gibani 2020 | Salmonella typhi | 1 | Yes | "Hospital admission for IV fluids": "Nausea and vomiting, unresponsive to oral anti-emetics. Tachycardia. Admitted overnight for IV fluids and anti-emetics. Treatment switched to IV Ceftriaxone." | "Discharged after <24 hours and completed course of oral ciprofloxacin." | ND |

| | | | | | | |
|---|---|---|---|---|---|---|
| Gibani 2020 | Salmonella typhi | 1 | Yes (Rechallenge, not counted in main paper table of results) | "Raised ALT": "Alanine aminotransferase elevated to 898 IU/L 5 days after diagnosis. Ascribed to paratyphoid fever plus possible adverse drug reaction (azithromycin + paracetamol). Antibiotic switched from azithromycin to ciprofloxacin. Paracetamol withheld." | "Resolved." | ND |
| Gibani 2020 | Salmonella typhi | 1 | No | "Episode of collapse with loss of consciousness 12 months after challenge. Reported left arm and left leg weakness 24 hours' duration. Diagnosed as possible generalised seizure following neurology review. Instructed not to drive for 6/12." | "No medication prescribed, and no subsequent events." | ND |
| Gibani 2020 | Salmonella typhi | 1 | No | "Right loin pain with evidence of renal calculus on CT KUB. Diagnosed after enrolment but prior to challenge. Withdrawn from study and not challenged." | ND | ND |
| Graham 1983 | E. coli | 4 | Yes | "four subjects became sufficiently ill that they received adjunctive therapy (defined as therapy in addition to the randomized drugs used in the study): intravenous fluids with antinauseants or tetracycline, or both." | ND | ND |
| Jin 2017 | Salmonella typhi | 1 | No | "One Vi-TT participant was diagnosed with inflammatory bowel disease and withdrawn from the study (symptoms preceded study enrolment)" | ND | ND |
| Jin 2017 | Salmonella typhi | 2 | No | "two Vi-PS participants were hospitalised (urinary retention and semi-elective tonsillectomy)" | ND | ND |
| Jin 2017 | Salmonella typhi | 1 | Maybe (counted in main paper table of | "one Vi-PS participant was diagnosed with reactive arthritis possibly related to S. Typhi challenge or antibiotic treatment." | ND | ND |

| | | | | | | |
|---|---|---|---|---|---|---|
| | | | results) | | | |
| Lyke 2015 | Malaria | 1 | Yes | "Subject M11UMD012 (Group C: 1 × 104 PfSPZ 2 × 10 µL) was noted to be qPCR positive on Day 28 after therapy and was reported and assessed as a Serious Adverse Event by the sponsor Medical Monitor. Following ID CHMI, a 44-year-old, male volunteer developed the expected outcome of falciparum malaria on Day 15 post-CHMI with a parasite density of 270 parasites/µL by smear. This volunteer missed one interval appointment on the day before detection. He was treated with a total of 3 g of rapid-acting, directly observed atovaquone-proguanil (Malarone) over 3 days. On Day 22 post-CHMI, qPCR and malaria smears were negative. On Day 28 post-CHMI, a routine study qPCR returned positive (295 parasites/mL); however, the malaria smear was negative and the subject was asymptomatic. A repeat qPCR on Day 33 (the day after initial qPCR positive results were forwarded to investigators) post-CHMI was borderline positive (31 parasites/mL, lower limit of detection 20–40), and the subject remained asymptomatic. On Day 35 post-CHMI, the qPCR and malaria smear results were negative." | "Subsequent malaria smears and qPCR remained negative and no evidence of asexual or replicating stages were found. Terminal CQ treatment was administered per study sponsor request." | "A P. falciparum culture (considered the gold standard for detection of parasites) was performed from blood obtained on Day 28 and remained negative for 4 weeks after incubation." |
| McCarthy 2016a | Malaria | 1 | No | "The only serious adverse event (recurrent tonsillitis) was considered unrelated to the clinical trial treatment." | ND | ND |

| | | | | | | |
|---|---|---|---|---|---|---|
| McCarthy 2016c | Malaria | 3 | Maybe (counted in main paper table of results) | "This study was put on hold following the occurrence of these three separate events occurring in three subjects that were classified as serious adverse events (SAEs) by the sponsor [...] In the absence of the identification of another plausible cause, and with the temporal association following ferroquine administration it was concluded that these adverse events were likely attributable to ferroquine. However, given the concurrence of a range of other potential hepatic irritants (notably malaria, and concurrent medications), it not possible to exclude these as cofactors." | ND | ND |
| Rickman 1990 | Malaria | 1 | Yes | "Subject no. 1 was the only subject hospitalized. He was hospitalized overnight and was treated only with acetaminophen in addition to chloroquine. His platelet count dropped from 225,000/ul to 126.000/ul 2 days after chloroquine treatment began. This subject also developed a mild leukopenia (3,300 WBC/ul) and had pyuria (5--10 WBC/HPF) and microscopic hematuria (5--10 RBC/HPF) for 8 hr without evidence of renal insufficiency." | ND | ND |

| | | | | | | |
|---|---|---|---|---|---|---|
| Roestenberg 2013 | Malaria | 1 | Yes | "One SAE occurred in a volunteer who reported chest pain 1 day after the first dose of atovaquone/proguanil. Based on medical history, the chest pain was initially considered possibly consistent with angina pectoris." | "Pain resolved within 1 hour without treatment. The volunteer was admitted to the cardiac care unit for monitoring for 6.5 hours. The first electrocardiogram (ECG) had a negative T-wave in V2, which was absent at the time of study initiation. All subsequent ECGs, beginning 2.5 hours after the first ECG, were comparable to baseline, with a negative T in V1 only. Troponin T levels were normal at the time of chest pain, 6 and 17 hours later, daily for the next 3 days and at trial Days 28 and 35." | ND |
| Seder 2013 | Malaria | 1 | No | "One subject was diagnosed with colon cancer and did not participate in CHMI." | ND | ND |

| | | | | | | |
|---|---|---|---|---|---|---|
| Seder 2013 | Malaria | 1 | No | "After CHMI, one vaccine recipient developed anxiety and headache 14 days after completing chloroquine treatment and was hospitalized overnight for diagnostic purposes." | "The symptoms, which were attributed to chloroquine, resolved within 1 month." | ND |
| Smith 2017 | Malaria | 1 | No | "One adverse event (renal colic) was classified as serious; however, this event was determined not to be related to study treatments." | "All adverse events had resolved by the end of study." | ND |
| Sulyok 2017 | Malaria | 1 | No | "During follow-up, one serious adverse event occurred in cohort 1A: a bilateral pulmonary embolism occurred in a non-smoking woman under oral contraceptive treatment 34 days after DSM265 administration while she was on a transatlantic flight." | "The volunteer never became parasitaemic and recovered fully, except for a grade 1 sinus tachycardia, which still persisted at the last follow-up visit." | ND |

### Appendix I - Other

Many studies reported individual AEs by symptom or total individual AEs rather than the number of participants that experienced at least one AE. To handle this unclear AE data, the number of participants that must have experienced at least one AE based on the data that was provided was derived and extracted as a minimum value, and the lesser of the sum or the total number of participants challenged was extracted as a maximum value.

Assumptions about AEs:

- Schiff 2000: unclear whether AEs were treatment related or challenge related; counted as challenge related to avoid potential underestimate (deemed more problematic than potential overestimate)

- Sulyok 2017: unclear whether AEs were treatment related or challenge related; counted as challenge related to avoid potential underestimate (deemed more problematic than potential overestimate)
- Turner 1999: unclear whether AEs were treatment related or challenge related; counted as challenge related to avoid potential underestimate (deemed more problematic than potential overestimate)
- Turner 2001: unclear whether AEs were treatment related or challenge related; counted as challenge related to avoid potential underestimate (deemed more problematic than potential overestimate)
- Turner 2005: unclear whether AEs were treatment related or challenge related; counted as challenge related to avoid potential underestimate (deemed more problematic than potential overestimate)

Study Counts:

- As described in the paper, an individual "study" is identified by trial registration in a database (such as ClinicalTrials.gov), or per the article description if trial registration is not reported. Without universal trial registration, counting the number of "studies" based on what is reported in published articles is an inexact process. Articles often report multiple cohorts challenged at different times and with different strains within the pathogen being investigated, making it difficult to ascertain whether multiple "studies" occurred, or whether all challenges were part of a single "study." It is understood that the number of studies described in the dataset used for this review may reasonably be counted differently by following different methodology; however, this does not impact data on challenged participants.

Data summing:

- Data were combined for Rylance 2019 and Hales 2020 (discussed same study but reported slightly different data about it)
- Data were combined for Lyke 2010 and Laurens 2013 (same ClinicalTrials registration number, each paper reported data for different cohorts)
- Data were combined for Atmar 2008 and Atmar 2014 (updated data from 2014 wasn't found in searches, but necessary to properly extract)

Safety concerns:

- Al-Tawfiq 1998: "Three volunteers were inoculated with a mixed culture of H. ducreyi and Pseudomonas aeruginosa"
- Feary 2009: "11 of the 13 participants who completed the study chose not to take the treatment, citing either a perceived improvement in hayfever symptoms, or that they wished to see if they had a change in symptoms the following year. In accordance with our protocol, participants in the placebo group were offered hookworm infection at the end of the trial, of whom 11 chose to be infected."

- Fortney 2000: “Three volunteers were excluded from the analysis because they were inadvertently inoculated with a culture that was contaminated with Candida parapsilosis”
- Peterson 2009: Notes AEs “linked to recognized effects of HRV infections (for example, earache, pharyngitis and headache)” but does not provide detail on them

### Reviewer Guidelines

Note that the guidelines are included below, but annotated studies are not included due to copyright issues.
