## Supplementary material for "A Systematic Review of Human Challenge Trials, Designs, and Safety": Review workflow

### Systematic Review Workflow

Danny Toomey – 1Day Sooner

### Purpose – why do a systematic review?

- A systematic review is a study of studies, in which case:
  - Our methods are how we found the studies we chose to analyze and why we decided those studies were appropriate to analyze
  - Our results are what those studies found in aggregate and in context with one another
- What kind of questions can a systematic review answer?
  - What is the consensus of a given field on a given topic?
  - What results have been reliably found on a given outcome of interest?
  - What gaps in knowledge are frequently observed on a given topic?
- What are the questions we want to answer on this project?
  - Are human challenge trials dangerous?
  - Are serious adverse events frequently reported?
  - Are there any important pieces of information frequently omitted by studies of human challenge trials?
- The following slides will detail an example systematic review workflow that will be supplemented by a spreadsheet

### Systematic review workflow - Search

- On Example Spreadsheet, see column F rows 3-6 on sheet “Search Results” for reporting an example search. The exact algorithm used, databased accessed, date accessed, and number of results are listed. This is useful for reporting the search and cross-referencing results at later dates.
- Columns A-D are copied from the CSV output from the PubMed search.

### Systematic review workflow - Screening

- On Example Spreadsheet, see column I rows 2-3 on sheet “Screening” for some example inclusion and exclusion criteria I used to screen these papers. The actual inclusion and exclusion criteria we use will be different.
- I did not go through all the results from the search and I edited the screened results a little for example’s sake.
- Columns A-C are copied from the Search Results tab. Column D is a formula that creates a PubMed URL for each study. My preferred screening workflow is to paste the URL into a browser to read the title and abstract. I like this workflow because it’s fast and you can do most of what you need to do with keyboard shortcuts.
- Columns E and F note whether the paper is excluded and, if so, the reason for exclusion. These reasons are based off reading the title and abstract alone. Another thing I like about the PubMed workflow is that it will say in the top left corner if a paper is a review or case report, which saves a lot of time on reading in the long run.
- Column G notes whether the paper is eligible for full text review based on title and abstract alone. Some papers will be clearly eligible, some will not. I personally think it’s better to mark a paper that may be useful as eligible and determine that isn’t useful down the road, rather than miss a potentially good paper.

### Systematic review workflow – Full text review

- On Example Spreadsheet, see sheet “Full-text Review.” You’ll notice it looks similar to the Screening tab, except the only papers included are the papers marked ‘yes’ under ‘Eligible for full-text review?’ from the Screening tab.
- Column F details the reasons for exclusion after full-text review. These reasons will often be more detailed and nuanced than the reasons for exclusion during screening.
- The inclusion and exclusion criteria always stay the same. The only thing that changes is the level of detail required to exclude.
- If it makes the cut, mark it for inclusion!

### Included papers – annotated readings

- The following slides will provide screenshots from annotated readings of Gerding 2015 and Liebowitz 2020
- Gerding 2015 is included in the set of papers previously collected
- Liebowitz 2020 is a new paper I found from the results I screened

### Annotated reading – Gerding 2015, Abstract

1

**IMPORTANCE** *Clostridium difficile* is the most common cause of health care–associated infection in US hospitals. Recurrence occurs in 25% to 30% of patients.

2

**OBJECTIVE** To determine the safety, fecal colonization, recurrence rate, and optimal dosing schedule of nontoxigenic *C difficile* strain M3 (VP20621; NTCD-M3) for prevention of recurrent *C difficile* infection (CDI).

3

**DESIGN, SETTING, AND PARTICIPANTS** Phase 2, randomized, double-blind, placebo-controlled, dose-ranging study conducted from June 2011 to June 2013 a[illegible] 4 [illegible]g 173 patients aged 18 years or older who were diagnosed as having CDI (first episode or fi[illegible] [illegible]currence) and had successfully completed treatment with metronidazole, oral vancomycin, or both at 44 study centers in the United States, Canada, and Europe.

5

**INTERVENTIONS** Patients were randomly assigned to receive 1 of 4 treatments: oral liquid formulation of NTCD-M3, $10^4$ spores/d for 7 days (n = 43), $10^7$ spores/d for 7 days (n = 44), or $10^7$ spores/d for 14 days (n = 42), or placebo for 14 days (n = 44).

**RESULTS** A 6 [illegible] 168 patients who started treatment, 157 completed treatment. One or more treatment-eme 7 [illegible] adverse events were reported in 78% of patients receiving NTCD-M3 and 86% of pa[illegible]receiving placebo. Diarrhea and abdominal pain were reported in 46% and 17% of patients receiving NTCD-M3 and 60% and 33% of placebo patients, respectively. 8 Serious treatment-emergent adverse events were reported in 7% of patients receiving placebo and 3% of all patients who received NTCD-M3. 9 Headache was reported in 10% patients receiving NTCD-M3 and 2% of placebo patients. Fecal colonization occurred in 69% of NTCD-M3 patients: 71% with $10^7$ spores/d and 63% with $10^4$ spores/d. Recurrence of CDI occurred in 13 (30%) of 43 placebo patients and 14 (11%) of 125 NTCD-M3 patients (odds ratio [OR], 0.28; 95% CI, 0.11-0.69; *P* = .006); the lowest recurrence was in 2 (5%) of 43 patients receiving $10^7$ spores/d for 7 days (OR, 0.1; 95% CI, 0.0-0.6; *P* = .01 vs placebo]). Recurrence occurred in 2 (2%) of 86 patients who were colonized vs 12 (31%) of 39 patients who received NTCD-M3 and were not colonized (OR, 0.01; 95% CI, 0.00-0.05; *P* < .001).

10

**TRIAL REGISTRATION** clinicaltrials.gov Identifier: NCT01259726

*JAMA*. 2015;313(17):1719-1727. doi:10.1001/jama.2015.3725 11

1

| | O |
|---|---|
| 1 | Disease |
| 2 | Clostridium difficile |

2, 3

| | I |
|---|---|
| 1 | Study Type (RCT?) |
| 2 | RCDoseRanging |

2

| | Q | R |
|---|---|---|
| 1 | Attenuated or full disease | If Attenuated, how |
| 2 | Attenuated | Utilized non-toxic C. difficile strain NTCD-M3 |

4

| | AE | AF |
|---|---|---|
| 1 | SCREENING: # of people | SCREENING: Age MIN |
| 2 | 173 | 18 |

5

| | N |
|---|---|
| 1 | Route (oral, shot, nasal, aerososol, intradermal) |
| 2 | oral |

8

| | AP |
|---|---|
| 1 | DRUG: Name (If Administered) |
| 2 | NTCD-M3 |

5

| | M |
|---|---|
| 1 | Dose if listed |
| 2 | 10^4 - 10^7 spores |

6

| | P |
|---|---|
| 1 | Size of cohort |
| 2 | 168 |

7

| | U |
|---|---|
| 1 | Fraction of test group developing symptoms |
| 2 | 0.784 |

9

| | S |
|---|---|
| 1 | Fraction of test group developing "severe" symptoms |
| 2 | 0.032 |

10

| | H |
|---|---|
| 1 | Study Clinical Trial Number |
| 2 | NCT01259726 |

11

| | L |
|---|---|
| 1 | Study DOI |
| 2 | 10.1001/jama.2015.3725 |

### Annotated reading – Gerding 2015, Introduction

*Clostridium diffici*[illegible] 1 [illegible]n anaerobic spore-forming bacterium, **is the cause of one of the most common and deadly health care-associated infections**, linked to 29 000 US deaths each year.[1] Rates of *C difficile* infection (CDI) remain at unprecedented high levels in US hospitals, and *C difficile* is the most commonly identified health care pathogen.[1,2] Patients, especially elderly people taking antibi-

Gastrointestinal colonization of patients and hamsters by these nontoxigenic *C difficile* strains has been shown to prevent CDI with exposure to a toxigenic strain 2 **One of these NTCD strains, M3 (VP20621; NTCD-M3), has been shown to safely colonize volunteers aged 60 years or older when given at doses ranging from $10^4$ to $10^8$ spores/d for 14 days following 5 days of vancomycin to disrupt the normal microbiota and simulate CDI treatment.**[9] Herein, we report results of a phase 2 trial of the safety and efficacy of NTCD-M3 spores in colonizing and preventing recurrent CDI following successful treatment of the first episode or first recurrence of CDI.

1

| | AD | |
|---|---|---|
| 1 | ENDEMICITY - is the disease endemic? Volunteers might have already had it or have a background risk of having gotten it before. | S # |
| 2 | Yes | 1 |

2

| | AP | |
|---|---|---|
| 1 | DRUG: Name (If Administered) | N D |
| 2 | NTCD-M3 | |

2

| | R | |
|---|---|---|
| 1 | If Attenuated, how | F g " |
| 2 | Utilized non-toxic C. difficile strain NTCD-M3 | C |

### Annotated reading – Gerding 2015, Methods

etiology in the opinion of the investigator. The qualifying episode of CDI was either a primary episode or a first recurrence 8 weeks or sooner after primary [1] Severe CDI was defined as 10 or more unformed stools per day or white blood cell count of 15 000/μL or higher. Patients were treated with metronidazole, oral vancomycin, or both for 10 to 21 days and had clini- [2]

Exclusion criteria included the following: more than 1 episode of CDI (other than qualifying episode) 6 months or less before randomization; qualifying episode treated with any antimicrobial other than metronidazole or oral vancomycin; treatment with immunotherapy (eg, intravenous immu-

**Study Design and Interventions**

This randomized, double-blind, placebo-controlled, dose-ranging st [3] was performed from June 2011 to June 2013 at 44 study cen [4] (33 in the United States, 7 in Europe, and 4 in Canada). Patients were randomly assigned to 1 of 4 different

of etiology) were included in the safety analy [5] Serious treatment-emergent adverse events included death, a life-threatening event, inpatient hospitalization or prolongation of an existing hospitalization, a persistent or significant incapacity or disruption of normal life functions, a congenital anomaly or birth defect, and other medically important events that jeopardized a patient or may have required intervention to prevent any of the above serious outcomes occurring through week 26.

1

| | AI |
|---|---|
| 1 | SCREENING: Antibody titers taken |
| 2 | No |

1

| | Z | AA |
|---|---|---|
| 1 | Number of test group showing antibodies | Notes on antibody testing |
| 2 | ND | White blood cell count was used to determine participant's response to pathogen, not antibodies |

2

| | AK | AL | AM |
|---|---|---|---|
| 1 | SCREENING: Lack of a condition to accept? | SCREENING: Presence of a condition to accept? | Disease limitation |
| 2 | Yes | No | No symptoms |

5

| | T |
|---|---|
| 1 | Notes on severe symptoms |
| 2 | "Serious treatment-emergent adverse events included death, a life-threatening event, inpatient hospitalization or prolongation of an existing hospitalization, a persistent or significant incapacity or disruption of normal life functions, a congenital anomaly or birth defect, and other medically important events that jeopardized a patient or may have required intervention to prevent any of the above serious outcomes occurring through week 26." |

5

| | V |
|---|---|
| 1 | Symptoms defined as adverse events |
| 2 | death, a life-threatening event, inpatient hospitalization or prolongation of an existing hospitalization, a persistent or significant incapacity or disruption of normal life functions, a congenital anomaly or birth defect |

1, 5

| | AC |
|---|---|
| 1 | Microbiological vs Clinical: Infection model vs Disease Model |
| 2 | Both |

3

| | AN |
|---|---|
| 1 | Trial Actually Conducted yet? |
| 2 | Yes |

4

| | AH |
|---|---|
| 1 | SCREENING: Population Locale |
| 2 | USA,EU,CA |

### Annotated reading – Gerding 2015, Results

1

| CDI qualifying episode data | | | | |
|---|---|---|---|---|
| Age, median (range), y | 58 (18-90) | 58 (22-89) | 58 (21-94) | 64 (20-90) |
| Female | 26 (61) | 26 (63) | 24 (56) | 28 (68) |

1

| | AJ |
|---|---|
| 1 | SCREENING: Just one sex? |
| 2 | No |

1

| | AF | AG |
|---|---|---|
| 1 | SCREENING: Age MIN | SCREENING: Age MAX |
| 2 | 18 | 94 |

2

| Adverse Events in Intention-to-Treat Safety Population | Placebo (n = 43) | NTCD-M3 Dosage: $10^4$ Spores/d for 7 d (n = 41) | NTCD-M3 Dosage: $10^7$ Spores/d for 7 d (n = 43) | NTCD-M3 Dosage: $10^7$ Spores/d for 14 d (n = 41) | All (n = 125) |
|---|---|---|---|---|---|
| ≥1 Treatment-emergent adverse event[a] | 37 (86) | 33 (80) | 34 (79) | 31 (76) | 98 (78) |

2

| | W |
|---|---|
| 1 | Fraction of control group developing symptoms |
| 2 | 0.860465116 |

2

| | U |
|---|---|
| 1 | Fraction of test group developing symptoms |
| 2 | 0.784 |

3

| | | | | | |
|---|---|---|---|---|---|
| ≥1 Serious treatment-emergent adverse event[d] | 3 (7) | 1 (2) | 1 (2) | 2 (5) | 4 (3) |
| Deaths | 1 (2) | 0 | 0 | 1 (2) | 1 (1) |

[d] See Methods section of text for definition of serious treatment-emergent adverse events.

3

| | X | Y |
|---|---|---|
| 1 | Number of test group developing serious adverse events | Number of control group developing serious adverse events |
| 2 | 4 | 3 |

4

| Participants, No./Total (%) | Placebo: Culture Positive, Toxin Negative (NTCD) | Placebo: Culture Positive, Toxin Positive | NTCD-M3, $10^4$ Spores/d for 7 d: Culture Positive, Toxin Negative (NTCD) | NTCD-M3, $10^4$ Spores/d for 7 d: Culture Positive, Toxin Positive | NTCD-M3, $10^7$ Spores/d for 7 d: Culture Positive, Toxin Negative (NTCD) | NTCD-M3, $10^7$ Spores/d for 7 d: Culture Positive, Toxin Positive | NTCD-M3, $10^7$ Spores/d for 14 d: Culture Positive, Toxin Negative (NTCD) | NTCD-M3, $10^7$ Spores/d for 14 d: Culture Positive, Toxin Positive |
|---|---|---|---|---|---|---|---|---|
| Week 6 | 4/39 (10) | 13/39 (33) | 14/39 (36) | 2/39 (5) | 20/41 (49) | 4/41 (10) | 10/37 (27) | 7/37 (19) |

[a] The first 2 weeks were the NTCD-M3 or placebo treatment period. End of treatment to week 6 was the period used to assess NTCD-M3 colonization.

4

| | AB |
|---|---|
| 1 | Infection rate (how many volunteers do you need if only 30% or something will actually get it when exposed? |
| 2 | 0.307692308 |

### Annotated reading – Liebowitz 2020, abstract

Backg[illegible] 1 Influenza is an important public health problem and existing vaccines are not completely protective. New vaccines that pr[illegible]by alternative mechanisms are needed to improve efficacy of influenza vaccines. In 2015, we did a phase 1 trial 2 [illegible] oral influenza vaccine, VXA-A1.1. A favourable safety profile and robust immunogenicity results in that trial supported progression of the vaccine to the current phase 2 trial. The aim of this study was to evaluate efficacy of the vaccine in a human influenza challenge model.

4 [illegible]ods We did a singl 3 [illegible], placebo-controlled and active-controlled, phase 2 study at WCCT Global, Costa Mesa, CA, [illegible] Eligible individuals had an initial A/California/H1N1 haemagglutination inhibition titre of less than 20 and were aged 18–49 years and in good health. Individuals were randomly assigned (2:2:1) to receive a single immunisation of 5 either $10^{11}$ infectious units of VXA-A1.1 (a monovalent tablet vaccine) orally, a full human dose of quadrivalent inactivated influenza vaccine (IIV) via intramuscular injection, or matched placebo. Randomisation was done by computer-generated assignments with block size of five. An unmasked pharmacist provided the appropriate vaccines and placebos [illegible] investigator-assessed contraindicat 6 [illegible] were challenged intranasally with 0·5 mL wild-type A/CA/like(H1N1)pdm09 influenza virus. The primary outcomes were safety, which was assessed in all immunised participants through 365 days, and influenza-positive illness after viral challenge, which was assessed in individuals that received the viral challenge and the required number of assessments post viral challenge. This trial is registered with ClinicalTrials.gov, number 7 NCT02918006.

Res 8 Between Aug 31, 2016, and Jan 23, 2017, 374 individuals were assessed for eligibility, of wh 9 179 were randomly assigned to receive either VXA-A1.1 (n=71 [one individual did not provide a diary card, thus the solicited events were assessed in 70 individuals]), IIV (n=72), or placebo (n=36). Between Dec 2, 20 10 [illegible]d April 26, 2017, 143 eligible individuals (58 in the VXA-A1.1 group, 54 in the IIV group, and 31 in the placebo [illegible]) were challenged with influenza virus. VXA-A1.1 was well tolerated with no serious or medically significant adverse events. The most prevalent solicited adverse events for each of the treatment groups after immunisat 11 [illegible]re headache in the VXA-A1.1 (in five [7%] of 70 participants) and placebo (in seven [19%] of 36 participants) grou[illegible] tenderness at injection site in the IIV group (in 19 [26%] of 72 participants). Influenza-positive illness after challenge was detected in 17 (29%) of 58 individuals in the VXA-A1.1 group, 19 (35%) of 54 in the IIV group, and 15 (48%) of 31 in the placebo group. 12

Lancet Infect Dis 2020
Publis[illegible] 13
Janua[illegible]
https://doi.org/10.1016/S1473-3099(19)30584-5

1

| | O |
|---|---|
| 1 | Disease |
| 2 | influenza |

2

| | N |
|---|---|
| 1 | Route (oral, shot, nasal, aerososol, intradermal) |
| 2 | oral |

13

| | L |
|---|---|
| 1 | Study DOI |
| 2 | 10.1016/S1473-3099(19)30584-5 |

3

| | I |
|---|---|
| 1 | Study Type (RCT?) |
| 2 | RCT |

4

| | AF | AG | AH | AI |
|---|---|---|---|---|
| 1 | SCREENING: Age MIN | SCREENING: Age MAX | SCREENING: Population Locale | SCREENING: Antibody titers taken |
| 2 | 18 | 49 | USA | yes |

4

| | AK | AL | AM | AN | AO |
|---|---|---|---|---|---|
| 1 | SCREENING: Lack of a condition to accept? | SCREENING: Presence of a condition to accept? | Disease limitation | Trial Actually Conducted yet? | VACCINE: Name (If tested) |
| 2 | yes, needed HAI titre of less than | no | none | yes | VXA-A1.1 |

6

| | Q |
|---|---|
| 1 | Attenuated or full disease |
| 2 | full |

12

| | AB |
|---|---|
| 1 | Infection rate (how many volunteers do you need if only 30% or something will actually get it when exposed? |
| 2 | 0.356643357 |

8

| | AE |
|---|---|
| 1 | SCREENING: # of people |
| 2 | 374 |

9

| | P |
|---|---|
| 1 | Size of cohort |
| 2 | 179 |

10

| | S |
|---|---|
| 1 | Fraction of test group developing "severe" symptoms |
| 2 | 0 |

7

| | H |
|---|---|
| 1 | Study Clinical Trial Number |
| 2 | NCT02918006 |

5

| | M |
|---|---|
| 1 | Dose if listed |
| 2 | 10^11 infectious units |

### Annotated reading – Liebowitz 2020, Introduction

**Introduction**

Seasonal influenza continues to be a major public health problem, and is estimated to be associated with 114018–633001 hospitalisations, 18476–96667 intensive care unit admissions, and 4866–27810 deaths per year

| | AD |
|---|---|
| 1 | **ENDEMICITY - is the disease endemic? Volunteers might have already had it or have a background risk of having gotten it before.** |
| 2 | yes |

### Annotated reading – Liebowitz 2020, Methods

**Procedures** 1

Participants in the VXA-A1.1 group received $10^{11}$ infectious units of VXA-A1.1 in a single oral dose (in seven tablets)

eligibility to participate in the viral challenge part of the stu 2 Individuals were excluded if they had clinically significant symptoms or signs of influenza, an oral temperature of higher than 37·9°C, a positive result for respiratory viral shedding on a Biofire test (Biofire, Salt Lake City, UT, USA), or any clinical or laboratory finding that in the opinion of the investigator could affect the participant's safety. Individuals who were not eligible to participate in the viral challenge part of the study were

llenge at day 90 were r 3 essed for eligibility with the subsequent cohort; there[illegible], all individuals in the efficacy analysis were challenged between 90 and 132 days after vaccination.

3

| | AS |
|---|---|
| 1 | **Disease: Time to challenge** |
| 3 | 90 - 132 days after vaccination |

1

| | M |
|---|---|
| 1 | **Dose if listed** |
| 2 | 10^11 infectious units |

2

| | AC |
|---|---|
| 1 | **Microbiological vs Clinical: Infection model vs Disease Model** |
| 2 | Both |

4

| | V |
|---|---|
| 1 | **Symptoms defined as adverse events** |
| 2 | nausea, vomiting, diarrhoea, abdominal pain, localised pain, tenderness, erythema and induration, malaise or fatigue, myalgia, anorexia, fever, and headache |

5

| | T |
|---|---|
| 1 | **Notes on severe symptoms** |
| 2 | "the number of serious adverse events and adverse events of special interest (eg, new onset or exacerbation of specific autoimmune diseases, as listed in the protocol)" |

6

| | AA |
|---|---|
| 1 | **Notes on antibody testing** |
| 2 | Described antibody titres in geometric mean titre |

**Outcomes**

The primary safety en 4 ints were the number of individual reporting solic symptoms (nausea, vomiting, diarrhoea, abdominal pain, localised pain, tenderness, erythema and induration, malaise or fatigue, myalgia, anorexia, fever, and headache) through day 8 after vaccination, the number of unsolicited adverse events through day 30 after receipt of the challenge vi 5 the number of serious adverse events and adverse events of special interest (eg, new onset or exacerbation of specific autoimmune diseases, as listed in the protocol) through

producing cells on day 8, and HAI assay and microneutralisation titres on day 30 after vaccina 6 Geometric mean titres and geometric fold responses were calculated for HAI and microneutralisation responses for each group. Additionally, HAI geometric mean titres were measured on day 90 on subjects that were challenged.

### Annotated reading – Liebowitz 2020, Results

| 1 | VXA-A1.1 (n=70) | IIV (n=72) | Placebo (n=36) |
|---|---|---|---|
| Any symptom | 20 (29%) | 26 (36%) | 15 (42%) |

between groups (appendix pp 4–5). The unsolicited adverse events are for all individuals in the safety data [2] No serious adverse events or adverse events of special interest were reported during the vaccination or challenge phases.

participants who went on to the inpatient challenge phase of the st[3] The HAI geometric mean titre was 31·4 (95% CI 24·1–40·1) in the VXA-A1.1 group, 186·7 (95% CI 126·7–275·2) in the IIV group, and 11·5 (95% CI 8·6–15·4) in the placebo group. Because HAI results differed between IIV and VXA-A1.1, yet the efficacy of the

| 4 | Vaccination phase | | | Challenge phase | | |
|---|---|---|---|---|---|---|
| | VXA-A1.1 (n=71) | IIV (n=72) | Placebo (n=36) | VXA-A1.1 (n=58) | IIV (n=54) | Placebo (n=31) |
| Sex | | | | | | |
| Male | 42 (59%) | 43 (60%) | 21 (58%) | 32 (55%) | 32 (59%) | 18 (58%) |
| Female | 29 (41%) | 29 (40%) | 15 (42%) | 26 (45%) | 22 (41%) | 13 (42%) |

1

| | U |
|---|---|
| 1 | Fraction of test group developing symptoms |
| 2 | 0.285714286 |

1

| | W |
|---|---|
| 1 | Fraction of control group developing symptoms |
| 2 | 0.416666667 |

2

| | X | Y |
|---|---|---|
| 1 | Number of test group developing serious adverse events | Number of control group developing serious adverse events |
| 2 | 0 | 0 |

3

| | Z |
|---|---|
| 1 | Number of test group showing antibodies |
| 2 | Antibody titres are described, but not in terms of # of patients positive for antibodies in each group |

4

| | AJ |
|---|---|
| 1 | SCREENING: Just one sex? |
| 2 | no |

### Closing thoughts

- This stuff is hard! Don’t hesitate to ask for help or a second pair of eyes on anything
